# Public opinion on sharing data from UK health services for clinical and research purposes without explicit consent

**DOI:** 10.1101/2021.07.19.21260635

**Authors:** Linda A. Jones, Jenny R. Nelder, Joseph M. Fryer, Philip H. Alsop, Michael R. Geary, Mark Prince, Rudolf N. Cardinal

**Affiliations:** Department of Psychiatry, University of Cambridge, UK; Unaffiliated; Cambridgeshire and Peterborough NHS Foundation Trust, UK

## Abstract

1.

**BACKGROUND:** In the UK, National Health Service (NHS/HSC) data is variably shared between healthcare organizations for direct care, and increasingly used in de-identified forms for research. Few large-scale studies have examined public opinion on sharing, including the treatment of mental health (MH) versus physical health (PH) data.

**METHODS:** Pre-registered anonymous online survey open to all UK residents, recruiting Feb–Sep 2020. Participants were randomized to one of three framing statements regarding MH versus PH data.

**FINDINGS:** Participants numbered 29275; 40% had experienced a MH condition. A majority supported identifiable data sharing for direct clinical care without explicit consent, but 20% opposed this. Preference for clinical/identifiable sharing decreased with distance and was slightly less for MH than PH data, with a small framing effect. Preference for research/de-identified data sharing without explicit consent showed the same small PH/MH and framing effects, plus greater preference for sharing structured data than de-identified free text. There was net support for research sharing to the NHS, academic institutions, and national research charities, net ambivalence about sharing to profit-making companies researching treatments, and net opposition to sharing to other companies (similar to sharing publicly). De-identified linkage to non-health data was generally supported, except to data held by private companies. We report demographic influences on preference. A clear majority supported a single NHS mechanism to choose uses of their data. Support for data sharing increased during the pandemic.

**INTERPRETATION:** Support for healthcare data sharing for direct care without explicit consent is broad but not universal. There is net support for the sharing of de-identified data for research to the NHS, academia, and the charitable sector, but not the commercial sector. A single national NHS-hosted system for patients to control the use of their NHS data for clinical purposes and for research would have broad public support.

**FUNDING:** MRC.

## 2. Introduction

In the United Kingdom (UK), health-related information is recorded routinely by health care professionals and patients within the National Health Service (NHS; England, Scotland, Wales) or Health and Social Care (HSC; Northern Ireland), henceforth “NHS” for brevity. When combined with personal identifiers such as names and addresses, the data represent “confidential patient information” (CPI),^1^ used to provide care and managed according to standard principles.^2–7^ It is “owned” legally and managed by the NHS organisation recording it.^5, 8^ De-identified or anonymised forms of the data may be used for research ( **Fig.1**) without explicit consent,^5, 6^ as pledged by the NHS.^9^ Identifiable data may be used for research with consent, or— under restricted circumstances—without.^1, 5, 6^ “Fully” anonymised data are not subject to UK data protection legislation.^5, 6^ However, even supposedly anonymised data relating to individual people carries some risk of re-identification via “jigsaw” attacks.^10^

**Figure 1.**
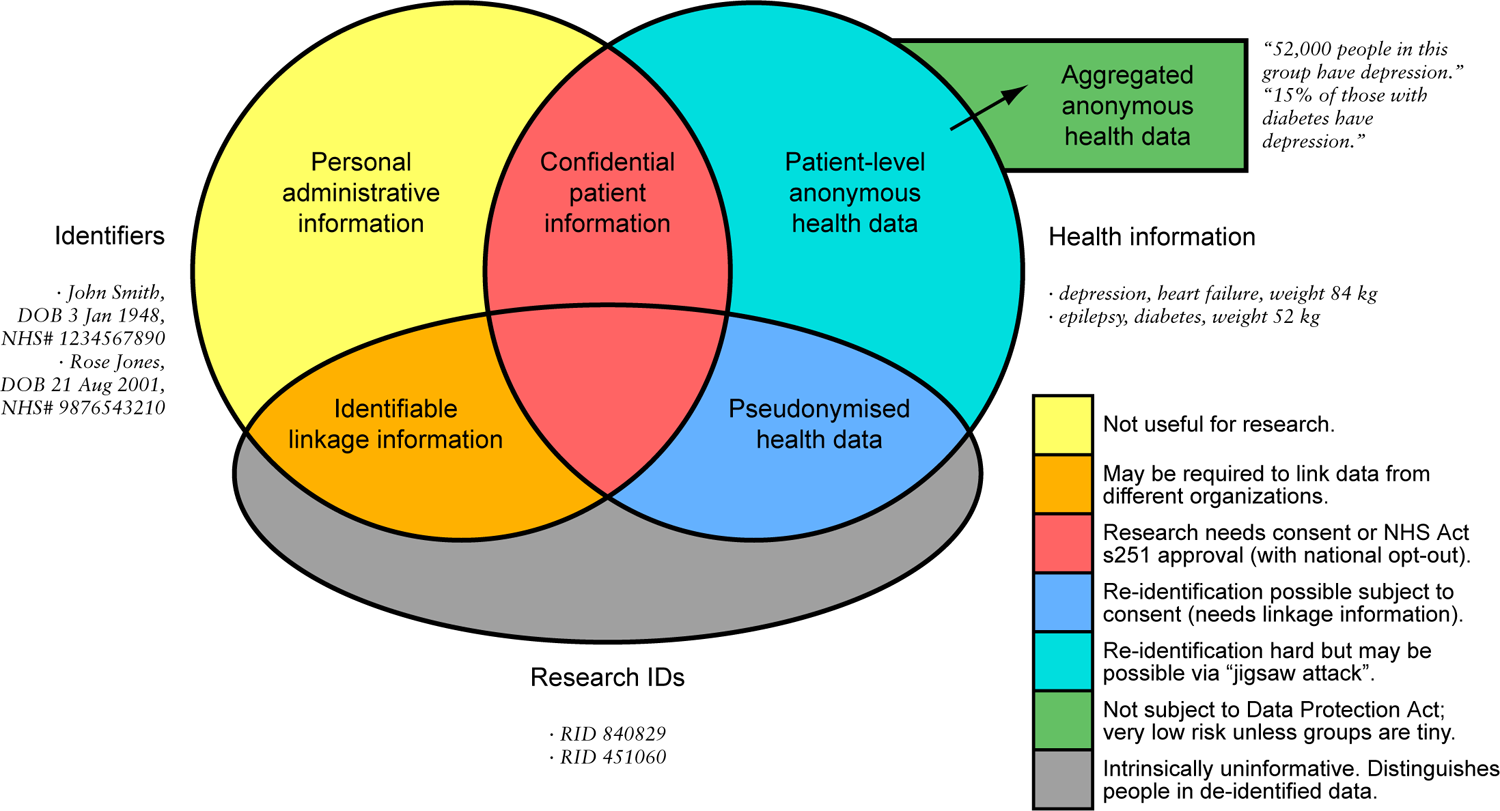
Classifying health data according to identifiability. At the “patient” level, the Venn diagram shows the overlaps between information that directly identifies a person, research identifiers (RIDs or pseudonyms), and health information, with simple examples. Anonymous health data may also existing in aggregated form, distinct from patient-level data; this aggregated form is the norm for public distribution. The level of identification risk and the research information governance requirements vary with the resulting categories of data, shown in the legend. All examples are fictional. (DOB, date of birth.)

Our understanding of public wishes about data sharing is incomplete.^11^ Information is sometimes not shared clinically when it should be,^3, 12^ and patients may be surprised and frustrated by failures to share in a “national” health service.^13, 14^ Previous work, whilst establishing themes in public views on data linkage and sharing for research,^11, 15–21^ has highlighted the very small scale of many studies, and the paucity of research about the views of minority groups and the acceptability of sharing some types of data, such as mental health (MH) data. Mental illnesses can carry significant stigma^22^ but are associated with substantial loss of life expectancy,^23^ necessitating improvements in research and care. Some research requires multi-source data, but linkage is complex and may involve transient use of identifiable information.^24^ It is unclear to what extent the public supports such work, and whether support varies with the type of data to which health data is linked (e.g. education versus criminal justice); there is little prior research in this area. ^18^ Proposed national systems for NHS data research such as “care.data” have previously aroused public ire,^25^ as have information governance (IG) breaches,^26^ and there is current debate about the newest NHS data sharing proposal, General Practice Data for Planning and Research (GPDPR).^8, 27^

What do patients and the public want now? We studied views on the sharing of identifiable health data (for clinical purposes) and de-identified health data (for research) within the UK. We examined data destinations ranging from local NHS services to public distribution. We distinguished MH and physical health (PH) data, and for research also structured versus free-text (narrative) data. We asked about data linkage for research. We superimposed a randomized experiment to quantify how opinions on sharing were affected by the “framing”^28^ of risk versus benefit. We examined the effect of the COVID-19 pandemic upon preferences. We sought views on potential systems to govern NHS data sharing for clinical and research purposes and to offer direct participation in research.

## 3. Methods

### 3.1. Patient and public involvement

The research team advertised and formed a research advisory group (RAG) comprising patients and carers, who designed the study with the research team. Patients, carers, and other members of the public participated in the study.

### 3.2. Approvals

We obtained NHS ethics approvals (East of Scotland Research Ethics Service, reference 19/ES/0144) and pre-registered the study (https://doi.org/10.1186/ISRCTN37444142).

### 3.3. Inclusion and exclusion criteria; sample size

The inclusion criteria were current residence in the UK and informed consent. The ability to take an online survey (alone or supported) was implicit. Participants under 16 required the permission of their parent or guardian to participate and were asked to report whether they had assistance. We sought a power of 0·9 to detect a “small” effect (Cohen’s *d*=0·1) for the framing intervention (described below), with an estimated minimum *n*=433/group, but beyond that sought a large representative sample of the UK population.

### 3.4. Recruitment

Approvals covered public announcements and recruitment via health service sites, in person or through a variety of media. The study was adopted onto the National Institute for Health Research (NIHR) Clinical Research Network (CRN) portfolio; 216 general practice (GP) surgeries and 154 large healthcare organizations (e.g. acute care Trusts, MH Trusts, community hospitals, ambulance Trusts) supported recruitment. The study ran from 2020-02-07 to 2020-09-30.

### 3.5. Survey

Data were collected using REDCap.^29^ The survey is reproduced in the **Supplementary Methods**. It asked for the respondent’s views on current and desirable practice for sharing identifiable data for clinical care purposes; personal experience of MH/PH conditions and care; preference for sharing identifiable PH/MH data (for clinical care purposes) to a range of NHS “destinations”; preference for sharing de-identified structured PH/MH data (for research) to a range of potential research “destinations”; similarly for data including de-identified free-text notes; views about potential systems for managing data consent in the NHS; views about linkage for research to non-NHS data sources; and demographics.

### 3.6. Randomized framing intervention

We hypothesized that the context of questioning would affect willingness to share MH versus PH data, and sought to control and measure this effect. Before we asked about willingness to share different kinds of health data, we presented one of three framing statements: neutral, “concern” (about MH data being more sensitive), or “holistic” (about the importance of joined-up PH/MH care) (**Supplementary Methods**). Participants were randomized to one of the three statements.

### 3.7. Data processing

Where participants agreed to leave a postcode, this was converted to a larger Office for National Statistics (ONS) geographical area, to prevent inadvertent identification. The geographical area was linked to its known population and Index of Multiple Deprivation (IMD). If the participant provided sufficient information, the ONS National Statistics Socio-Economic Classification (NS-SEC) was also calculated. (See **Supplementary Methods**.)

After removal of all free text, anonymised data are available from **[URL_to_be_established_within_**https://www.data.cam.ac.uk/repository], with participants’ consent.^30^

### 3.8. Pandemic

By chance, our study spanned the UK onset of the COVID-19 pandemic. This had many consequences, including “lockdowns”. Major changes were made to NHS data handling, including instructions to share CPI for public health purposes relating to the pandemic,^31^ media reports of sharing of patient-level de-identified data with industry,^32^ and guidance for GPs to include additional information in patients’ Summary Care Record (SCR, England) unless they had previously opted out.^33^ We examined whether the pandemic was associated with changes in preference relating to data sharing, using 2020-03-23 (first UK “lockdown”) as the split point (factor “pandemic”: levels “before lockdown”, “at/after lockdown”).

### 3.9. Analysis

We analysed using R v3.6.3.^34^ We analysed categorical associations via χ^2^ tests, and effects upon ordinal Likert-type scales (phrased linguistically to approximate interval scales) via analysis of variance (ANOVA). With a large sample size, the central limit theorem means that the distribution of means and mean differences tends to normal even though the parent population is non-normal, and ANOVA is robust to non-normality,^35–37^ permitting ANOVA of discrete dependent variables. Scales measuring likelihood were quantified as −2 very unlikely, −1 unlikely, 0 not sure, +1 likely, +2 very likely. Yes/no scales were quantified as −1 no, 0 not sure, +1 yes. Models involving within-subjects terms were analysed using the *lmer* and *lmerTest* packages, using type III sums of squares, and are expressed thus (∼ “is predicted by”; A×B interaction; A*B denotes the inclusion of main effects A and B and their interactions). Statistics are shown to 3 significant figures and degrees of freedom are rounded to integers. We set *α*=0·05, and report “NS” for “not significant” and “VLP” for a very low *p* value, *p*<2·2×10^−16^.

Opinions on sharing clinical/identifiable data were analysed using a model termed C1: *sharing∼destination*nature*framing*pandemic+(1|subject)*. “Destination” had four levels (local, regional, national, UK-wide), “nature” had two (PH, MH), and framing had three (neutral, MH concern, holistic). We followed up nature×framing interactions by analysing MH and PH data separately using the simplified model C1B: *sharing∼destination*framing+(1|subject)*.

To examine the effects of demographic factors and experience, we used a larger model, C2: *sharing∼destination*nature*framing*pandemic+age+gender+ethnicity+education+sexuality+religion+nat ion+imd_quartile+nssec+mh_experience*nature+(1|subject)*. This was only possible for people who provided all necessary demographic information. Levels for demographic factors were as per **Supplementary Table 1**, plus sexuality (two levels: heterosexual/straight, LGBT+ [including homosexual/gay/lesbian, bisexual, other/self-described]) and NS-SEC (five levels).

Opinions on sharing de-identified data for research were analysed using model R1: *sharing∼destination*nature*detail*framing*pandemic+(1|subject)*. “Destination” had six levels (NHS, academia, charities, companies conducting treatment research, other companies, publicly); “detail” had two levels (structured only, free text); other factors were as before. To examine nature×framing interactions, we used the simplified model R1B: *sharing∼destination*framing+(1|subject)*. For demographic analysis we used model R2: *sharing∼destination*nature*detail*framing*pandemic+age+gender+ethnicity+education+sexuality+religion+nation+imd_quartile+nssec+mh_experience*nature+(1|subject)*.

Sensitivity analyses were conducted by weighting to UK population demographic proportions. Effect size plots were created for key models. (See **Supplementary Methods**.)

Willingness for linkage to non-NHS data for research (data source, eight levels) was analysed for all participants using model L1: *willingness∼source*pandemic+(1|subject)*. For demographic analysis we used model L2: *willingness∼source*pandemic+age+gender+ethnicity+education+sexuality+religion+nation+imd_quartile+nssec+mh_experience+(1|subject)*.

A thematic analysis was performed on free-text comments (see **Supplementary Methods**).

## 4. Results

### 4.1. Participants

Consenting participants numbered 29275. Recruitment is shown in **Supplementary Figure 1A–B**; 8019 participated before UK “lockdown” and 21256 on/after that date. Not everyone completed the survey: participation by stage is shown in **Suppl. Fig.1C**. Median completion time was 18·4 minutes. Participants were evenly distributed across framing conditions (neutral 9812, MH concern 9744, holistic 9719; χ^2^ =0.475, NS).

Demographics are shown in **Suppl. Fig.2** (with free-text responses in **Supplementary Results**). Relative to the UK population (**Supplementary Table 1**), our sample under-represented the youngest and oldest age ranges, males, those of non-white ethnicity, those with less formal education, those professing a religion, residents of UK nations other than England, and people living in more deprived areas. Weighting yielded substantial though incomplete improvement. There was coverage of most UK local authority areas (**Suppl. Fig.2I**).

**Figure 2.**
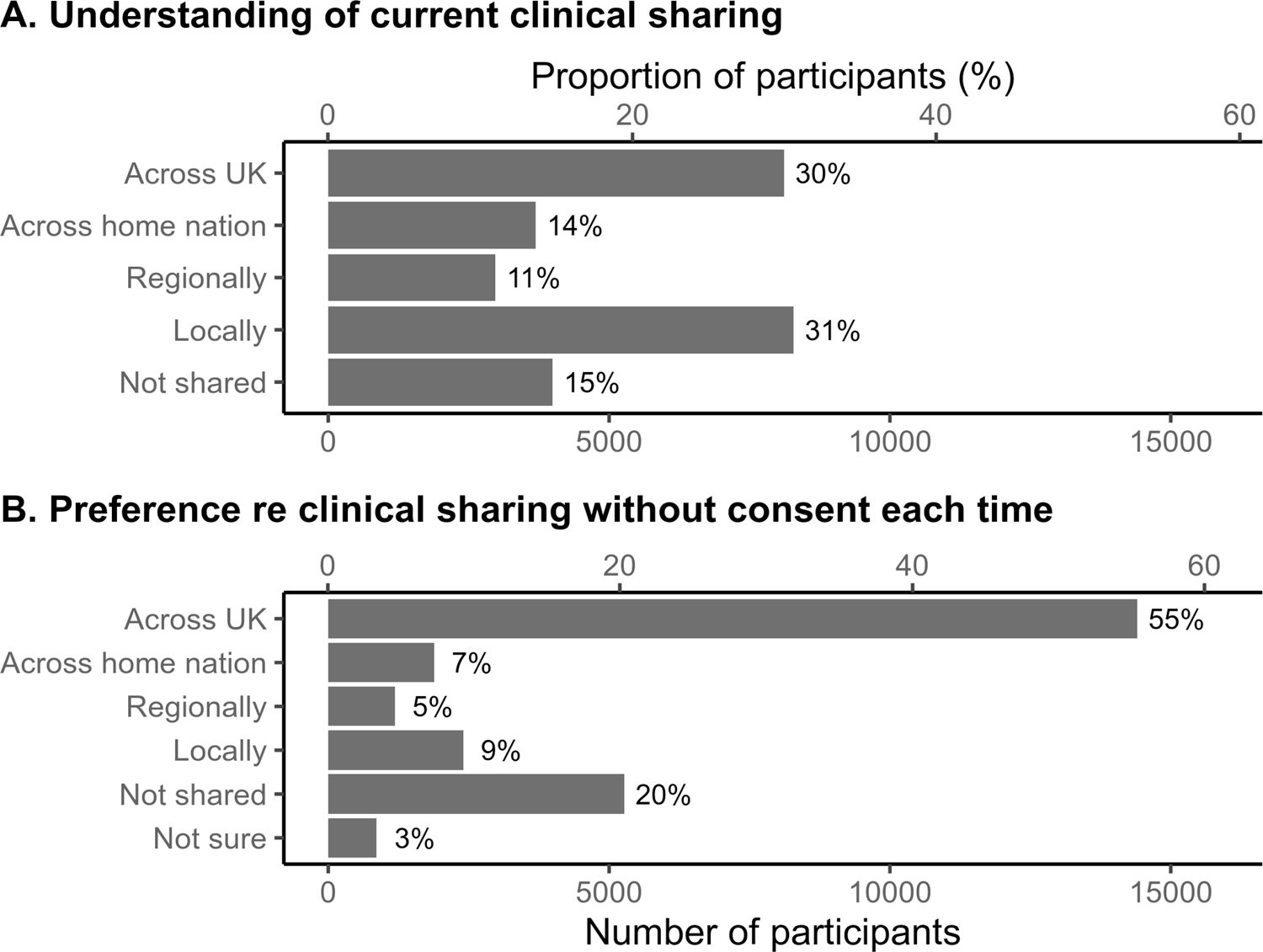
(A) Understanding of how health data are shared identifiably without explicit consent for clinical purposes, and **(B)** preference as to what should happen. The denominator for percentages is the number of people who answered each question.

Forty percent of participants had experienced a MH condition (**Suppl. Fig.3**), primarily depression and anxiety disorders. Of them, 85% had used MH services, primarily their GP and NHS psychological therapy services. Eighty-eight percent of respondents had used PH services, primarily GP and outpatient services.

**Figure 3.**
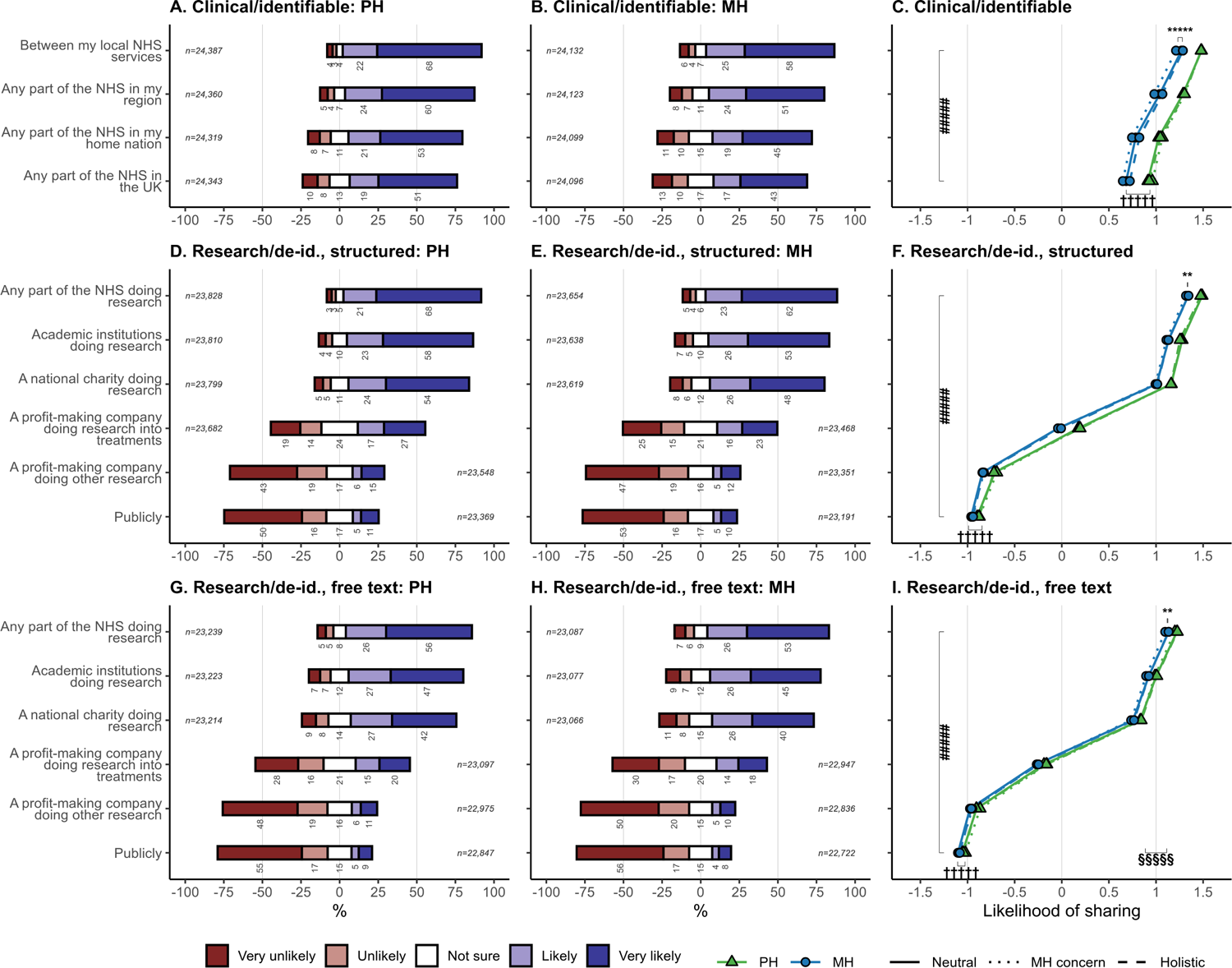
Participants’ self-reported likelihood of sharing mental and physical health data without explicit consent for clinical purposes (identifiably) or research (de-identified), according to destination, nature (MH versus PH), framing statement, and for research purposes also the level of detail (structured only versus with free text). The denominator for percentages is the number of people who answered each question. In plots C, F, and H, the abscissa is the mean of responses coded as –2 very unlikely, –1 unlikely, 0 not sure, +1 likely, +2 very likely. Analyses were from models C1 and R1 as described in the Methods. (De-id., de-identified; MH, mental health; PH, physical health; #####, *p*<10^−5^ for main effect of destination; †††††, *p*<10^−5^ for main effect of nature, with bar length showing mean difference between MH and PH; ** *p*<0.01 and ***** *p*<10^−5^ for framing×nature interaction, with bar showing the mean difference between “MH concern” and “neutral”; §§§§§, *p*<10^−5^, main effect of detail, comparing panel F with panel I, with bar length showing the mean difference between structured and free-text conditions.) See **Supplementary Figure 4** for corresponding weighted analysis.

### 4.2. Sharing identifiable data for clinical purposes

Understanding of current NHS practice regarding identifiable data sharing between care providers, without asking the patient each time, are shown in **Fig.2A**. In practice, sharing varies by area, e.g. depending on whether a local/regional shared care record is operative in part of England,^14^ or according to limited national systems such as the Intra-NHS Scotland Information Sharing Accord^38^ and Scottish Emergency Care Summary,^39^ the Northern Ireland Electronic Care Record,^40^ the English SCR,^41^ and a variety of systems in Wales.^42^ To our knowledge, there is no UK-wide sharing, but 30% of respondents thought that there was free sharing of identifiable data across the UK.

When asked preferences via a single multiple-choice question (**Fig.2B**), there was majority (55%) support for sharing identifiable data for direct care across the UK, without being asked first, and 76% support for sharing at least locally, but a substantial minority (20%) said that sharing should not occur without the patient being asked first.

### 4.3. Sharing mental and physical health data for clinical purposes and for research

Willingness to share health data without being asked every time is shown in **Fig.3** by purpose, type, and destination.

For clinical purposes (with identifiable data), there was strong net willingness to share (**Fig.3**). The most important determinant was destination, with stronger support the more local the sharing. People were slightly more willing to share PH than MH data. There were significant but very small effects of the framing statement, primarily that “MH concern” framing reduced willingness to share MH data. In the whole-sample analysis (model C1), there were highly significant effects of destination and nature, as well as interactions including nature×framing (**Fig.3**, **Suppl. Fig.6A**). This interaction was driven primarily by a simple effect of “MH concern” framing to reduce sharing for MH data [model C1B: PH data, no effect of framing (*F*_2,24461_=1·18, NS); MH data, effect of framing (*F*_2,24157_=8·36, *p*=0·000234); pairwise comparison within MH data, MH concern versus neutral, *p*=0·00443]. Framing effects were also lessened for geographically broader destinations.

For research purposes (with de-identified data), destination was an extremely strong driver of preference (**Fig.3**). On average, people expressed strong support for sharing to the NHS, academia, or national charities for research purposes. Support and opposition were approximately equally balanced for sharing to profit-making companies researching treatments. There was strong net opposition to sharing to other types of companies, approximately equal to that for sharing publicly. There was a small but significant preference for sharing PH (versus MH) data, and likewise higher preference for sharing structured-only versus free-text data. In the whole-sample analysis (model R1), there were highly significant effects of destination, nature, and detail, plus interactions including destination×nature×detail (**Suppl. Fig.6B**). Framing effects included nature×framing, though simple framing effects were not significant for PH or MH data separately (model R1B).

Sensitivity analyses weighted to UK population demographics (**Supplementary Results**) were consistent with the primary analysis.

### 4.4. Linkage to non-health data for research

We asked about linking of NHS data to non-health data sources for research. There was net support for all “state” sources and university-held data (**Fig.4**), but net opposition regarding private company data (**Fig.4**). Weighted responses were very similar (**Suppl. Fig.5**).

**Figure 4.**
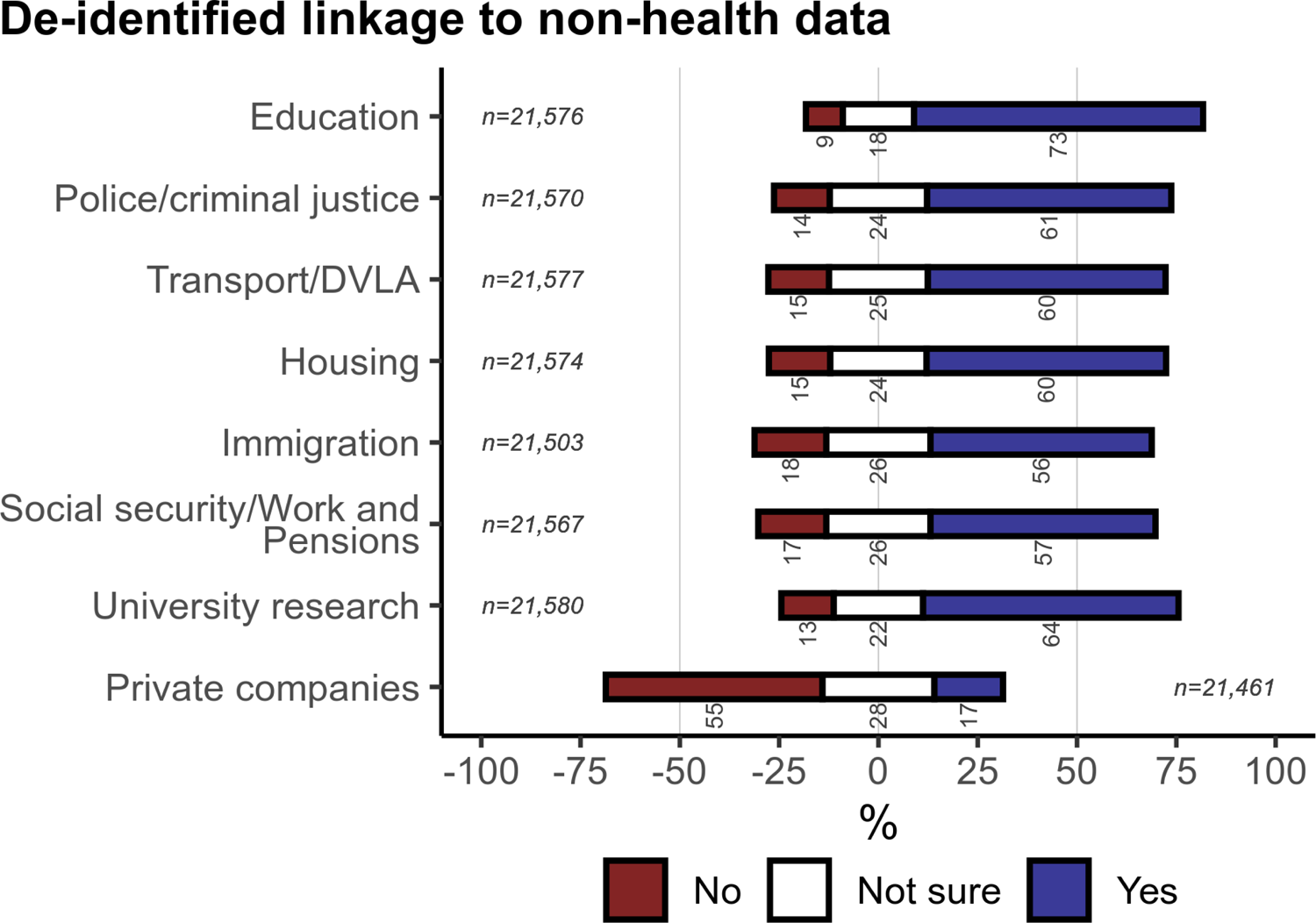
Participants’ willingness for their NHS data to be linked to non-health data of different kinds for research. The denominator for percentages is the number of people who answered each question. See **Supplementary Figure 5** for corresponding weighted analysis.

**Figure 5.**
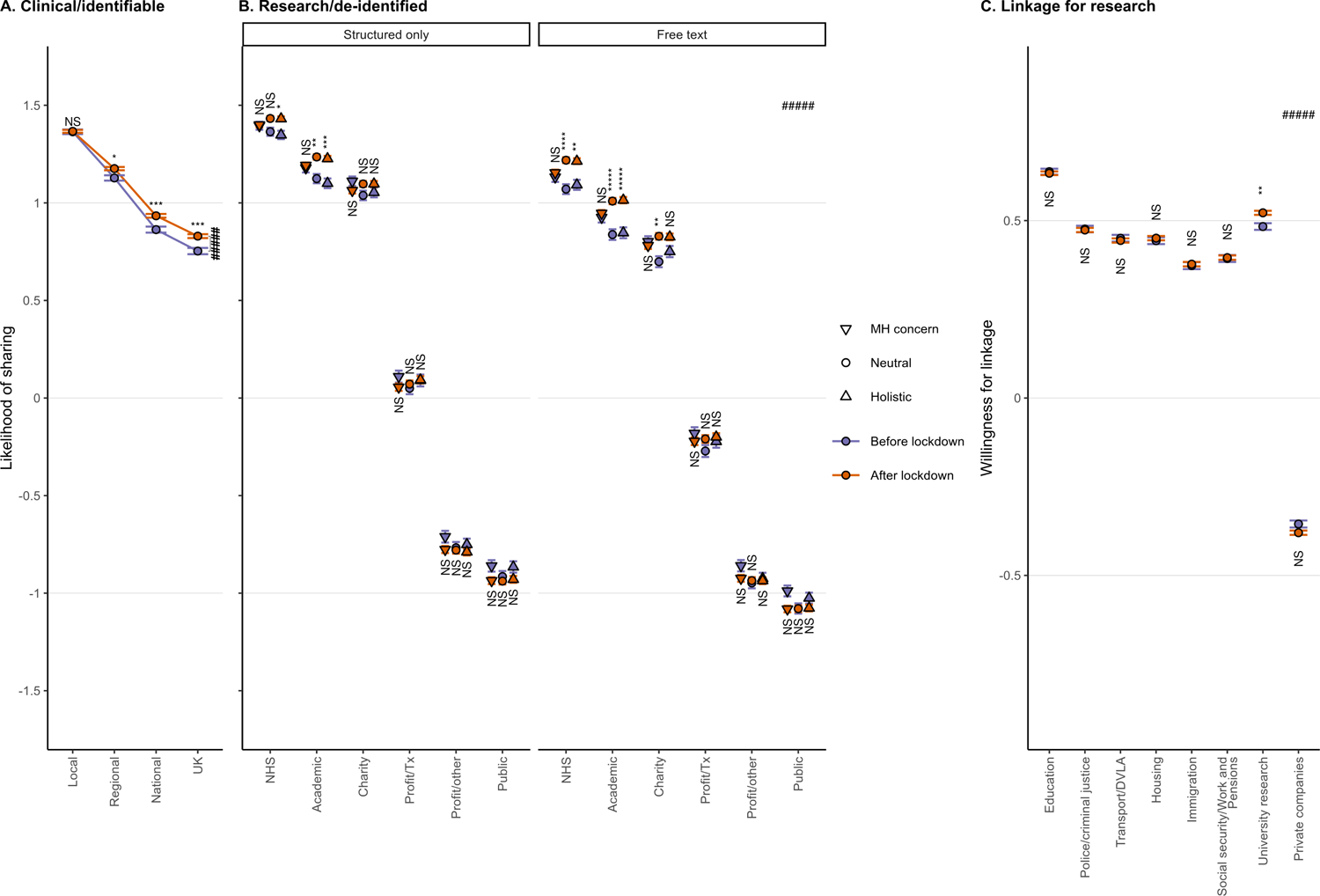
Change in preference in relation to the COVID-19 pandemic. **(A)** Clinical/identifiable data sharing, by destination. Dependent variable (preference for sharing) as for Figure 3C, now shown on the ordinate (*y*) axis. **(B)** Research/de-identified data sharing, by detail (structured versus free text) and destination. Dependent variable as for Figure 3F,H. **(C)** Linkage for research, by non-NHS data source type. Dependent variable coded as –1 no, 0 not sure, +1 yes. (Error bars show ±1 SEM; ***** *p*<10^−5^, **** *p*<10^−4^, *** *p*<10^−3^, ** *p*<10^−2^, * *p*<0.05 by two-sample *t* test Šidák-corrected for multiple comparisons; NS, not significant; ##### *p*⋘10^−5^, destination×pandemic interaction.)

### 4.5. Changes related to the COVID-19 pandemic

Following “lockdown”, willingness to share identifiable data for clinical purposes increased, with no significant change in the already high preference for local sharing, but progressive increases for sharing to more remote parts of the NHS (model C1, destination×pandemic, *F*_3,169348_=26·6, VLP; **Fig.5A**).

Willingness to share de-identified data for research purposes generally increased for more-preferred destinations (NHS, academia, charities), except in the “MH concern” framing condition (model R1, destination×pandemic, *F*_5,535334_=78·2, VLP; **Fig.5B**), but did not change for less-preferred destinations (commercial and public sharing).

Preference for linkage to university data increased (source×pandemic; **Fig.5C**; Suppl. Fig.6C). There was a less consistent decrease in preference for linkage to private data ( **Fig.6C**, Suppl. Fig.6C) and police data (model L2; **Fig.6C**).

### 4.6. Effect sizes and influence of demographic factors

Preference varied according to demographic factors and experience of MH illness. For clinical purposes, there were several demographic effects (model C2, **Fig.6A**). Age was a significant factor, with the age bands most willing to share being 25–44 and 75+, and the 18–24 band being least willing. Males were more willing to share data than females. Those of minority ethnicity were less willing to share than those of white ethnicity. Across educational levels, those of Level 3 were most willing and those of Level 4+ least willing. Those of minority religions were less willing to share. Those from the most-deprived IMD quartile were also less willing. There were no significant effects of sexuality (*F*_1,12334_=2·21, NS), NS-SEC (*F*_4,12335_=1·32, NS), or nation (*F*_3,12335_=2·13, NS). Personal experience of MH illness specifically reduced willingness to share MH data for clinical purposes (nature×MH experience).

For research purposes, significant effects were similarly observed for age, gender, ethnicity, religion, and IMD quartile (model R2, **Fig.6B**). The age distribution was clearly U-shaped, with greater willingness to share among the youngest and oldest groups. As before, there was no effect of sexuality ( *F*_1,12336_=1·58, NS). There was no effect of education (*F*_4,12348_=2·15, *p*=0·072), but there was an effect of nation (with people living in Wales more willing to share and those in Scotland less so, relative to England), and of socioeconomic status (NS-SEC; **Fig.6B**). People with MH experience were significantly *more* likely to share MH data for research purposes (nature×MH experience, *F*_1,296111_=6·15, *p*=0·0132).

For linkage, the patterns were broadly as before (**Fig.6C**, **Suppl. Fig.6C**). Data source strongly influenced preference (education>universities≈police≈housing≈transport>social security>immigration≫private companies). There were also effects as before of age, ethnicity, education, and IMD quartile. There was no effect of gender (*F*_1,12338_=2·32, NS), religion (*F*_2,12338_=1·86, NS), nation (*F*<1), or MH experience (*F*<1), but there was now an effect of sexuality, with LGBT+ people being less willing for linkage.

**Figure 6.**
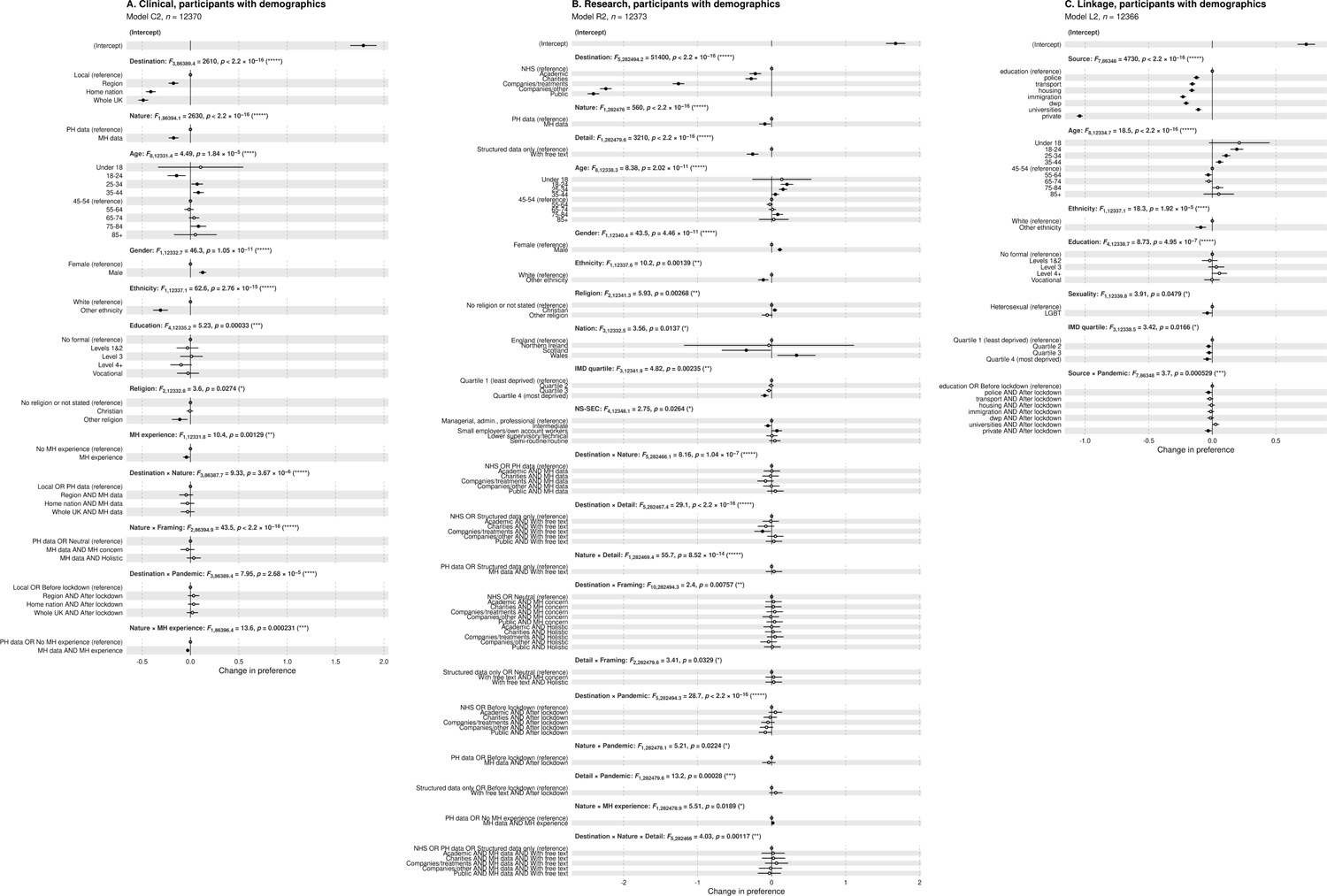
Effect sizes for **(A)** clinical data sharing via statistical model C2, **(B)** research data sharing via model R2, and **(C)** linkage via model L2. These models include only those participants who supplied full demographic information, to allow analysis by demographics; compare **Supplementary Figure 6** (all participants). Only those model terms with a significant *F* test are shown. Effect sizes with 95% confidence intervals are shown for each level as uncorrected pairwise comparisons to a reference category within each term (note the difference in what is being tested pairwise versus the omnibus *F* test for the term; see **Supplementary Methods**). ● *p*<*α*; ○ NS.

### 4.7. A possible national consent system

We proposed varieties of a national system for patients to decide how their NHS data is used. Participants were most willing to sign up and change their preferences via a web site or in person ( **Fig.7A,C**). Willingness was similar regardless of whether consent information was managed by the NHS centrally, a local NHS Trust, or the patient’s GP (**Fig.7B**). Overall, 89% of people said they were “likely” or “very likely” to sign up to such a system (**Fig.7D**). Weighted responses were very similar (**Suppl. Fig.7**). Most people wanted a single NHS system to sign up for participatory research (**Fig.7E; Suppl. Fig.7E**).

**Figure 7.**
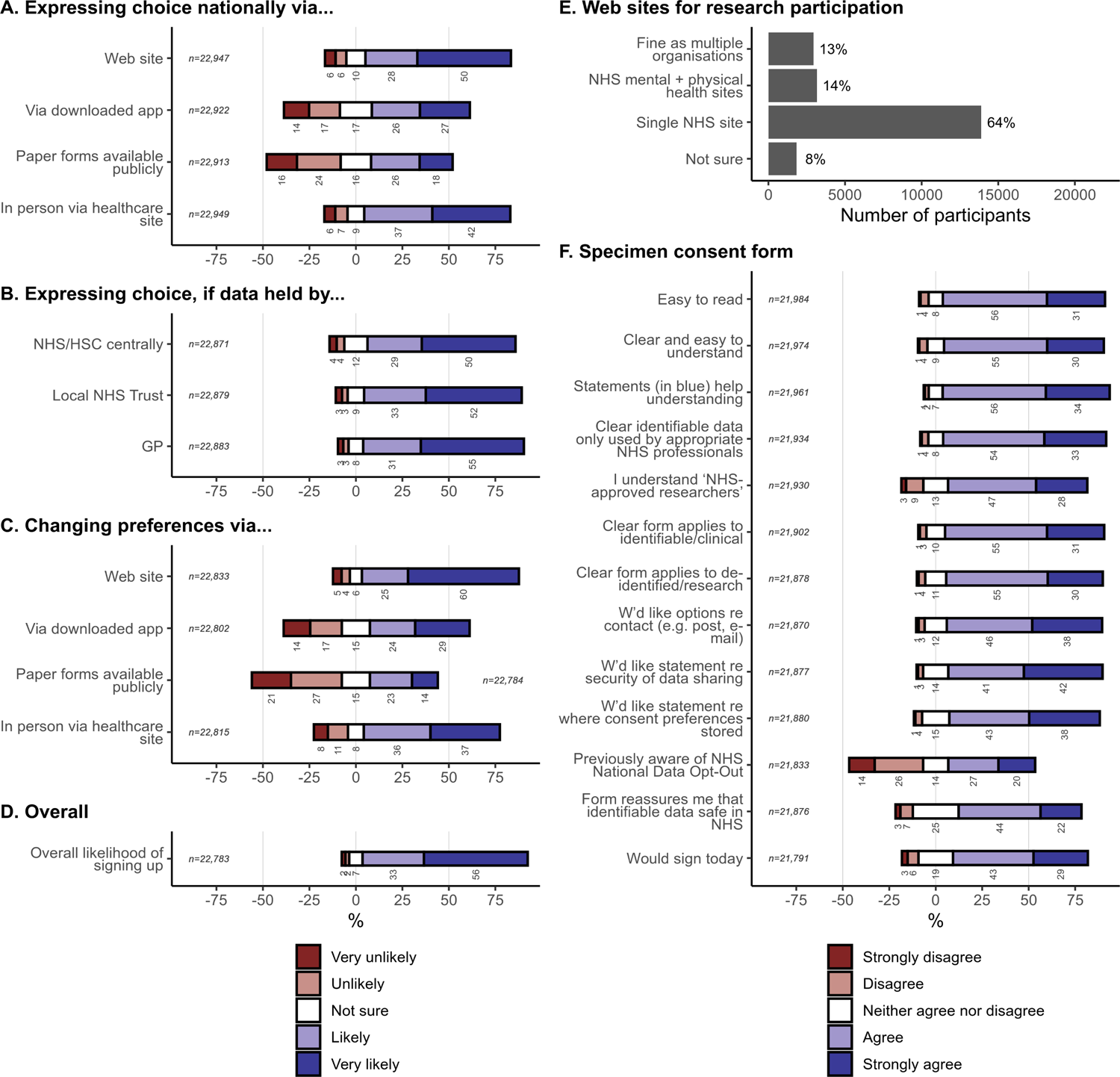
Views on a national data sharing consent system. The denominator for percentages is the number of people who answered each question. See **Supplementary Figure 7** for corresponding weighted analysis.

There was broad support for the draft consent form and for adding information about contact methods, data security, and management of the consent information (**Fig.7F**; **Suppl. Fig.7F**). Forty percent (unweighted) had been unaware of the NHS National Data Opt-Out.

Comment themes (*n*>100, **Supplementary Results**) included: the need for clarity around de-identification; the critical importance of healthcare data security; the desirability of data sharing; that opt-outs should be more prominent (or default) or linked to the NHS National Data Opt-Out; that profit-making use should not happen or that the NHS/patients should benefit from such profits; that clinical users should be specified in more detail; research users likewise; and that healthcare data should not be available to private or third-party companies without specific permission.

## 5. Discussion

### 5.1. Summary

Many respondents believed that health data is shared UK-wide for clinical purposes without explicit consent, when sharing is usually more limited. A majority supported such sharing, though a significant minority opposed it. Geographically broad sharing was endorsed, though with stronger support for more local destinations. People preferred to share PH (versus MH) data but this was less important than the destination.

For research, with de-identified data, there was strong net support for sharing without explicit consent to the NHS, academic research institutions, and research charities. There was net ambivalence regarding private companies researching treatments, and strong net opposition for sharing to other companies or publicly. There was a small preference for sharing PH over MH data (a smaller difference than for clinical purposes), and greater support for structured-only data over de-identified free text. There was net support for research linkage to state and university data sources, but opposition regarding data held by private companies.

Framing statements influenced MH/PH preferences, but only to a small degree. Age, gender, ethnicity, education, religion, and IMD were associated with willingness to have health data shared or linked, though not nearly as strongly as destination/source. Personal experience of MH conditions was associated with reduced willingness to share MH data for clinical purposes, but greater willingness to share it for research. After COVID-19 lockdown there was greater willingness regarding already-preferred destinations.

Respondents endorsed a suggested UK-wide system allowing patients to control the clinical/research uses of their data and to sign up for participatory research. They frequently emphasized the importance of data security and that NHS data should not be made available to private or third-party companies without specific permission.

### 5.2. Strengths and weaknesses

Strengths include patient/public involvement in the study design; the largest such study to date by 1–3 orders of magnitude,^15, 17, 19–21, 43^ giving high power; sensitivity analyses weighted to population demographics; detailed questions about data sharing for clinical/research purposes, including about the type of data and the destination, plus linkage to non-health data; a randomized framing experiment to control and measure this source of variation; quantitative analysis including of relative effect sizes; consultation on ways to improve the current situation; and serendipitous examination of the effects of COVID-19 on data sharing views.

The major weakness is that the sample remained under-representative of some groups despite weighting, with potential for unmeasured selection (including self-selection) bias, reducing generalisability.

### 5.3. Destination and purpose

The Caldicott framework (2013) and review (2016), regarding safe information sharing for direct clinical care, included the principle that the “duty to share information can be as important as the duty to protect patient confidentiality”^3^ and noted that information was often not shared when it should be, for fear of inappropriate disclosure.^12^ That was despite legislation creating a duty upon providers to share information with professionals when that is likely to facilitate the individual’s health or social care, disclosure is in their best interests, and they do not or are not likely to object.^44^ That is in essence an opt-out system. This legislation conflicted with some prior studies of public opinion.^45^ The review noted low public understanding around how health information is used, but “an expectation that information is shared for direct care”.^12^ We observed net support for such sharing that varied with geographical destination and was by no means universal, but was nevertheless strong.

In relation to research and other non-clinical activities, the recommendation that people be able to opt out from personal confidential data being used beyond their own direct care^12^ led to the NHS National Data Opt-Out.^46, 47^ This relates to the use of CPI (identifiable information) for purposes such as research, conducted under NHS Act Section 251 (s251) approvals.^1, 48^ It does not apply to direct clinical care, local audit or service evaluation, or de-identified information.^46, 47^ Our study and others show it remains unknown to many.^43^ Furthermore, it is not simple and we suspect many do not fully understand its scope. Conversely, from the researcher’s perspective, s251 approval is often still required for linkage studies in which researchers never see identifiable information: there is no standardized “trusted third party” system for centralized linkage of identifiable information, and inconsistent adoption of de-identified linkage methods.

“Destination” was by far the strongest driver of preference for sharing and linkage. This pattern is established: willingness to allow researchers/clinicians access to health data, but far greater reservations about industry.^49, 50^ An important basis for this is mistrust of the security and/or motives of commercial organizations,^25, 26, 51^ as our participants noted.

### 5.4. Demographic effects

A common demographic theme was that minority groups (of ethnicity, religion, and sexuality) and deprived groups were less willing to share. This might reflect experience of disadvantage to, or discrimination against, these groups.^52, 53^ Ethnicity has had mixed effects on preference for national electronic health record (EHR) systems.^19^ In our study, age effects were generally biphasic, with higher willingness amount the youngest and eldest. Youth may be associated with familiarity with data and/or greater support for EHRs,^54^ and older age with an increasing burden of illness, itself associated with support for national EHR systems. ^54^ Educational effects were relatively inconsistent. Males were slightly more willing to share than females. Similar results have been observed before,^43^ but not always;^54, 55^ one reason might be gender-based healthcare discrimination.^56^ Higher support for research sharing in Wales may relate to established national research systems there;^57, 58^ the reasons for reduced willingness in Scotland are unclear, but similar systems there are younger.^59^ Those with personal experience of MH illness were less willing to share identifiable MH data for direct care. This may reflect experience of discrimination or stigma^60, 61^—which can have disproportionate effects in subgroups.^61, 62^ However, the same people were more willing to share de-identified MH data for research, potentially reflecting increased prioritization of MH research.^63^

Demographic variations in preference may reflect differences in perception of current data rules or security practices, reasons for concern about uses of health data, or degree of concern. UK law prohibits variation of policy according to these factors.^64^ Better understanding and public information may be required to address these groups’ concerns,^11, 65^ but improvements in health equity are also required.^17, 52^ However, the effect sizes of these demographic predictors were not large enough to override the net support for data sharing, given the right destinations.

### 5.5. Framing and pandemic effects

We observed small but significant framing effects.^28^ Our framing statements were true and non-alarmist, so real-world framing effects might sometimes be larger. Others have observed larger effects via “loss framing” (emphasizing the potential adverse consequences of not consenting over the potential benefits of consenting), and through other manipulations like the placement of framing statements.^66^ Media coverage of health data sharing is influential.^67, 68^ Despite best intentions it is impossible to avoid framing effects entirely,^28^ so those presenting information should be aware of these whilst presenting accurately the risks and benefits of data sharing/linkage.

During COVID-19, despite press coverage^32^ of an enforced increase in sharing,^31, 33^ support for sharing/linkage increased—but only for some already-favoured destinations. Publicity regarding NHS care ^69^ and research regarding COVID-19^70, 71^ may have driven the increase in support for sharing with the NHS, universities, and research charities.

## 5.6. Conclusions

Participants supported a central system for patients to control the uses of their data, and likewise a single NHS mechanism to sign up for active research participation. There is a trade-off between the scientific desirability of everyone contributing de-identified data, including to avoid bias, ^72, 73^ and the desirability of individual control over data use.^74^ A reasonable balance might be a central system to opt out from identifiable clinical use, identifiable (s251) research use, or de-identified research use of one’s data, and to opt in for participatory research. This would complement efforts to improve people’s access to their own data.^75^

The majority support observed for clinical sharing without explicit consent perhaps makes such sharing reasonable as a default (opt-out) position, given the potential advantages for many people’s own care, subject to strict IG rules (who has access, when). However, a significant minority opposed this, mandating at least a public information campaign about opt-outs if this were to occur.

There was strong net support for NHS, academic, and charity researchers accessing de-identified health data. A standard method is a trusted research environment (TRE).^24, 57, 76, 77^ Approved researchers come “into” the secure environment to interact with relevant data (e.g. pseudonymised; **Fig.1**). After analysis, aggregation, and other statistical disclosure control (SDC),^78^ results go “out” for publication (**Fig.1**).

In contrast, respondents did not support research sharing to private companies. Some have suggested this is addressable in part by public education.^11^ We suggest respecting public preference, and not giving commercial organisations direct access to patient-level NHS data for research, even de-identified, without consent. (This is distinct from the common NHS practice of employing companies, such as EHR software providers, to manage NHS data securely for clinical purposes.) We think that this does not rule out all industrial research uses of data, which could happen according to at least three methods. The first is via consent, as for commercial treatment trials. Secondly, companies could collaborate with NHS/academic researchers. For example, an artificial intelligence company could provide an untrained algorithm; NHS staff could train it on patient-level data; the company could receive a trained algorithm back whilst never having access to the data (assuming verification that the algorithm cannot “embed” detailed data features during training). Thirdly, methods exist whereby software queries come “in” to the TRE, and semi-automatic or automatic SDC occurs before results go “out”.^79–81^ This allows research to take place without researchers having access to patient-level data, and can also support “federated” queries across sites. Data that have undergone suitable SDC (e.g. aggregation) can be published, and are therefore suitable for industrial access if desired. Regardless, as our participants commented, the NHS might charge for such access,^11, 82^ and full transparency is essential. Formal, consultation-based standards governing this NHS–commercial interface would be desirable.

Governance of UK health data must be transparent and reflect the views of patients.^11, 16^ As the UK Government seeks to change data legislation^83^ and emphasize health data in its science strategy,^84^ we hope this study contributes to the conversation.

## Data Availability

Full anonymous data set to be made available (with participant consent) from the University of Cambridge Data Repository -- URL to be established.

## 6. Acknowledgements

We thank all those who took part in the study and gave their views. We thank all the investigators, local collaborators, local NIHR CRN staff, and health care staff across the UK who supported the study. We thank also Elizabeth Rotherham (research advisory group member) for help with design; Nicola Gleadle for practical support; Jane Gaffa, Rachel Kyd, and Mary-Beth Sherwood for Research and Development support; Sally-Anne Hurford and Ruth Hudson for NIHR CRN advice; Chess Denman for investigator approval; Laura Marshall for CPFT publicity; Hannah Clarke for helpful comments; REC and HRA staff for ethical and regulatory approvals; the Cambridge Autism Research Database for additional publicity.

## 7. Author contributions

RNC conceived the study. LAJ, JRN, PHA, MRG, MP and RNC designed it (with ER). LAJ coordinated its execution. JMF advised on REDCap implementation and execution. RNC, LAJ, and JMF had access to the original data; the final data set is public. RNC analysed the data. RNC and LAJ drafted the manuscript. All authors edited the manuscript and approved submission.

## SUPPLEMENTARY MATERIALS

### S1. Supplementary Methods

#### S1.1. Survey design

The survey sequence is shown in full in the **Appendices.** In outline, it was as follows:

- Information, consent, and a question how the participant heard about the survey. (Participants were asked to indicate the health organization, if any, through which they had heard about the study and recruitment was attributed to those sites.)
- An explanation of “NHS” (used as shorthand for NHS/HSC), “health data”, “identifiable health data”, and “clinical care”.
  - Questions about **current and desired sharing of identifiable health data for clinical care purposes.**
  - A multiple-choice question on the respondent’s understanding of how NHS clinical care providers
  - *currently* share their identifiable data for clinical care purposes, without asking each time.
  - A statement that most NHS providers are separate and do not currently share identifiable data without asking, followed by a multiple-choice question as to if/how the NHS *should* share the respondent’s identifiable data for clinical care purposes, without asking each time (with all stems ending “… without asking me first”).
- Questions about **personal experience** of health conditions.
  - Whether the respondent had or hadn’t experienced a mental health (MH) condition at some point (or a prefer-not-to-say option). If answered positively, categorical questions about what sort of condition that was, and whether it was recent or >5 years ago, and what sort of support had been obtained (e.g. from NHS or other sources).
  - Whether the respondent had ever used physical health services, and if so, which categories.
- Fictional examples of **identifiable health data** were presented. These included identifiers, diagnoses, and notes.
- A **framing statement** was presented, randomized to be one of the following:
  - *Neutral:* “We would like to find out your perspective on using information about your mental health and your physical health.”
  - *MH concern:* “Previous surveys have found that people have more concerns about the use of their identifiable health data relating to their mental health than other aspects of their physical health care.” This statement was based on previous findings.^1^
  - *Holistic:* “Mental and physical illnesses overlap, so holistic health care is important. Mental health problems have physical consequences, and physical illnesses have important consequences for mental health.” This statement is also true (e.g. ^2, 3^).
- Likert-style questions about how likely the respondent would be to share their *identifiable physical health* data, for *clinical care* purposes, with a range of NHS “destinations”, without being asked each time.
- Likert-style questions about how likely the respondent would be to share their *identifiable mental health* data, for *clinical care* purposes, with a range of NHS destinations, without being asked each time.
- Fictional examples of **de-identified structured health data** were presented (the same examples as before, now de-identified, and without any free text). These included alphanumeric research identifiers, “blurred” demographics (age, sex, geographical region), and diagnoses.
- Likert-style questions about how likely the respondent would be to share their *de-identified (structured) physical health* data with a range of research destinations, without giving consent each time.
- Likert-style questions about how likely the respondent would be to share their *de-identified (structured) mental health* data with a range of research destinations, without giving consent each time.
- Fictional examples of **de-identified free-text health data** were presented. These were the same de-identified data as before, but now with de-identified versions of the free-text notes. Explicit commentary was given that there was more information present, and a slightly higher risk of inadvertent identification. We gave the example of a hypothetical newspaper report that might enable someone to re-identify a patient.
- Likert-style questions about how likely the respondent would be to share their *de-identified (free text) physical health* data with a range of research destinations, without giving consent each time.
- Likert-style questions about how likely the respondent would be to share their *de-identified (free text) mental health* data with a range of research destinations, without giving consent each time.
- An explicit change of topic was noted.
- We asked Likert-style questions about how likely people would be to sign up to a **single system for controlling how one’s NHS data is used** (clinically and for research), for a variety of types of system (e.g. online, in person), who should look after such consent-related data, what the respondent’s preferred method would for changing their preferences, and overall how likely they would be to sign up to such a system.
- We showed a **specimen consent form** for such a system (see below for full details).
  - First, the specimen form asked about sharing data for direct health care purposes.
  - ▪ It defined “confidential patient information” ^4^, and set the context in terms of health care being provided directly to the respondent.
  - ▪ It offered a yes/no decision: “I agree that all NHS care providers and professionals may share my confidential patient information with each other for the purposes of my treatment and care.”
- Second, the specimen form asked about the use of de-identified data for research.
- ▪ It provided brief information about the NHS’s promises to use anonymised data for research (e.g. ^5, 6^). It said that research was conducted by the NHS and by NHS-approved researchers such as universities. It referred to strict security controls and NHS oversight.
- ▪ It offered a yes/no decision: “I agree that all NHS care providers may share my confidential patient information with each other and de-identify it for the purpose of research.”
- ▪ We phrased the question in this way because it is already permitted, given NHS research ethics approvals, for NHS bodies to de-identify health data for research.^7, 8^ However, cross-site linkage within the NHS is more challenging; if linkage is conducted with direct identifiers such as NHS numbers, this is work involving confidential patient information for research, which requires either explicit consent or approvals (in England) under section 251 of the NHS Act 2006 ^4^ (as amended) and the Health Service (Control of Patient Information) Regulations 2002.^9^ That is the case even if the data are de-identified subsequently and researchers never see identifiable information.
- ▪ We said also that saying no would not prevent all uses of one’s confidential information for research. At present, in England, the NHS National Data Opt-Out is the mechanism to opt out from uses of one’s confidential personal information where that use is governed by section 251 of the NHS Act.^10, 11^
  - Third, the specimen form asked about taking part in research.
  - ▪ It discussed briefly research involving direct participation, stating that the NHS promised to inform people of research studies for which they may be eligible,^5, 6^ and saying that there is never a commitment to take part.
  - ▪ It offered a yes/no decision: “I agree that NHS-approved researchers may learn my identity and contact me directly about research studies for which I may be eligible.”
- Having showed the respondent the specimen consent form, we asked Likert-style questions about its clarity, how the respondent understood its meaning, whether some additional aspects should be added, whether the respondent was previously aware of the NHS National Data Opt-Out, and whether the respondent would choose to sign such a form if it were available to them today.
- We asked, via a multiple-choice question, whether **sign-up portals for research** should be multiple (as they are now), single (across the NHS), or split by mental/physical health research.
- We asked about the respondents’ preference for **linkage of their health data to other data sources** for research. We set out a basic method commonly used for identifiable linkage, in which special permissions are sought, trusted third-party linkage is conducted (using identifiable information), followed by de-identification for research. We asked whether the respondent would be happy (yes / not sure / no / prefer not to say) for their health data to be linked to a number of “state” sources (education, police/criminal justice, transport, housing, immigration, social security), giving simple examples of the potential research reasons for each linkage, plus universities (e.g. if the respondent had volunteered for research studies) and data held by private companies.
- Finally, we asked optional **demographic** questions: gender; age range; ethnicity; sexuality; religion; employment status, with conditional questions sufficient to determine the UK Office for National Statistics (ONS) National Statistics Socio-Economic Classification (NS-SEC)^12^ via the “self-coded” method^13^ if the respondent answered in full; UK nation of residence; and (if the respondent was willing) their postcode, to calculate a “blurred” geographical version as described below. We used ONS demographic categories where available. We offered the option to leave an e-mail address to receive a summary of results when available.
- The survey closed by thanking the participant (not shown in the Appendices).

The REDCap design for the survey is shown in **Appendix A,** and the resulting survey (including the specimen consent form) is shown in **Appendix B.**

#### S1.2. Analysis of geography including Index of Multiple Deprivation

Where participants agreed to leave a postcode, this was converted to a larger ONS geographical area, so that individuals could not inadvertently be identified. In turn, the larger geographical area was converted to an Index of Multiple Deprivation (IMD), a composite rank measure covering income, employment, education, health, crime, barriers to housing and services, and living environment.^14^ A lower raw IMD indicates greater deprivation. For England and Wales, postcodes were converted to a Lower Layer Super Output Area; these have a minimum population of 1,000 and a mean population of 1,500.^15^ For Scotland, the Data Zone was used; for Northern Ireland (NI), the Super Output Area (SOA). Here, we use the term “geographical area” (GA) for LSOA (England and Wales), DZ (Scotland), or SOA (NI). We used GAs from the 2011 Census.

GAs were converted to a UK-wide IMD score,^16, 17^ using the scale relative to England, in which high numbers represent greater deprivation. As this UK-wide data set uses 2001 DZs for Scotland, we mapped these to 2011 DZs.^18^ Where multiple 2001 DZs mapped to one 2011 DZ, we took the mean of their UK-wide IMD scores, weighted by their population contribution to the 2011 DZ.^18^

Since GAs do not have equal populations, we corrected for population when calculating centile of deprivation. We used mid-2019 ONS estimates of GA population.^19–21^ We calculated deprivation centile (100% meaning most deprived) by calculating, for each GA, “what percentage of the total UK population (66,796,806) live in a GA with a UK IMD score that is equal to or lower than that of this GA (i.e. in areas that are equally or less deprived than this GA)?”. We calculated quartiles similarly.

To show the distribution of deprivation, we plotted the distribution of deprivation centile using a Gaussian kernel density estimate.

For map representations, the Apr 2019 ONS local authority district boundaries^22^ (which exclude the Channel Islands) were used with the Nov 2019 ONS Postcode Directory.^23^

#### S1.3. Sensitivity analysis via survey weighting

As a sensitivity analysis to correct for unrepresentative demographic sampling during general linear modelling, we used raking,^24^ specifically the American National Election Study (ANES) weighting algorithm^25^ via the *anesrake* package.^26, 27^ We defined dimensions (classification variables) and categories (within dimensions) as shown in **Supplementary Table 1,** collapsing across some low-frequency categories in the survey. We included all such variables in the raking, and used the default weight cap of 5.

The algorithm does not alter weights for categories for which population expected proportions are not known (e.g. those identifying as neither male nor female, for which Census data are not available, or those answering “prefer not to say” for a given question). Such respondents are therefore assigned a weight of 1 in that category, representing them fairly in the absence of any other information with which to weight them.

Population values for some questions were not available; for example, sexual orientation was not part of the UK 2011 Census.^28^

We weighted all respondents once only, consistently (ignoring the potential for discrepant drop-out rates across the survey).

We used weighted versions of the statistical models (see **Methods**) that did not include demographic predictors, labelling these with “W” (e.g. model CW1 was a weighted version of model C1).

#### S1.4. Effect size plots

We show effect sizes with 95% confidence intervals from selected statistical models. We use the conventional language of “factor” (discrete predictor), “levels” (possible values of a factor), and “term” (individual predictive term in a GLM, such as “destination” or “destination × nature”). Effect sizes are shown as uncorrected pairwise comparison to a reference category within each term. For age, we used the central category (age range 45–54) as the reference category, because there were very few participants in the “under 18” category. For brevity, we restrict plots to those model terms with a significant *F* test. We show degrees of freedom to 1 decimal place and statistics to 3 significant figures.

*F* tests, as for the main results, were taken from *anova(model)* via *lmerTest::anova.lmerModLmerTest*, giving type III sums of squares via the Satterthwaite method for degrees of freedom.^29^ Type III sums of squares test the effect of each term “over and above” others.^30^ Pairwise comparisons were taken from *summary(model)* via *lmerTest::summary.lmerModLmerTest*, also using Satterthwaite’s method.

We note the important difference between these pairwise contrasts, helpful for basic visual display, and the omnibus *F* test for the term.^30^ In particular, we note firstly that for a single factor with three levels (A, B, and C with A as the reference level), the omnibus *F* test tests the null hypothesis A = B = C; if this null hypothesis is rejected, it remains possible that neither the pairwise hypothesis A = B nor the hypothesis A = C is rejected (for example, if the ordering is B < A < C). Thus, failure to observe differences in the specific pairwise contrasts does not imply that there are no differences in the data. Secondly, a similar effect can be observed with interactions: pairwise contrasts within interaction components may sometimes be suboptimal compared to subgroup simple effects analyses.^30^ Thus, we used simple effects analysis for detailed follow-up of significant interactions where appropriate.

#### S1.5. Free-text values and comments

Some respondents wrote in responses for “other” categories. Some provided free-text comments where invited to do so. A few e-mailed the study team separately. We provide narrative summaries of these answers, paraphrasing to avoid direct quotations and reporting “*n* < 10” where appropriate to mask small numbers.

For free-text comments on the proposed draft national consent form, we conducted a thematic analysis and report tallies by theme.

### S2. Supplementary Results

#### S2.1. Recruitment and completion

**Supplementary Figure 1** shows recruitment sources, recruitment rates, and survey participation by stage.

#### S2.2. Demographics

A demographic breakdown is shown in **Supplementary** Figure 2.

Free-text values provided in response to “other, please specify” categories included:

- *Mental health conditions* (*n* = 274): many unique responses, with non-unique responses distinct from the options offered including: ADHD [attention-deficit/hyperactivity disorder], Asperger’s syndrome/autism/autistic spectrum disorder, bereavement, body dysmorphic disorder, dissociative disorders, gambling disorders, insomnia, low mood, memory problems, menopause-associated symptoms, non-epileptic attack disorder, overdose, post-traumatic stress disorder, postnatal depression, suicidality, and stress.
- *Mental health services used* (*n* = 254): many unique responses, with non-unique responses including: counselling services, military services, specific named charities (including Mind and Samaritans), university mental health support services, and well-being services.
- *Physical health services used* (*n* = 258): many unique responses, with non-unique responses including: chiropractors, complementary therapists, dentistry, maternity services, NHS web sites, osteopaths, pharmacists, physiotherapists, and well-being services.
- *Gender* (*n* = 29): non-unique responses included: agender, gender fluid, non-binary, trans/transgender.
- *Ethnicity* (*n* = 150): a large number of responses more detailed than the ONS options on offer, with some critical of the premise.
- *Sexuality* (*n* = 65): a number of unique options and some critical of the premise or phrasing, plus some with multiple responses (e.g. asexual, panromantic, pansexual).
- *Religion* (*n* = 219): a range, some more closely specifying options offered (e.g. agnostic, atheist, Baptist, Catholic, Church of England, humanist, Jehovah’s Witness, Methodist, Orthodox, Quaker) and some for beliefs not listed (e.g. Baha’i, druid, Jedi, pagan, pantheist, spiritualist, Wiccan), including a number of unique responses.
- *Geography* (England *n =* 43, Scotland *n* < 10, Wales *n =* 20, Northern Ireland *n* < 10): a range of geographical divisions not matching the categories offered.

#### S2.3. Experience

Participants’ experience of mental health conditions/service and physical health services are shown in **Supplementary** Figure 3.

#### S2.4. Raking

The raking algorithm converged stably, and substantially improved the match to population marginal proportions, though was unable to make the resulting weights match the population marginals completely within the constraints specified. We show observed and raked proportions in **Supplementary Table 1**.

#### S2.5. Weighted analysis of sharing preferences

Weighted analyses (**Supplementary** Figure 4) were quantitatively similar to the primary results (compare Figure 3), and statistical analyses showed the same key patterns, although with somewhat weaker framing effects.

For clinical purposes (model CW1, *n* = 24497), there were effects including destination (*F*_3,181628_ = 6310, VLP), nature (*F*_1,181756_ = 4860, VLP), nature × framing (*F*_2,181756_ = 21·3, *p* = 5·41 × 10^−10^), destination × nature (*F*_3,181624_ = 5·84, *p* = 0·000557), and destination × framing (*F*_6,181628_ = 4·38, *p* = 0.000199). In sub-analyses of the framing effects (analysis of MH and PH data separately using model CW1B), there was no effect of framing for PH data (*F* < 1, NS) but this time the framing effect for MH data was not itself significant (framing, *F*_2,13607_ = 1·74, NS; destination × framing, *F*_6,84700_ = 1·91, *p* = 0.0748). As for the main analysis (model C1), the framing effects were slightly smaller for geographically broader (e.g. UK-wide) sharing, driving a destination × framing interaction via PH data.

For research purposes (model RW1, *n* = 23869), main effects included destination (*F*_5,547684_ = 74700, VLP) nature (*F*_1,547914_ = 1613, VLP), detail (*F*_1,548574_ = 5270, VLP), and framing (*F*_2,13472_ = 3·09, *p* = 0·045). Interactions included destination × nature × detail (*F*_5,547655_ = 8·64, *p* = 8·64 × 10^−8^) and nature × framing × pandemic (*F*_2,547914_ = 5·25, *p* = 0·00536). In sub-analyses of the framing effects (analysis of MH and PH data separately using model RW1B), framing main effects were not independently significant, though there were destination × framing interactions as before.

#### S2.6. Weighted views on linkage to non-health data for research

**Supplementary** Figure 5 shows weighted views on linkage to non-health data for research (compare Figure 4).

#### S2.7. Effect sizes

**Supplementary** Figure 6 shows effect sizes for models C1, R1, and L1. These models include data from all participants who answered the relevant questions, including those who did not supply full demographic information (compare Figure 6).

#### S2.8. Weighted views on a national data sharing consent system

**Supplementary** Figure 7 shows weighted views on a national data consent system (compare Figure 7).

#### S2.9. Free-text comments on the suggested consent form

Themes of free-text comments in relation to the proposed consent form (*n* = 3112) included:

- Form design:
- *Too long/complex* (*n* = 732). The form was too long and/or complex, or the text (or aspects of the text) should be clearer or simpler, including statements that the reader found it OK but felt that others (not specified in detail) would not.
- *Visual style* (*n* = 395). Regarding visual clarity and style (e.g. typography, layout, colours, and the spelling of ‘organization’ versus ‘organisation’).^31^
- *Clear/good* (*n* = 159). The form was satisfactory, good, clear, and/or concise.
- *Extra information or accessible versions* (*n* = 271). There should be accessible forms, easy-read versions, alternative language versions, or accompanying explanations (e.g. leaflet, illustrations, video, explanations of the benefits and risks of data sharing).
- *Too brief* (*n* = 18). It was too brief, and/or required more detail or explanation (in general).
- *Biased* (*n* = 16). It was biased, coercive, or misleading in its questions.
- Method of completion:
- *Digital* (*n* = 69). A digital or online version would be desirable or preferable (or in some cases that a paper copy would be desirable or necessary as well).
- *Non-digital* (*n* = 26). A paper copy would be desirable or necessary, or that not everyone could use internet-based methods.
- *One-to-one support* (*n* = 90). One-to-one support (e.g. face-to-face, e-mail, telephone) would be desirable or necessary (from a clinician or other unbiased person, including support for those who may lack capacity to decide, and for children).
- *Copies* (*n* = 27). People should have copies of what they agreed to.
- *Changing preferences* (*n* = 96). It should be easy to revisit and update one’s preferences and/or contact details (and/or clarity on what would happen to data previously shared under such circumstances).
- *Enough time* (*n* = 49). It is important people have time to complete the form and do not feel pressured to do so, and/or are not asked during times of personal medical crisis.
- Legal aspects:
- *Legal* (*n* = 58). That the Data Protection Act and/or European Union (EU) General Data Protection Regulation (GDPR), Access to Health Records Act, Information Commissioner’s Office, NHS privacy documents (e.g. privacy impact assessments, Caldicott Guardian framework), or mental capacity frameworks (e.g. Mental Capacity Act, Lasting Power of Attorney) should be referenced explicitly, or views that the proposed system/form would be (or that existing NHS information systems are) incompatible with one of these.
- Managing data related to consent:
- *Data for the consent process* (*n* = 83). Relating to self-identification when completing the form: that asking for an NHS number is problematic as people may not know this, that some identifiers (e.g. full name, address, e-mail address) are unnecessary or sometimes unnecessary, or that some (e.g. telephone number, previous names and addresses) should be added, or concerns about the management of identity information used for the consent form.
- Management of healthcare data:
- *De-identification* (*n* = 164). That aspects of de-identification/anonymisation/pseudonymisation needed to be clearer, including that asking for identity data whilst recording preferences about de-identified data use was confusing, or that any form of pseudonymisation (rather than full anonymisation) is undesirable, or that examples should be given of identifiable versus de-identified data.
- *Data security* (*n* = 594). That the security and privacy of healthcare data is of paramount importance, including that more detail should be provided about data security/sharing or privacy controls, or that the respondent did not trust the NHS or the UK to manage data security properly.
- *Centralization* (*n* = 30). Greater centralisation of data increases concern and risks.
- *Sharing desirable* (*n* = 290). Data sharing is desirable in general, for clinical or research purposes or not specified.
- *Sharing undesirable* (*n* = 74). Data sharing is undesirable in general, e.g. without case-by-case specific consent (± except in emergencies).
- *Opt-outs* (*n* = 134). That opt-outs should be more prominent, the default, that the system should be linked with the NHS National Data Opt-Out (directly or by implication), or that the relationship to NHS Act Section 251 approval was unclear.
- Categories of healthcare data:
- *Distinguish mental/physical health* (*n* = 45). Mental health and physical health data should be distinguished.
- *Combine mental/physical health* (*n* < 10). Mental health and physical health data should not be distinguished.
- *Distinguish health data in other ways* (*n* = 11). Further subtypes of health data (e.g. sexual health, drug/alcohol use) should be distinguished.
- *Distinguish free text from structured data* (*n* = 23). Free/narrative text should be distinguished from structured data.
- *Distinguish data by age* (*n* < 10). Data should be distinguished by its age (historical versus recent).
- Categories of use:
- *Clinical versus research* (*n* = 44). Clinical and research uses of data should be more clearly distinguished (e.g. not included on the same form).
- *Transparency of use* (*n* = 34). That people should be informed about, or be able to inspect, or choose regarding, each individual use of their data, and/or the results of research involving their data.
- *Profit-making* (*n* = 198). That profit-making use should not happen, or people should be paid directly for providing data used for profit, or that the NHS should ensure that it profits from such data use.
- Data or use/users, not distinguished:
- *More detail* (*n* = 74). More detailed options should be available (in general).
- End users:
- *Specify clinical users more* (*n* = 147). Permissions to share clinical data should not relate to the whole NHS, but be more detailed (e.g. local, regional, or regarding specific staff groups, or regarding private healthcare providers, or the relationship to social rather than health care).
- *Consent not needed* (*n* < 10). That consent (or at least explicit consent) is not required for aspects of data sharing, or should not be required.
- *Specify researchers more* (*n* = 268). That “NHS-approved researchers” is too vague, and that more detail about who such researchers are, or more detailed options to select from (up to and including case-by-case approval by the patient concerned), should be provided.
- *Third-party/private users* (*n* = 417). That healthcare data should not be available to private or third-party companies (or should require specific permission), including insurance companies and other profit-making organizations, or that later developments to provide them with data would be of concern.
- *Other state users* (*n* = 29). That use by other state/Government organizations would be of concern, or more detail would be required in that regard.
- *Overseas users* (*n* = 52). That use by overseas users (e.g. researchers) would be of concern, e.g. because other countries have different data regulations.
- Contact by researchers:
- *Contact by researchers* (*n* = 69). That more detailed options should be available about consent for research, or that being contact directly would be of concern (including about frequency of contact, and preferences that contact is always via clinical teams rather than researchers being given identifiable information).
- Overall objectives:
- *Good idea* (*n* = 128). The consent system or project was a good idea (in general).
- *Bad idea* (*n* = 16). The consent system or project was a bad idea, or unnecessary.
- Other:
- We did not tally comments relating to other aspects, such as the accuracy of health records themselves, patient access to health records (or holding the primary version of all their health data themselves), descriptions of an individual’s care, details of how the respondent would themselves choose, comments on the relationship to consent for clinical treatment, expressed wishes that the research team be shot, comments that people never read forms anyway, views that an independent body should oversee such a process, or concerns about research bias relating to who would or would not consent to their data being used.

#### S2.10. Other free-text comments

Themes of the free-text comments sent to the research team separately included:

- That the respondent’s preferences regarding mental versus physical health data had been driven by personal experience of information being misused (*n* < 10).
- That all research use of data should require informed consent, involving participants’ knowledge of the nature of the research (*n* < 10).

## S4. Supplementary Tables

**Supplementary Table 1.** Demographic dimensions and categories used for weighting, with UK population marginal proportions (*e*), observed marginal proportions amongst respondents (*o*, ignoring those who did not answer or answered via a category not acknowledged by UK population statistics), and marginal proportions after raking ( *r*, likewise). Proportions are shown to 3 decimal places; VLP, *p*<2·2×10^−16^.

| Variable (dimension) | Category | UK expected population proportion, $e$ | Reference for $e$ | Observed proportion (ignoring other/unknown), $o$ | Sample versus population proportions | Raked proportion (ignoring other/unknown), $r$ |
| --- | --- | --- | --- | --- | --- | --- |
| Age | Under 18 | 0.211 | <sup>32,33</sup> | 0.005 | $\chi^2_8=8860$, VLP | 0.024 |
|  | 18–24 | 0.085 |  | 0.050 |  | 0.082 |
|  | 25–34 | 0.135 |  | 0.139 |  | 0.167 |
|  | 35–44 | 0.126 |  | 0.163 |  | 0.159 |
|  | 45–54 | 0.136 |  | 0.218 |  | 0.177 |
|  | 55–64 | 0.122 |  | 0.228 |  | 0.160 |
|  | 65–74 | 0.100 |  | 0.150 |  | 0.133 |
|  | 75–84 | 0.060 |  | 0.043 |  | 0.078 |
|  | 85+ | 0.025 |  | 0.004 |  | 0.019 |
| Gender | Female | 0.506 | <sup>32</sup> | 0.673 | $\chi^2_1=2390$, VLP | 0.471 |
|  | Male | 0.494 |  | 0.327 |  | 0.529 |
| Ethnicity | White | 0.860 | <sup>34</sup> (data only available for England and Wales) | 0.910 | $\chi^2_1=442$, VLP | 0.861 |
|  | Other | 0.140 |  | 0.090 |  | 0.139 |
| Education | No formal | 0.232 | <sup>35</sup>; data for over-16s only; vocational data absent for Scotland (ignored in calculating proportions) | 0.025 | $\chi^2_4=13800$, VLP | 0.121 |
|  | Level 1 and Level 2 (approximating “secondary”) | 0.292 |  | 0.139 |  | 0.250 |
|  | Level 3 (approximating “A-Level/equivalent”) | 0.121 |  | 0.122 |  | 0.146 |
|  | Level 4+ (approximating undergraduate, postgraduate, and professional) | 0.270 |  | 0.589 |  | 0.364 |
|  | Vocational | 0.084 |  | 0.126 |  | 0.119 |
| Religion | No religion or not stated | 0.332 | <sup>36</sup> (Table B; data only available for England and Wales) | 0.618 | $\chi^2_2=10900$, VLP | 0.337 |
|  | Christian | 0.565 |  | 0.338 |  | 0.563 |
|  | Other | 0.102 |  | 0.043 |  | 0.100 |
| Nation | England | 0.843 | <sup>32</sup> (Channel Islands not included) | 0.994 | $\chi^2_3=3660$, VLP | 0.968 |
|  | Northern Ireland | 0.028 |  | 0.0002 |  | 0.001 |
|  | Scotland | 0.082 |  | 0.002 |  | 0.010 |
|  | Wales | 0.047 |  | 0.004 |  | 0.020 |
| Index of multiple deprivation | First (least deprived) quartile | 0.25 | By definition | 0.334 | $\chi^2_3=1620$, VLP | 0.247 |
|  | Second | 0.25 |  | 0.317 |  | 0.195 |
|  | Third | 0.25 |  | 0.233 |  | 0.279 |
|  | Fourth (most deprived) | 0.25 |  | 0.116 |  | 0.279 |

## S5. List of appendices

### Appendix A Underlying REDCap survey design

**Appendix A** shows a screenshot of the full REDCap design for the survey.

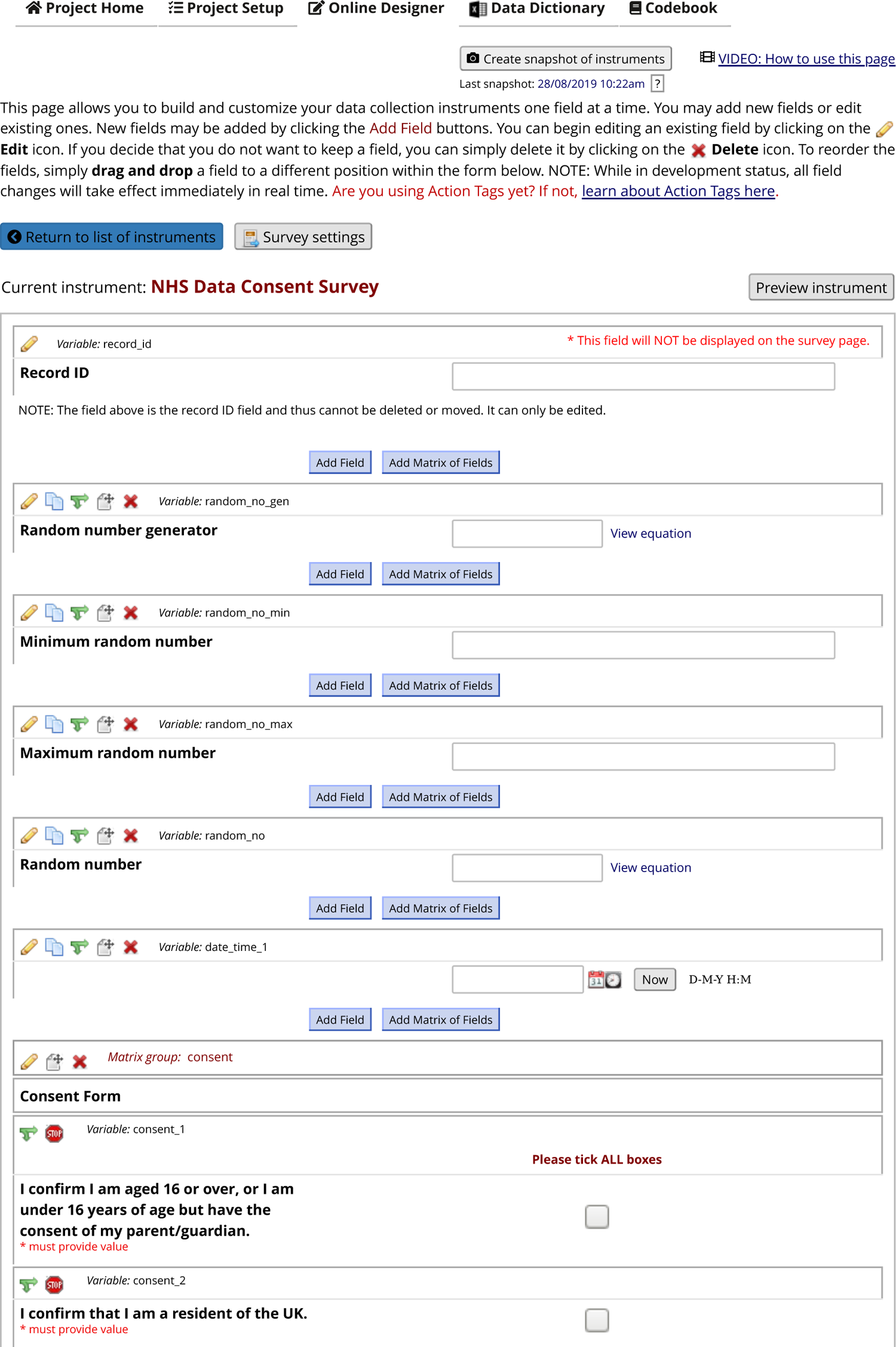
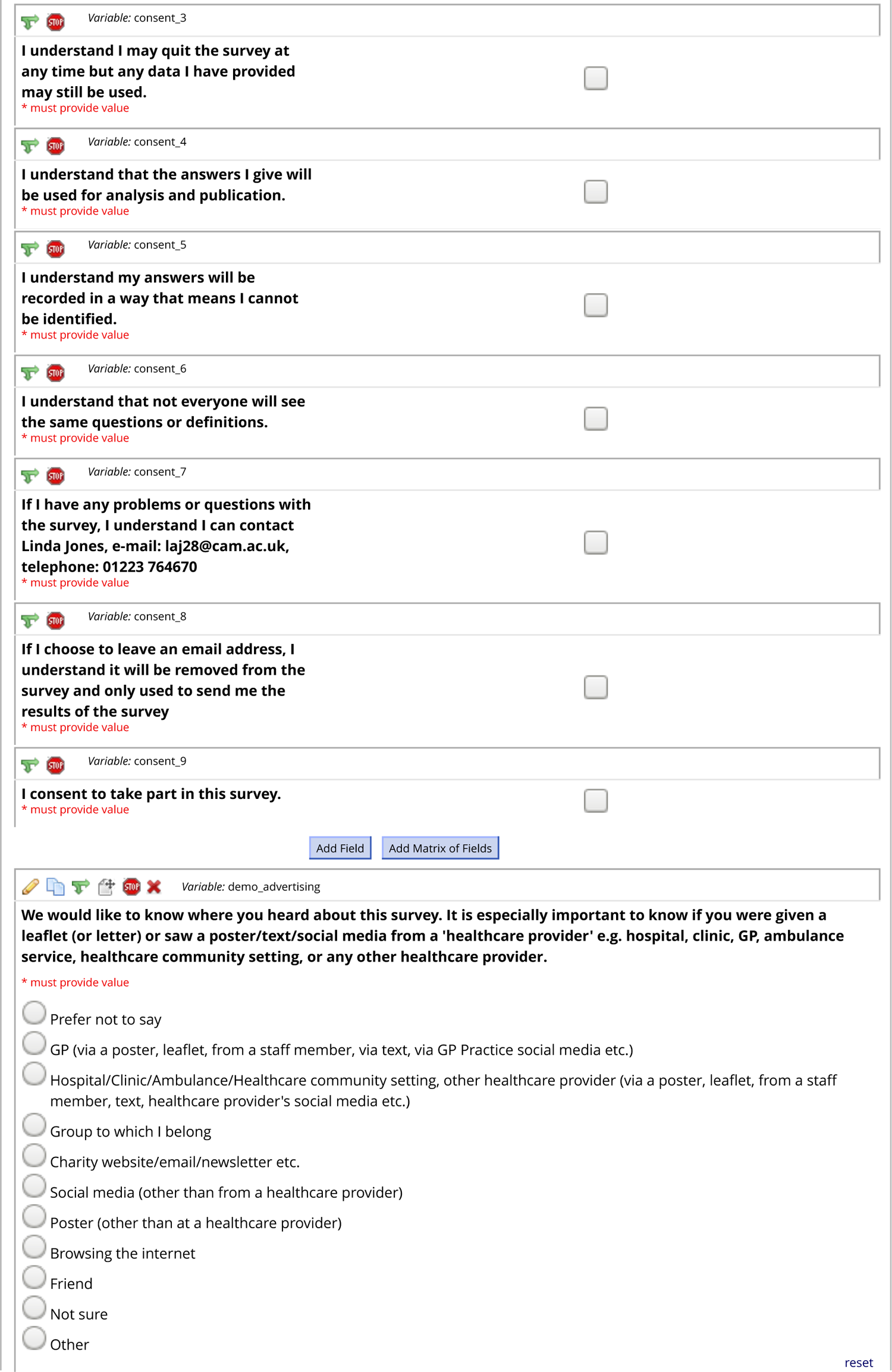
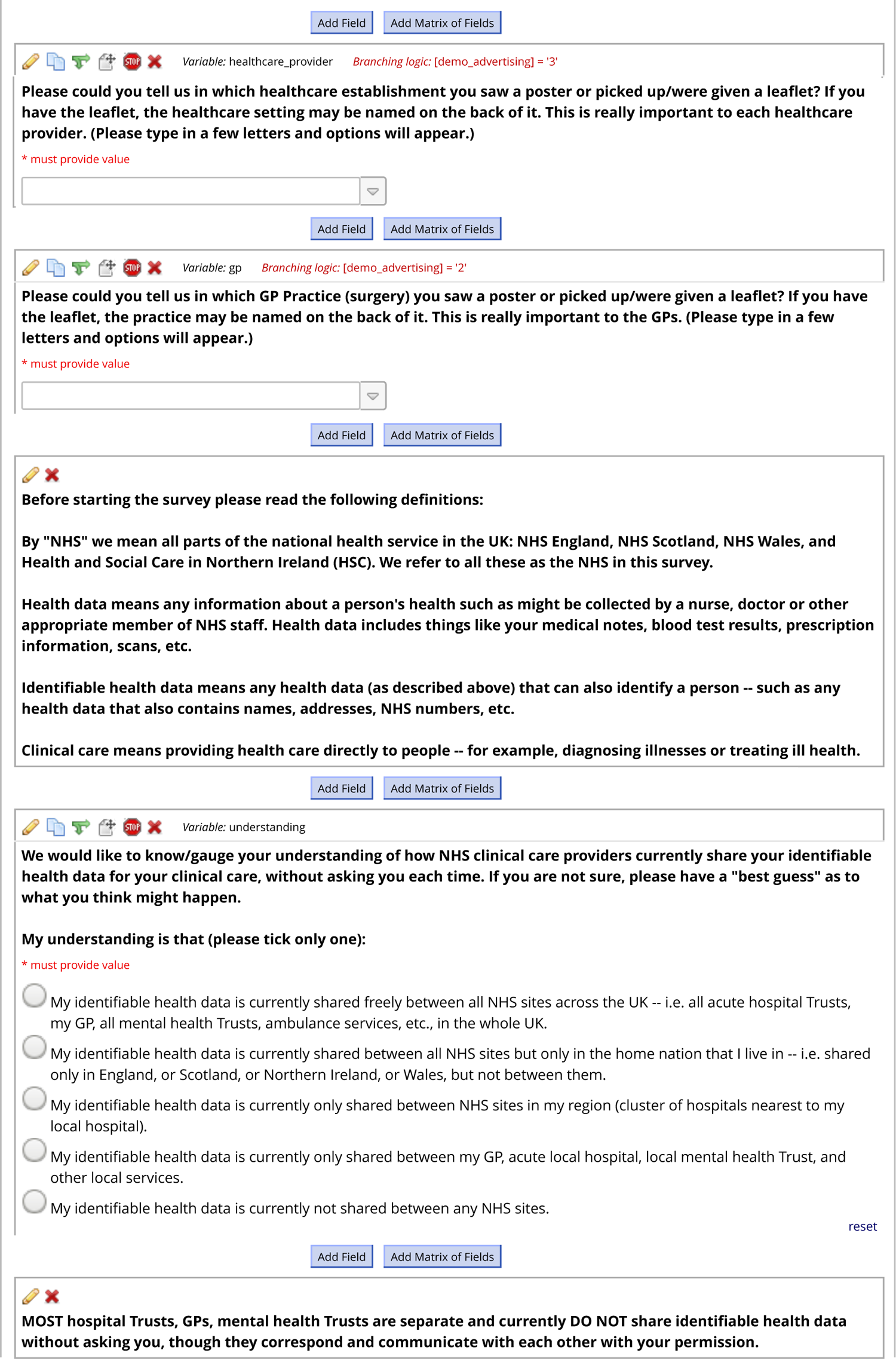
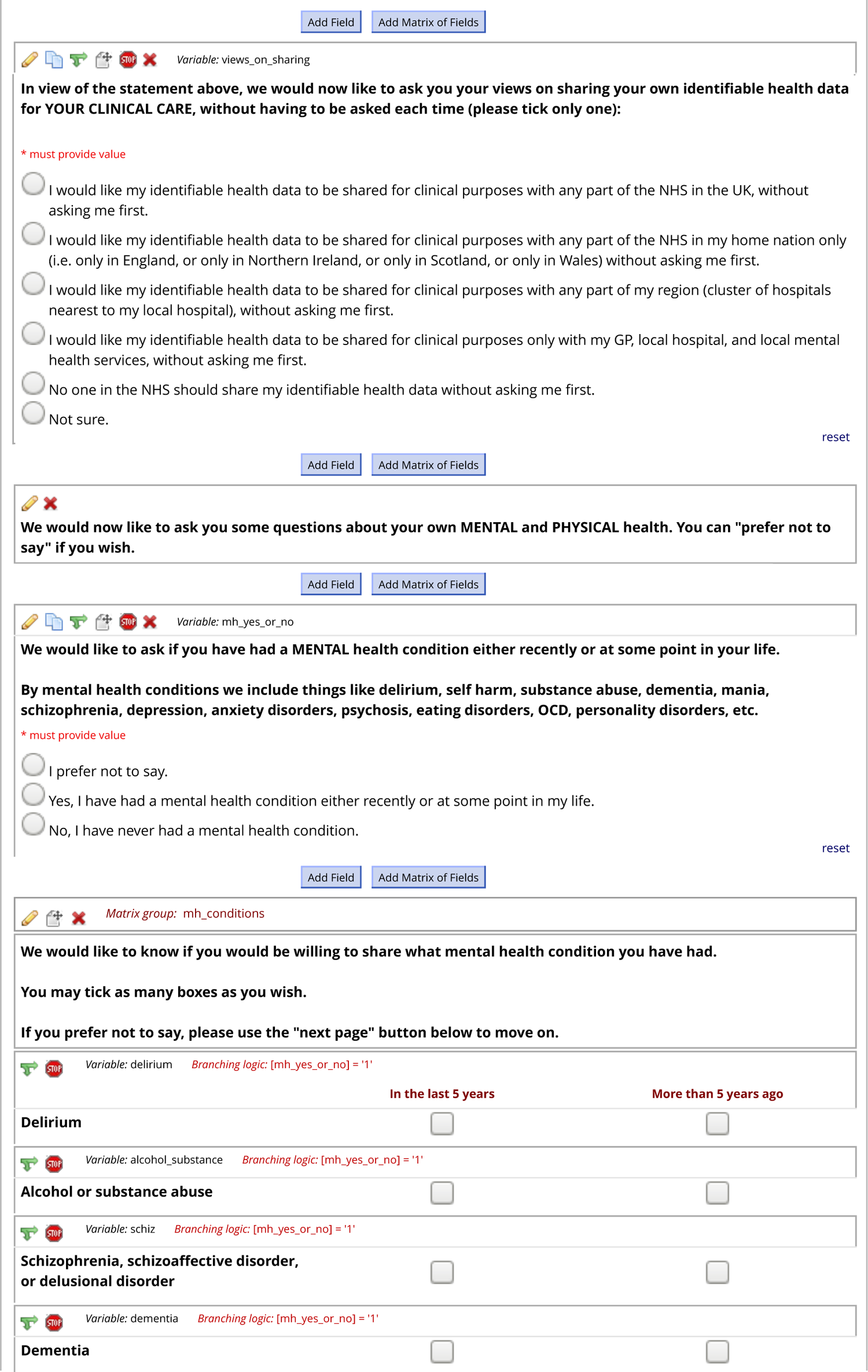
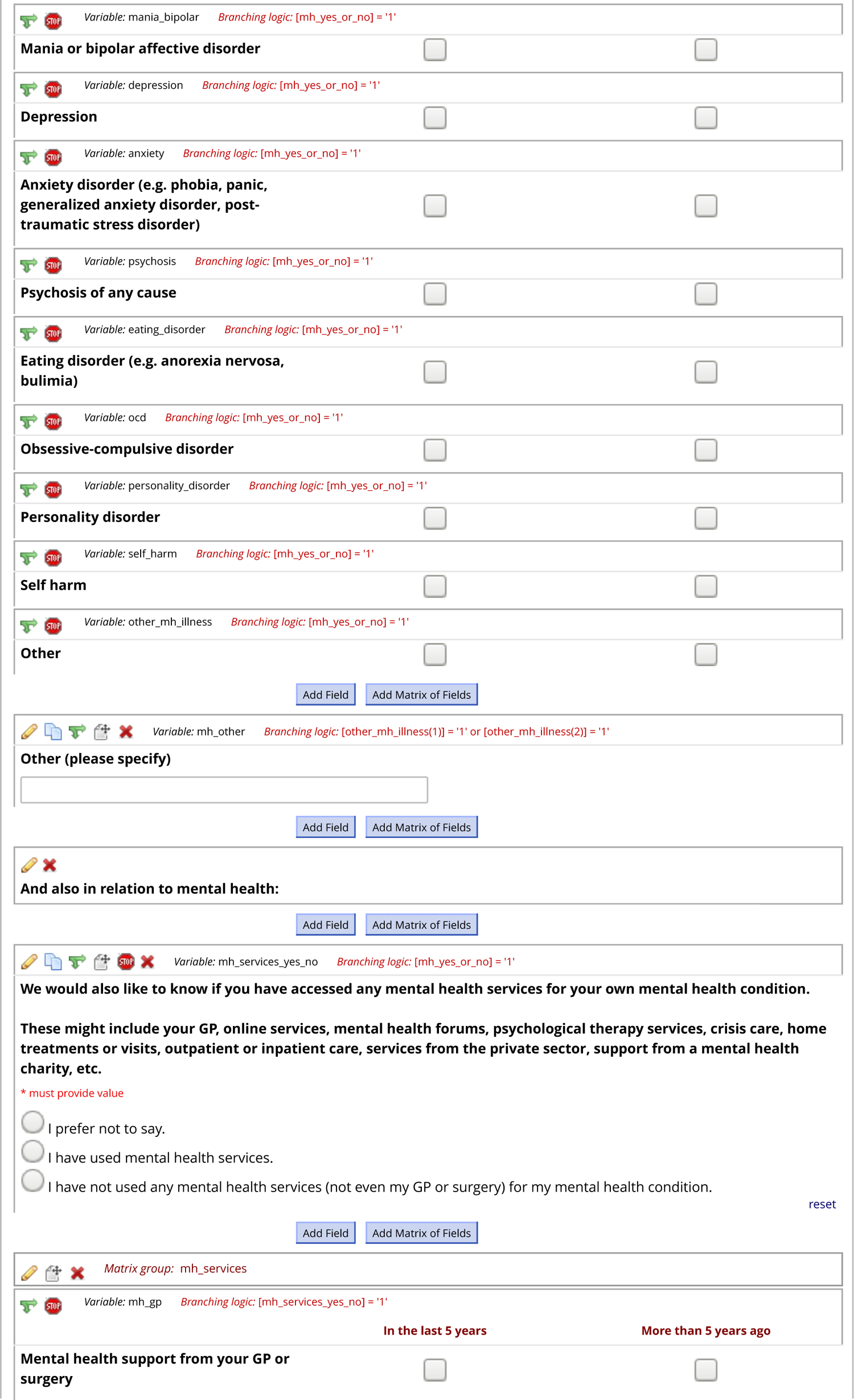
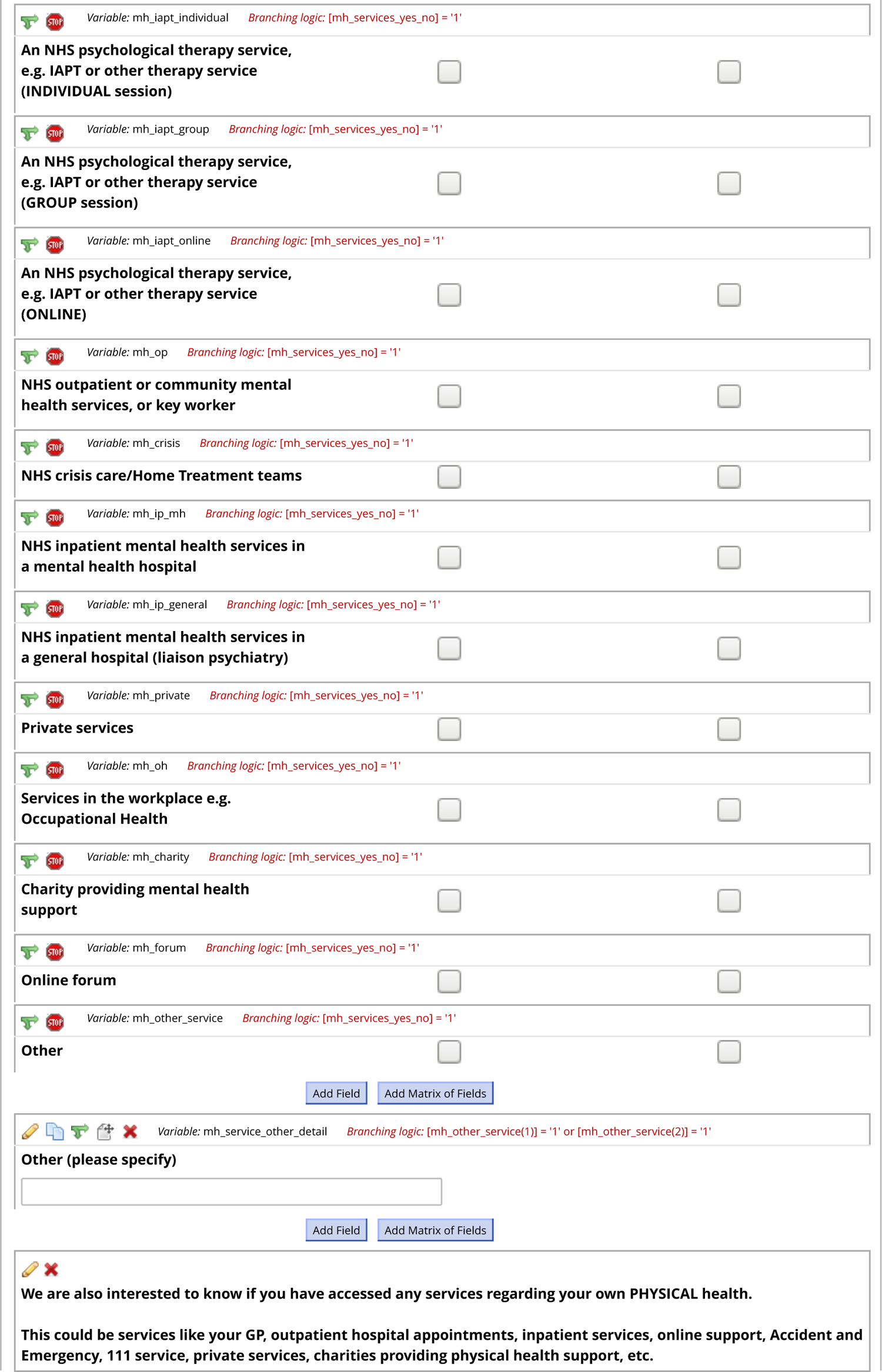
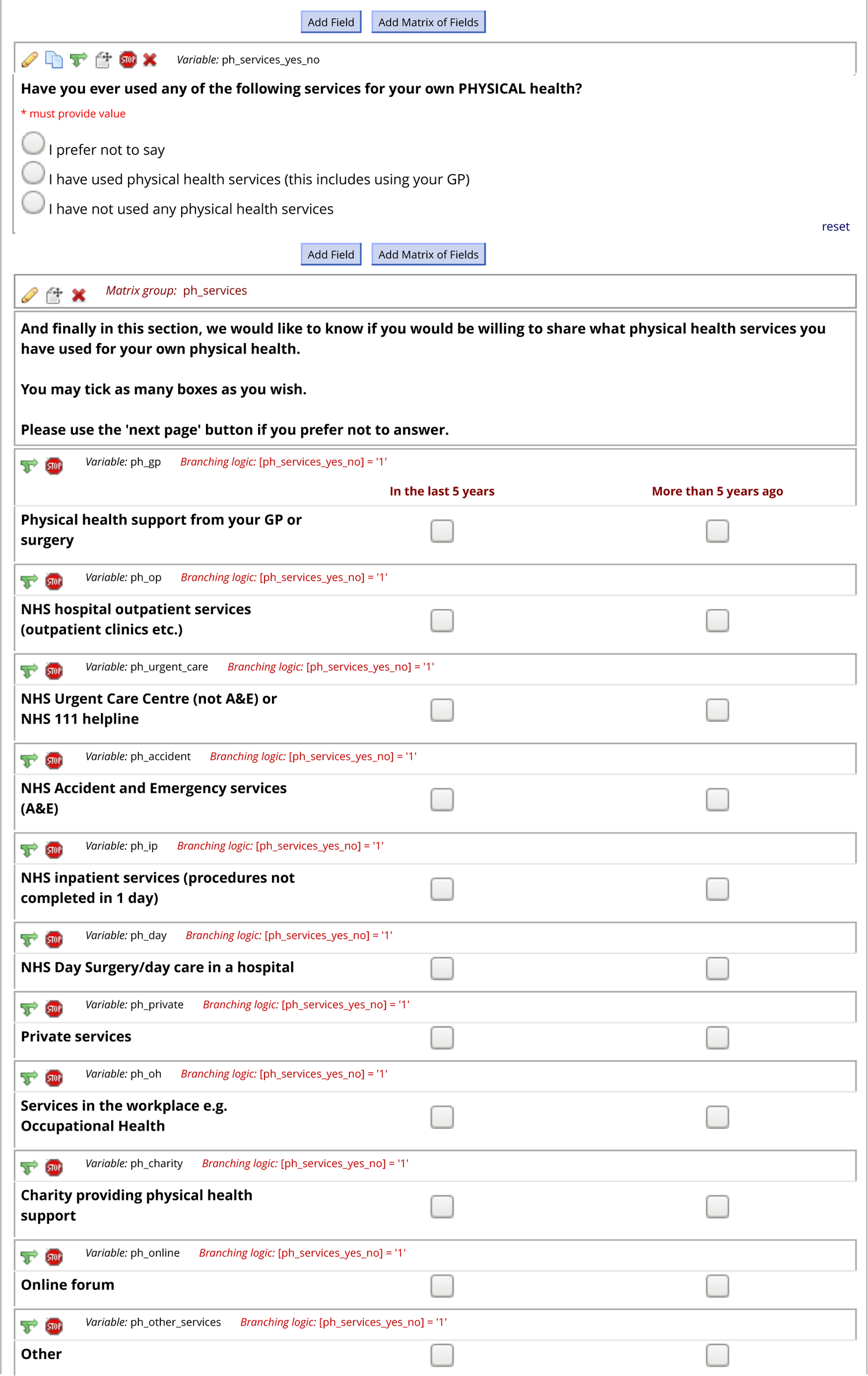
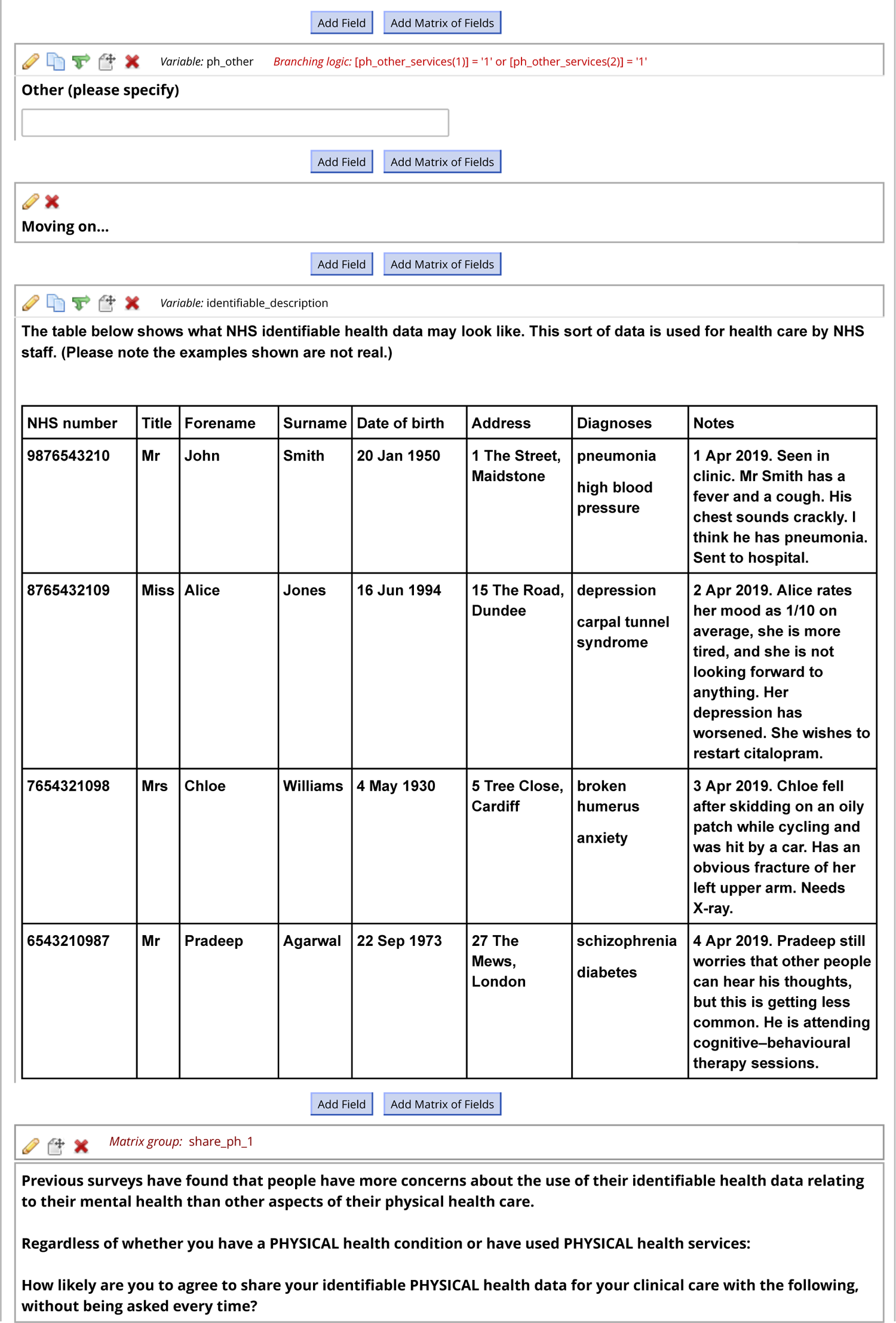
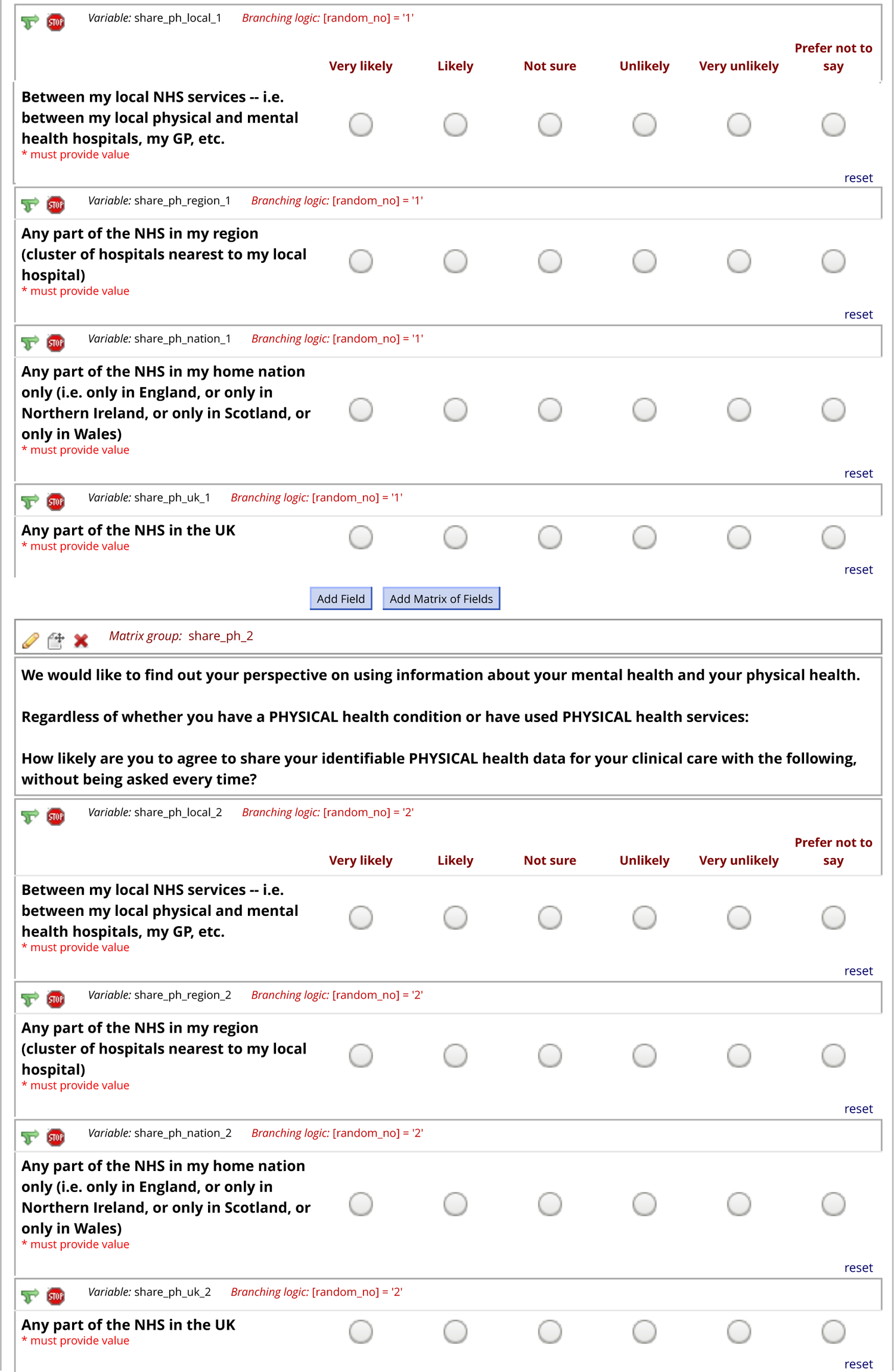
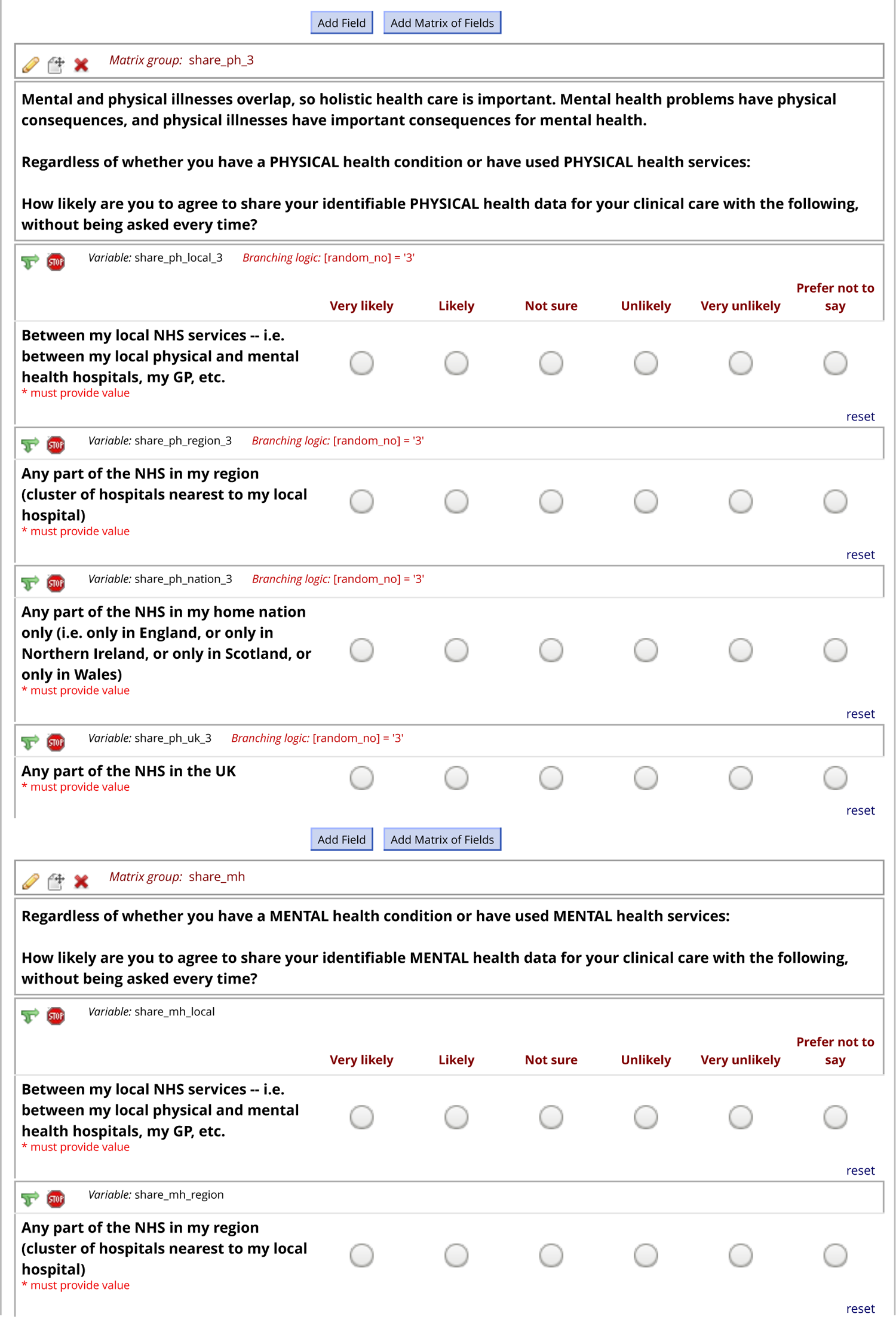
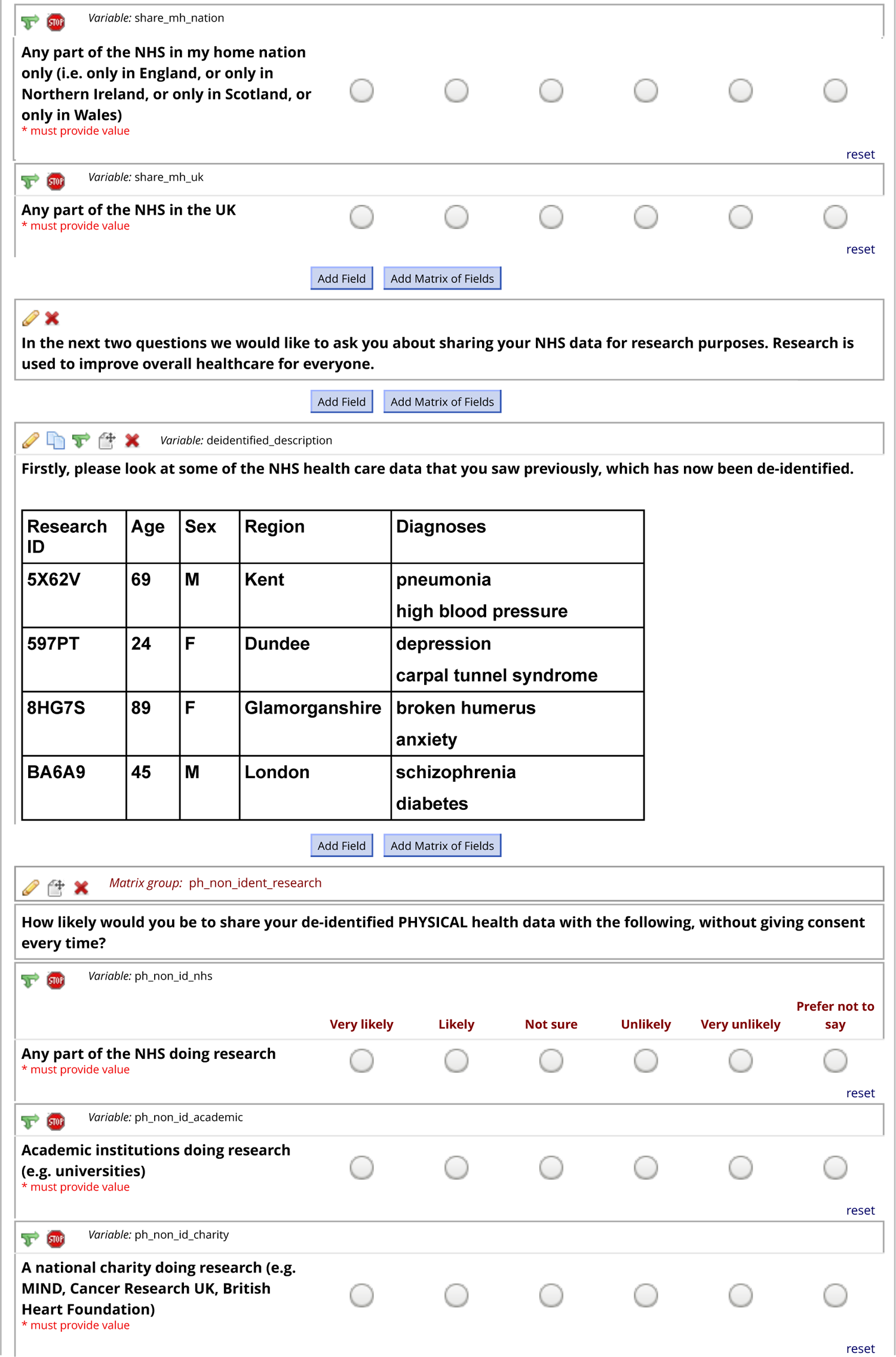
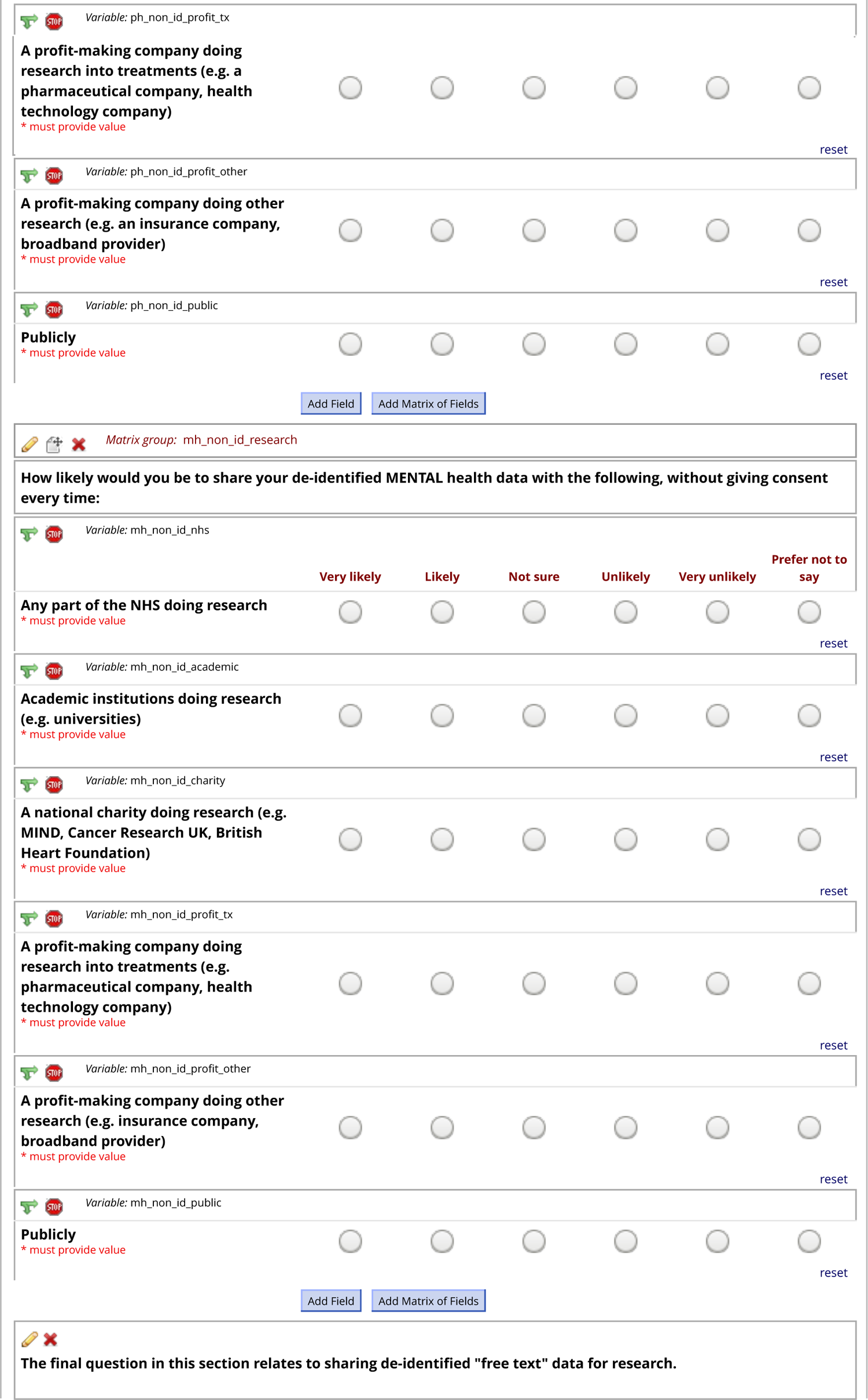
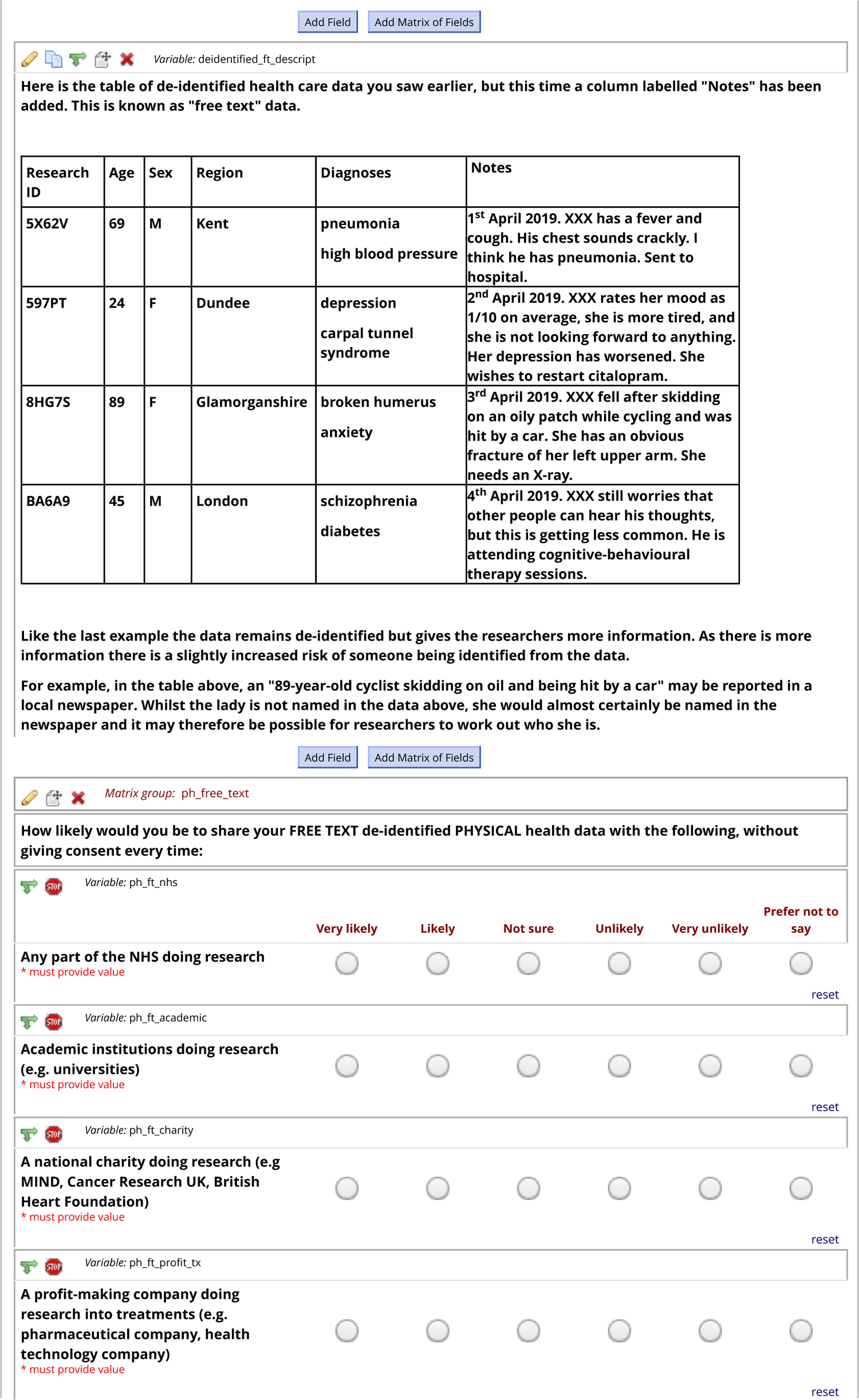
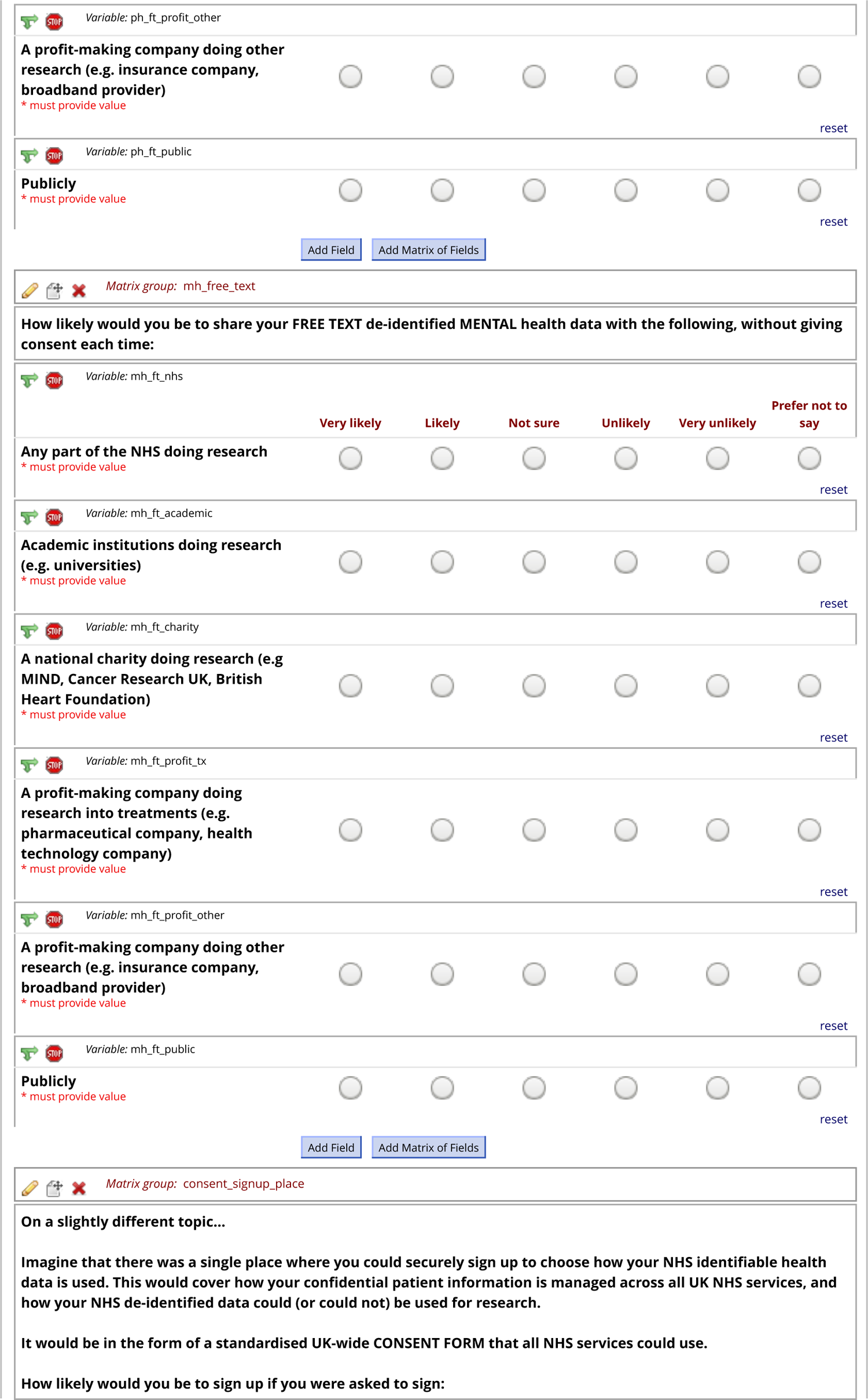
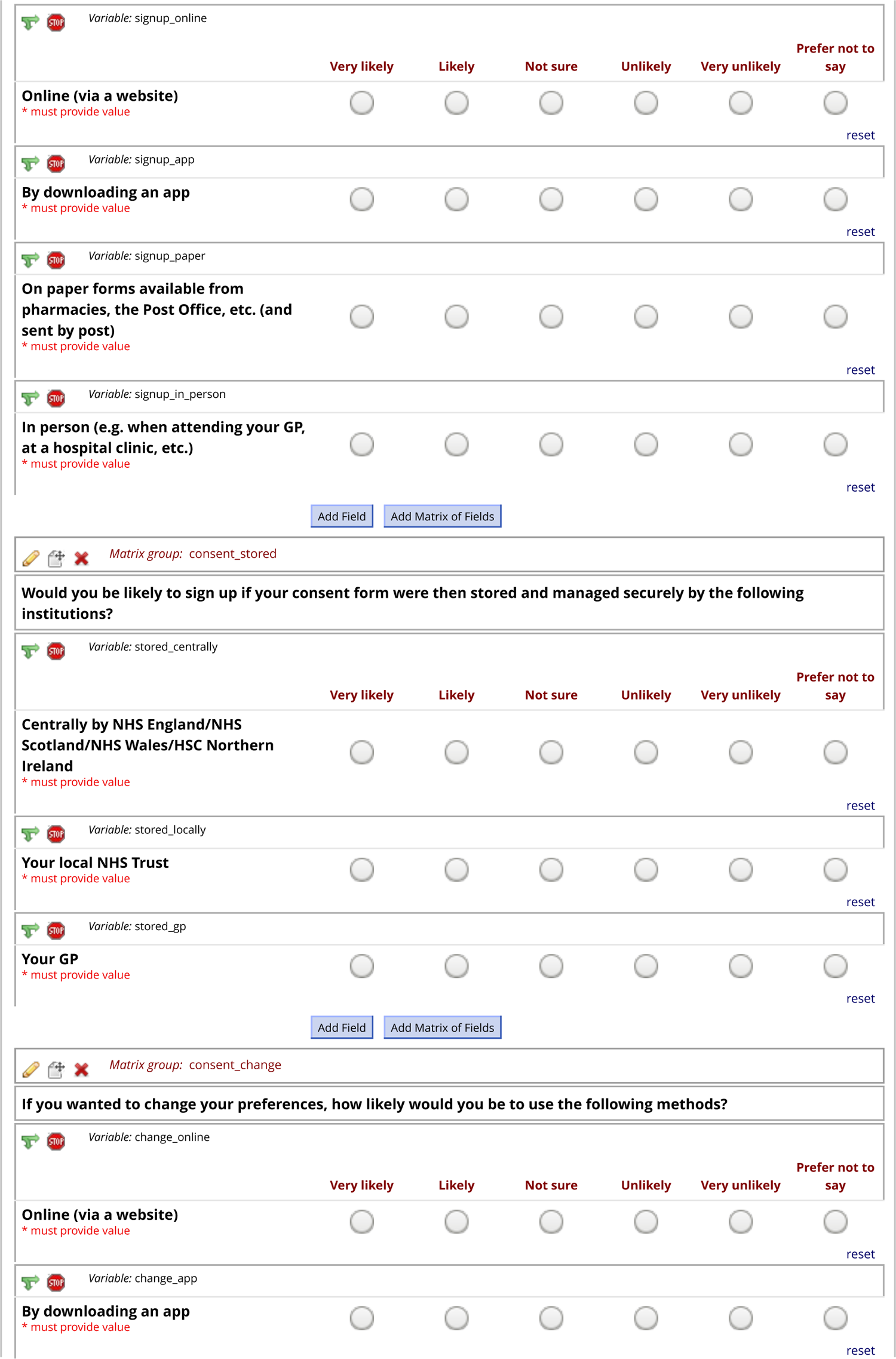
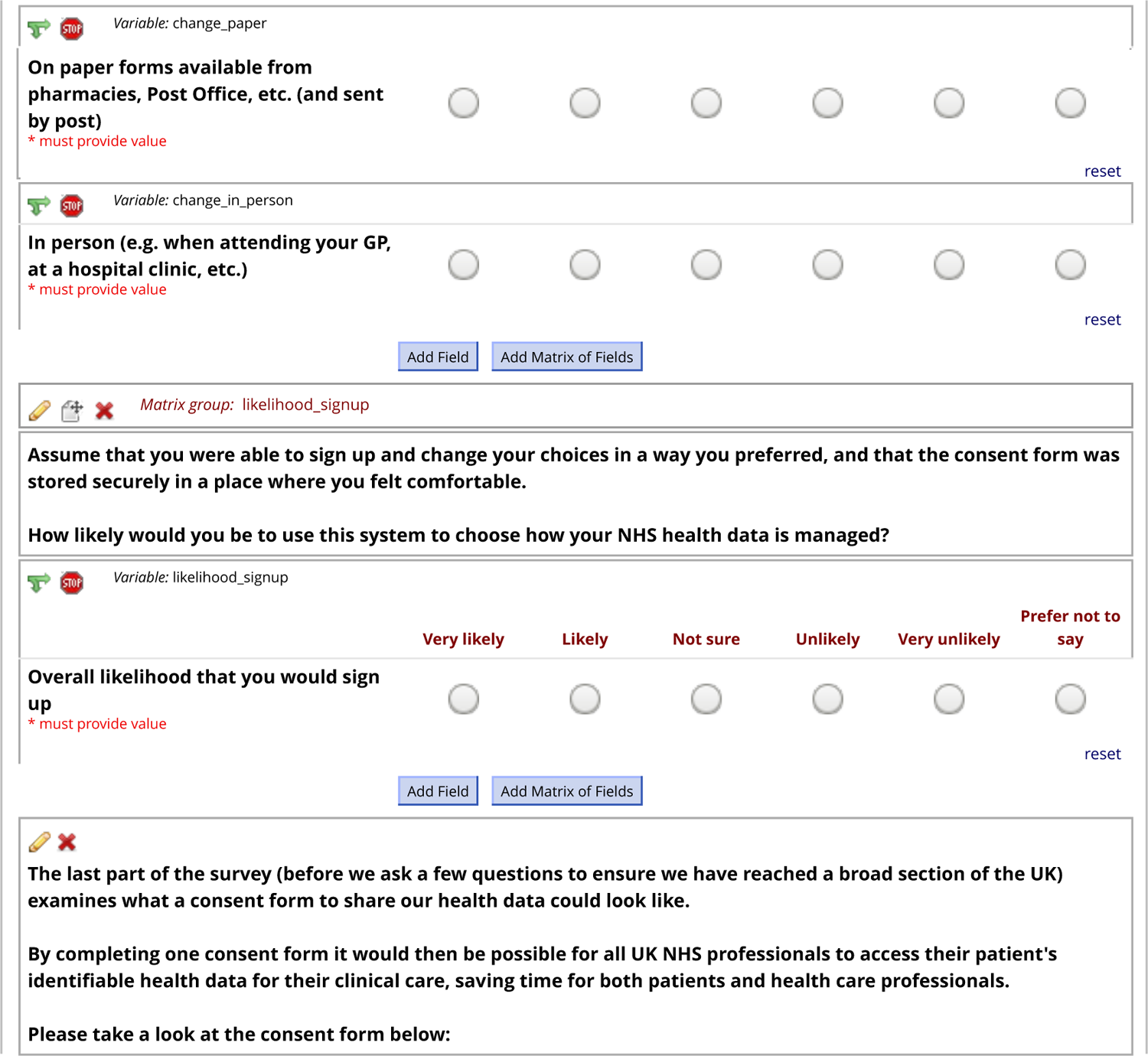
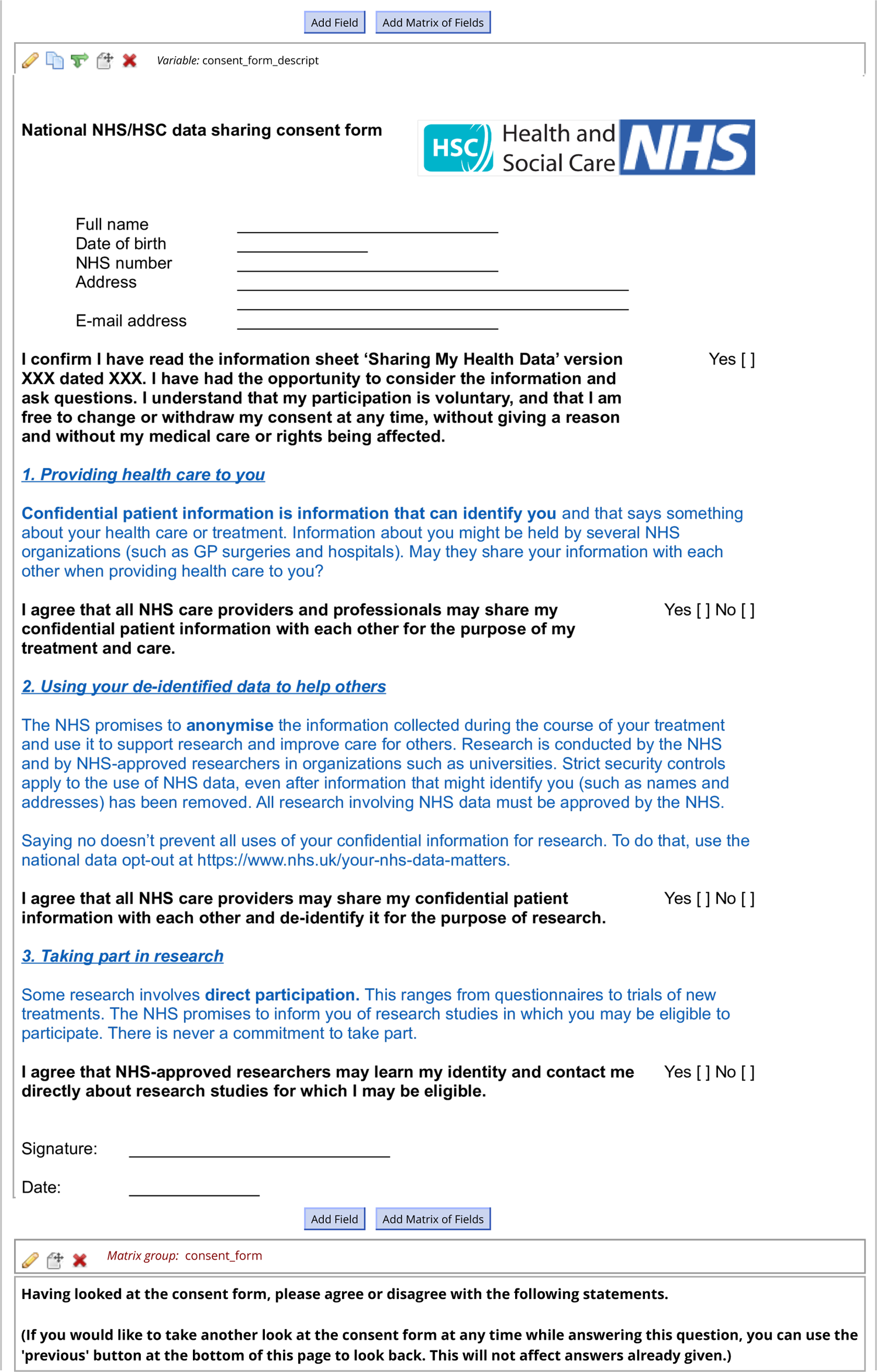
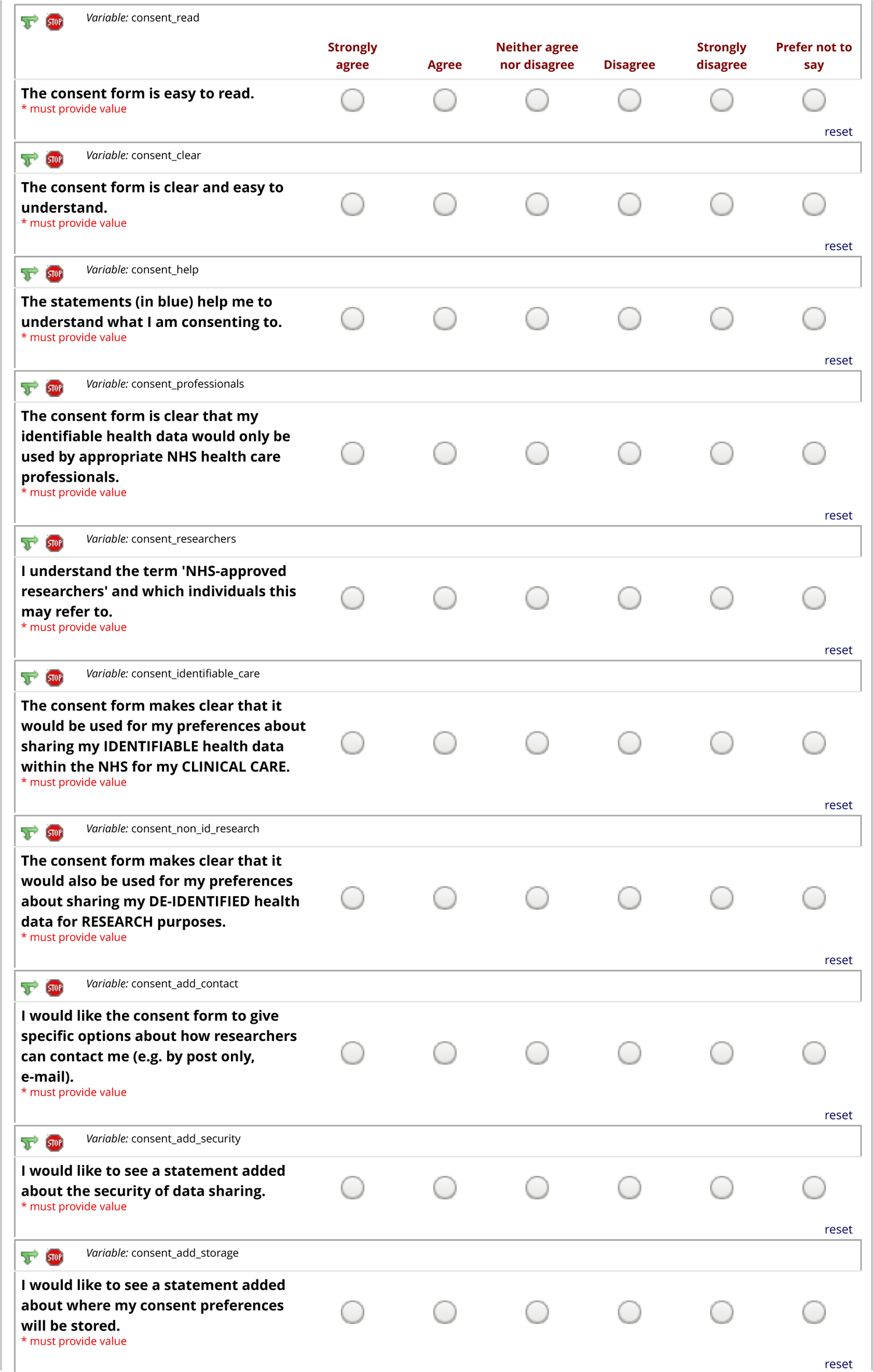
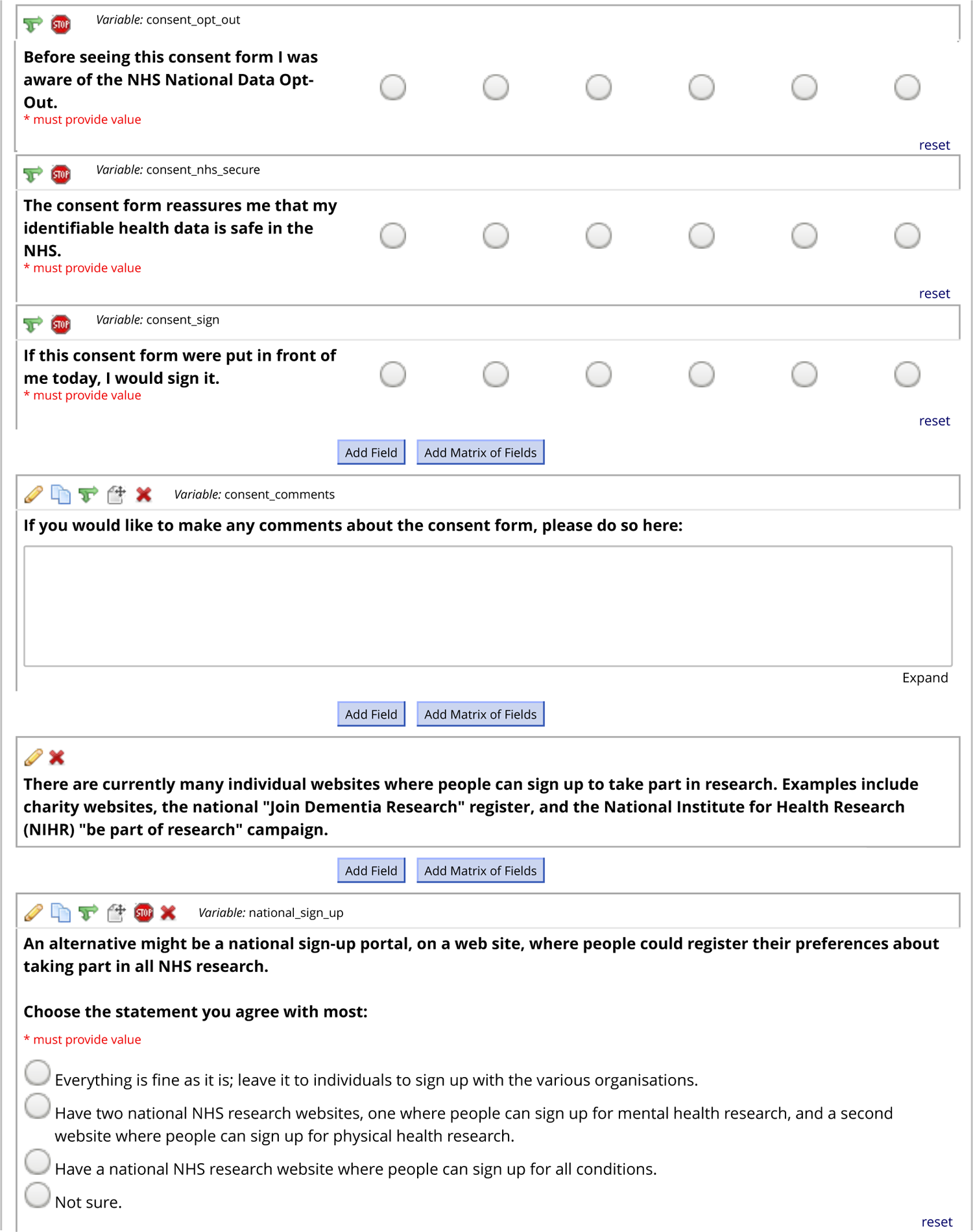
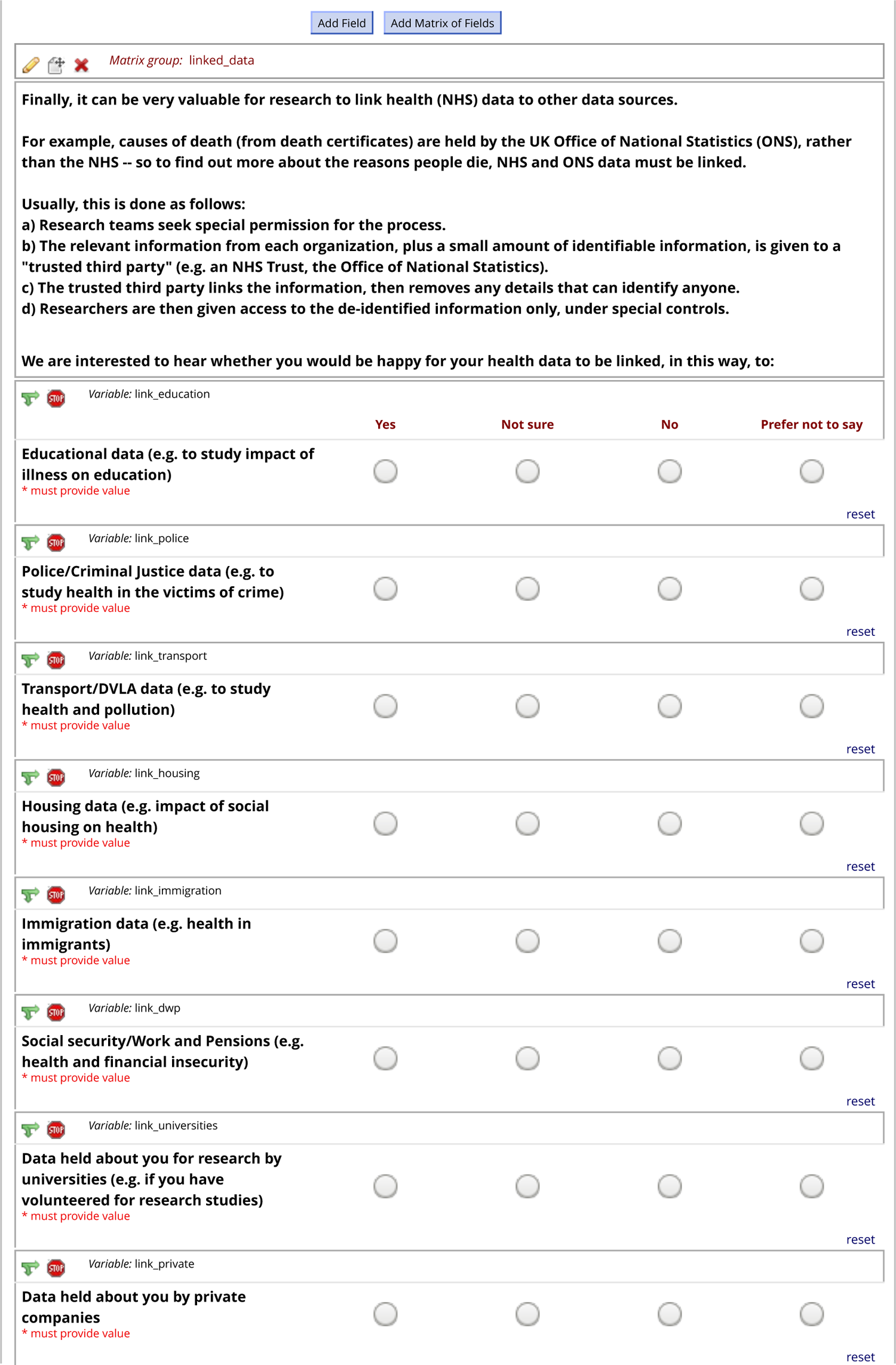
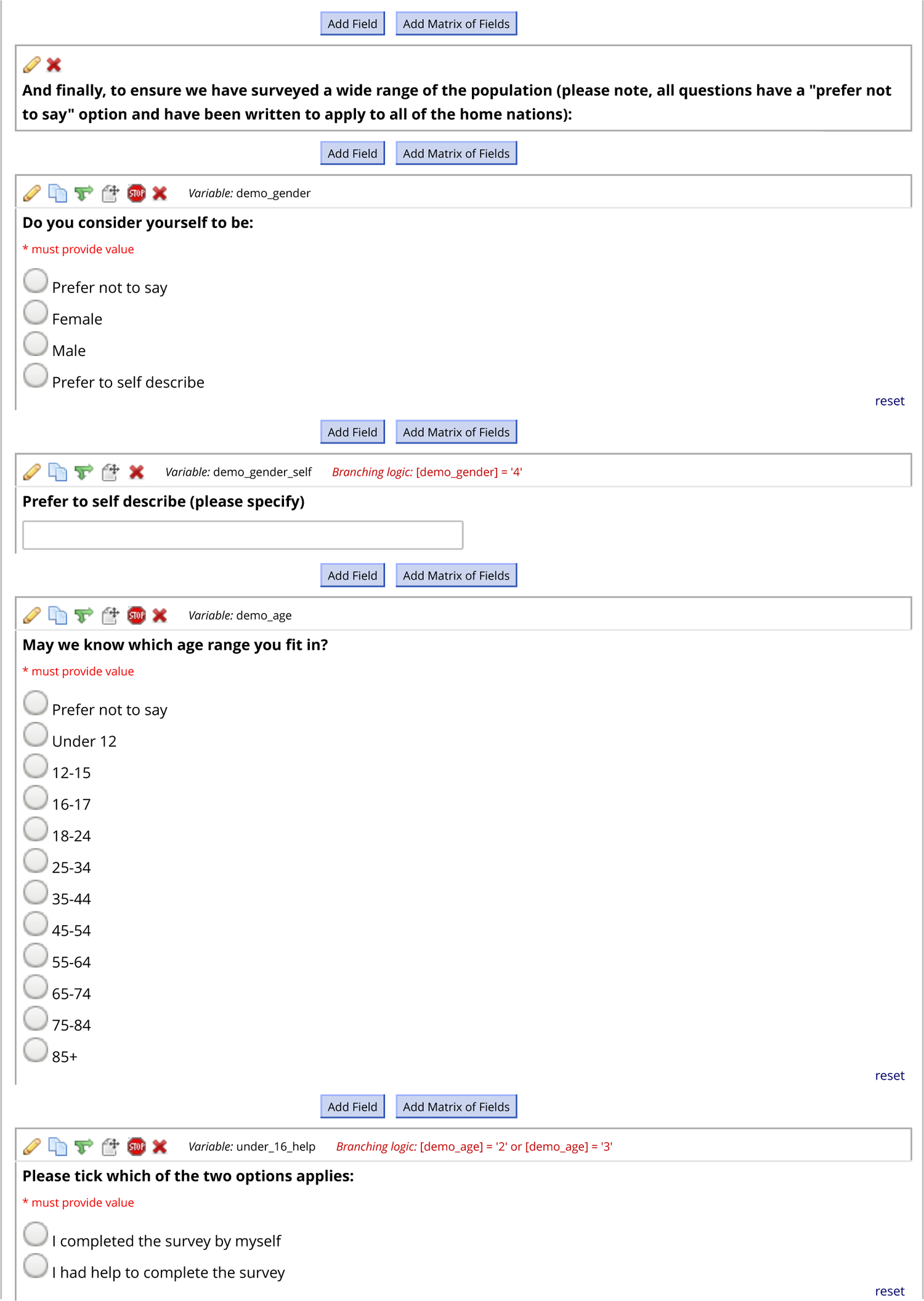
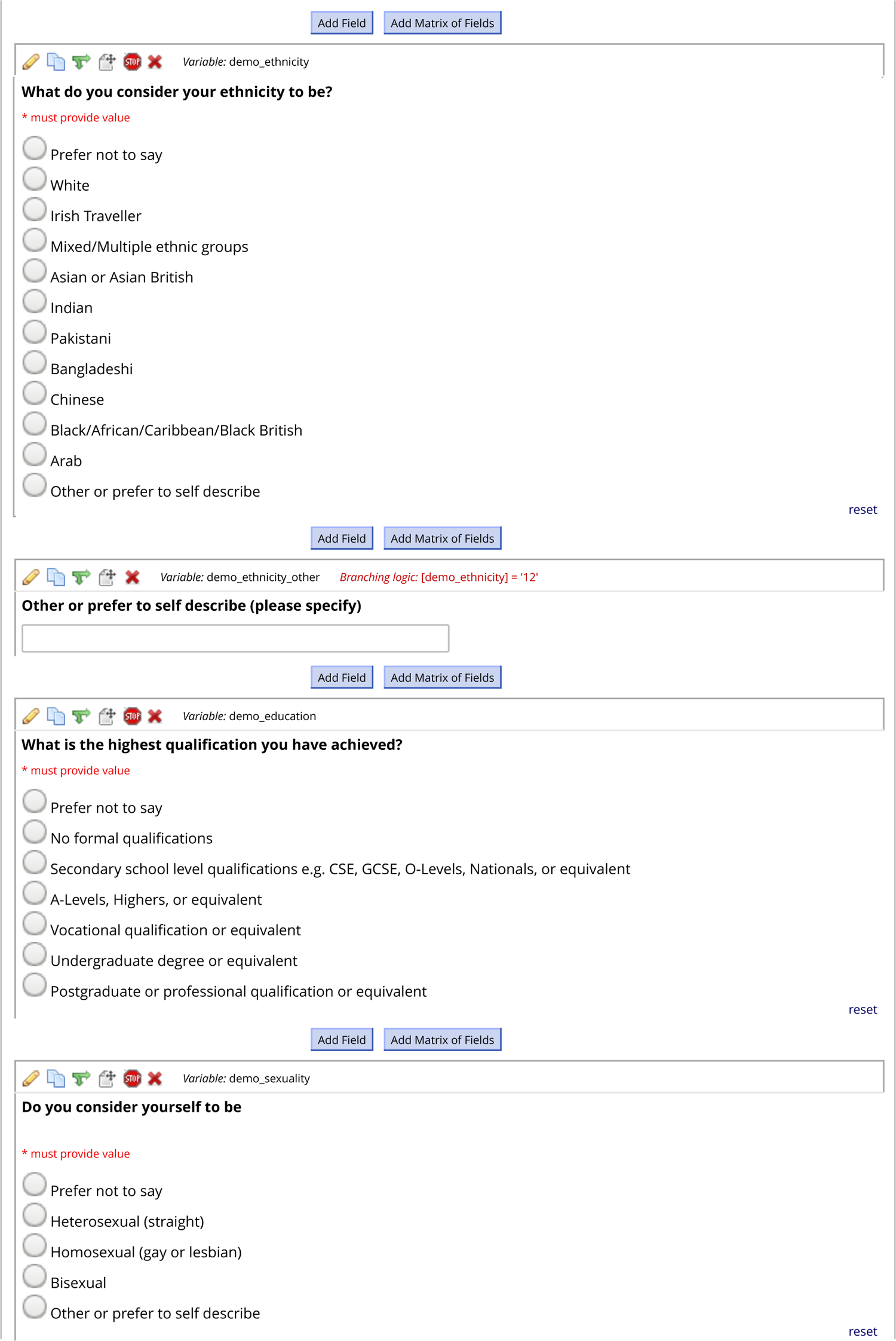
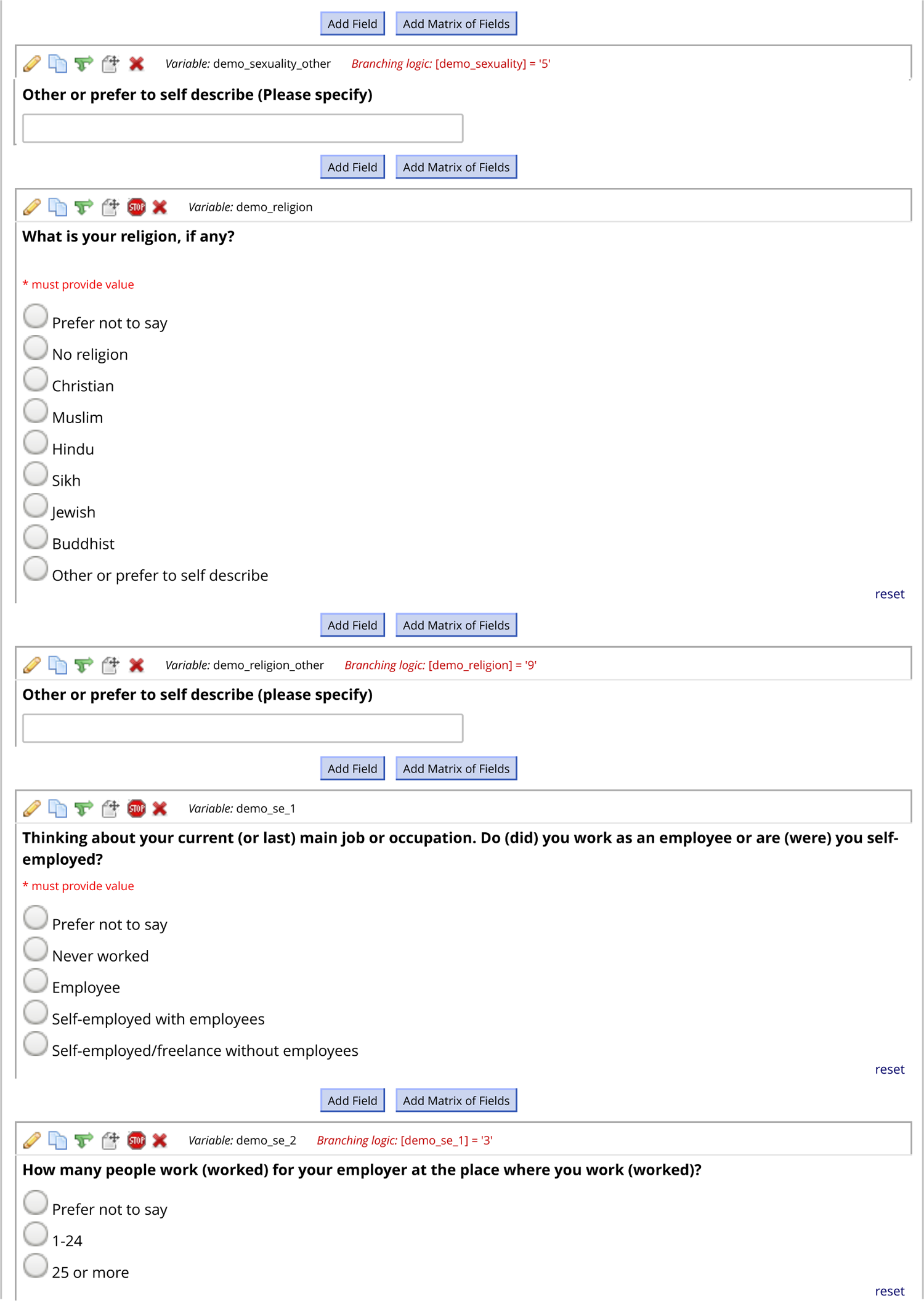
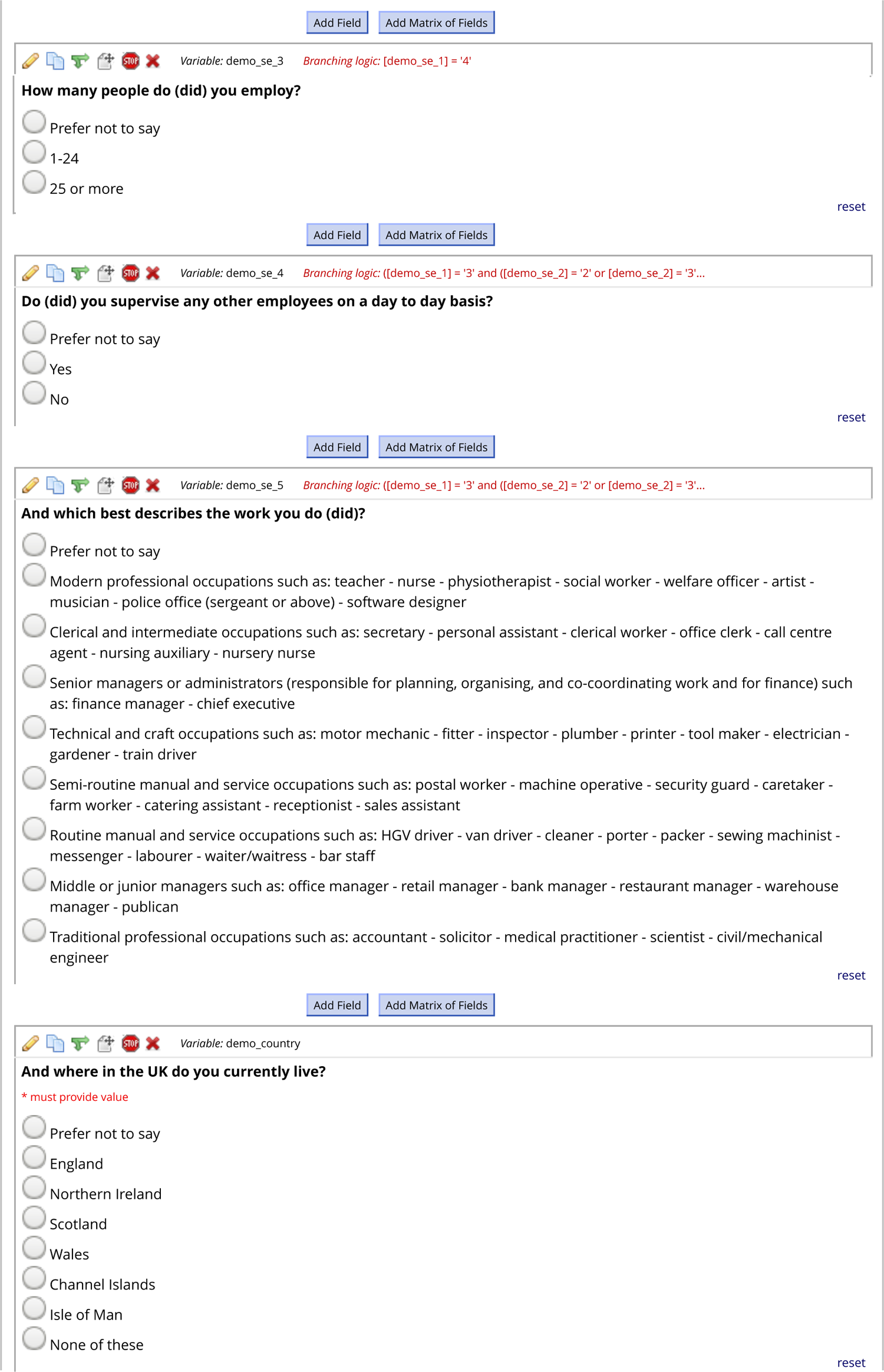
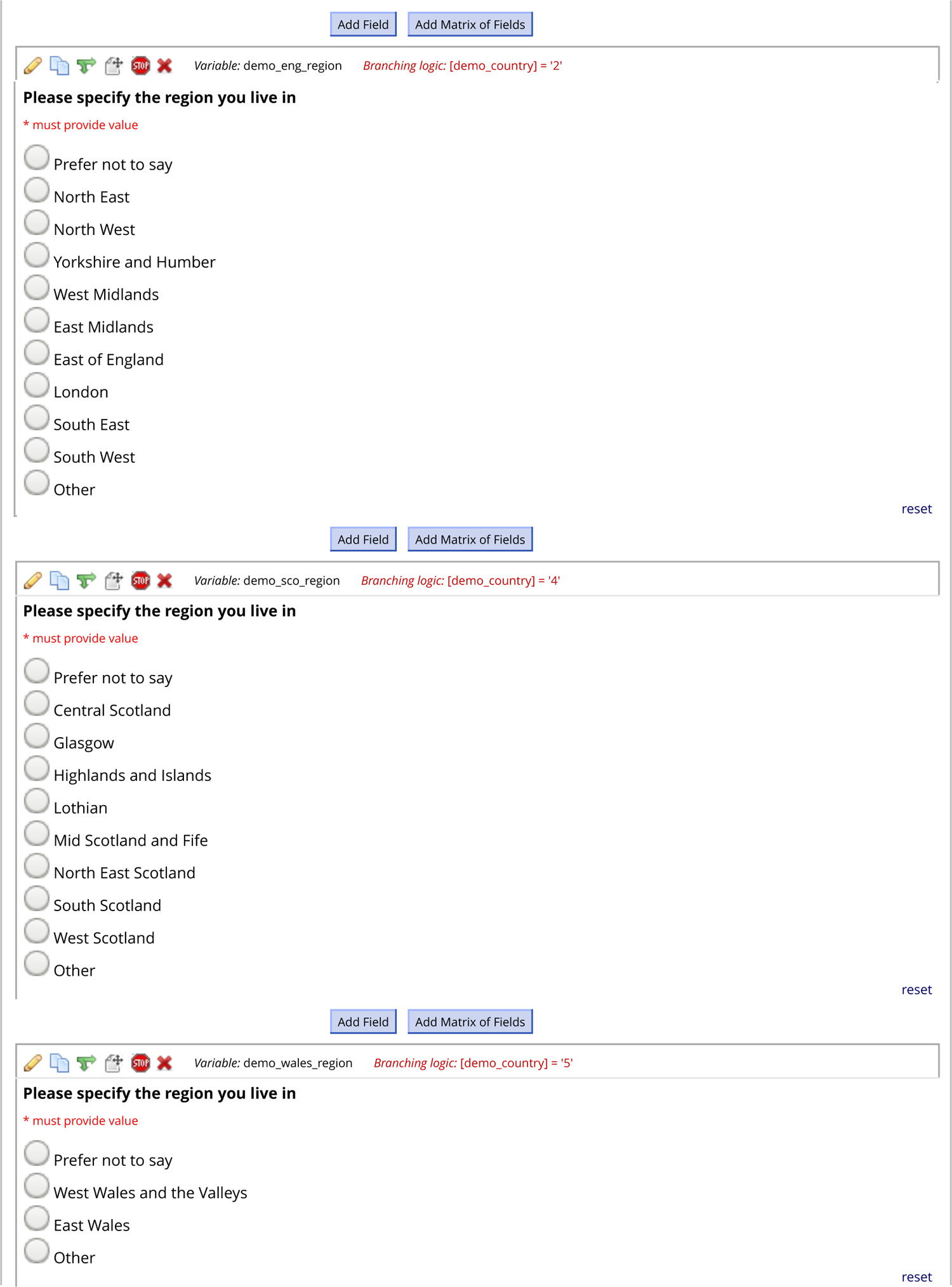
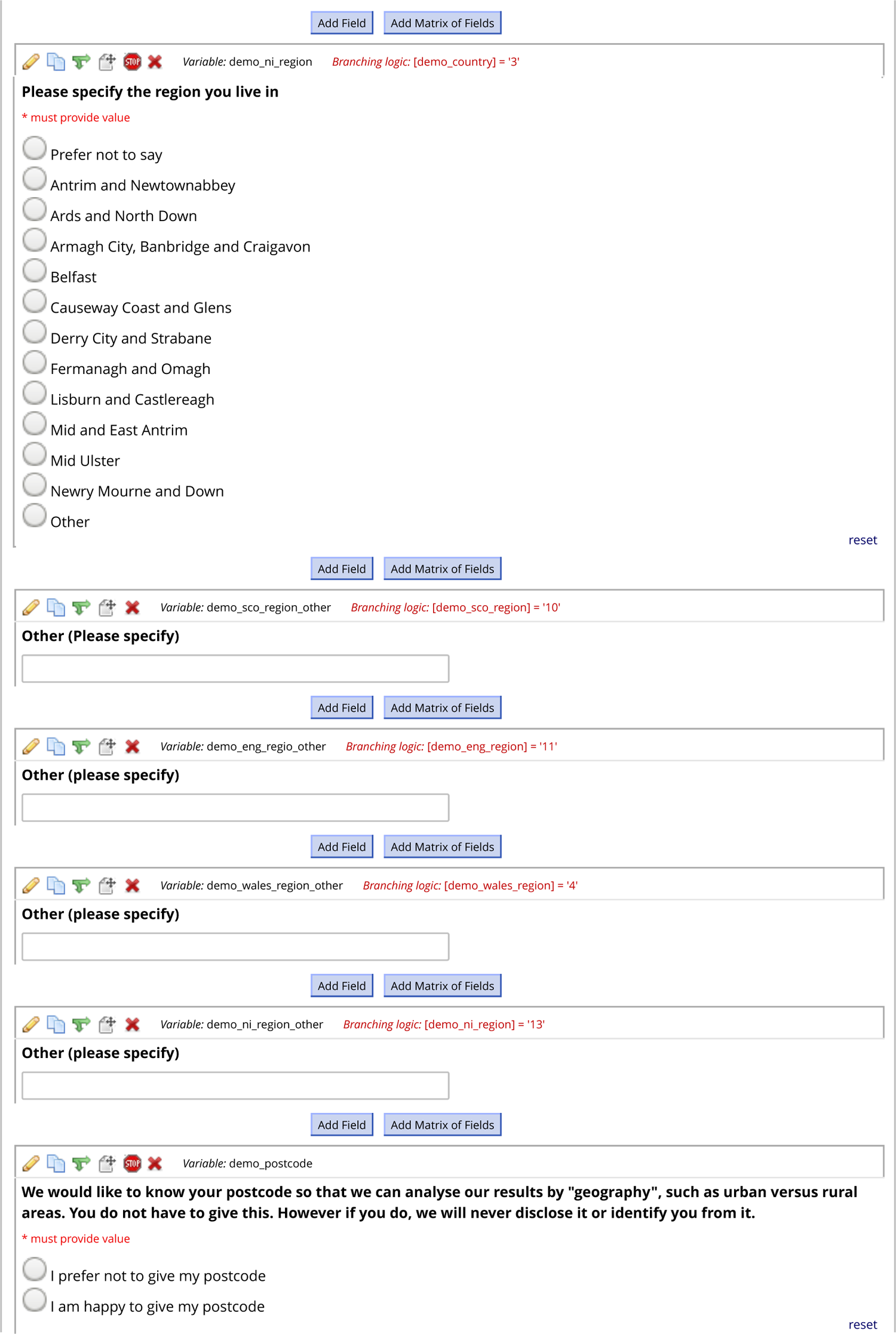
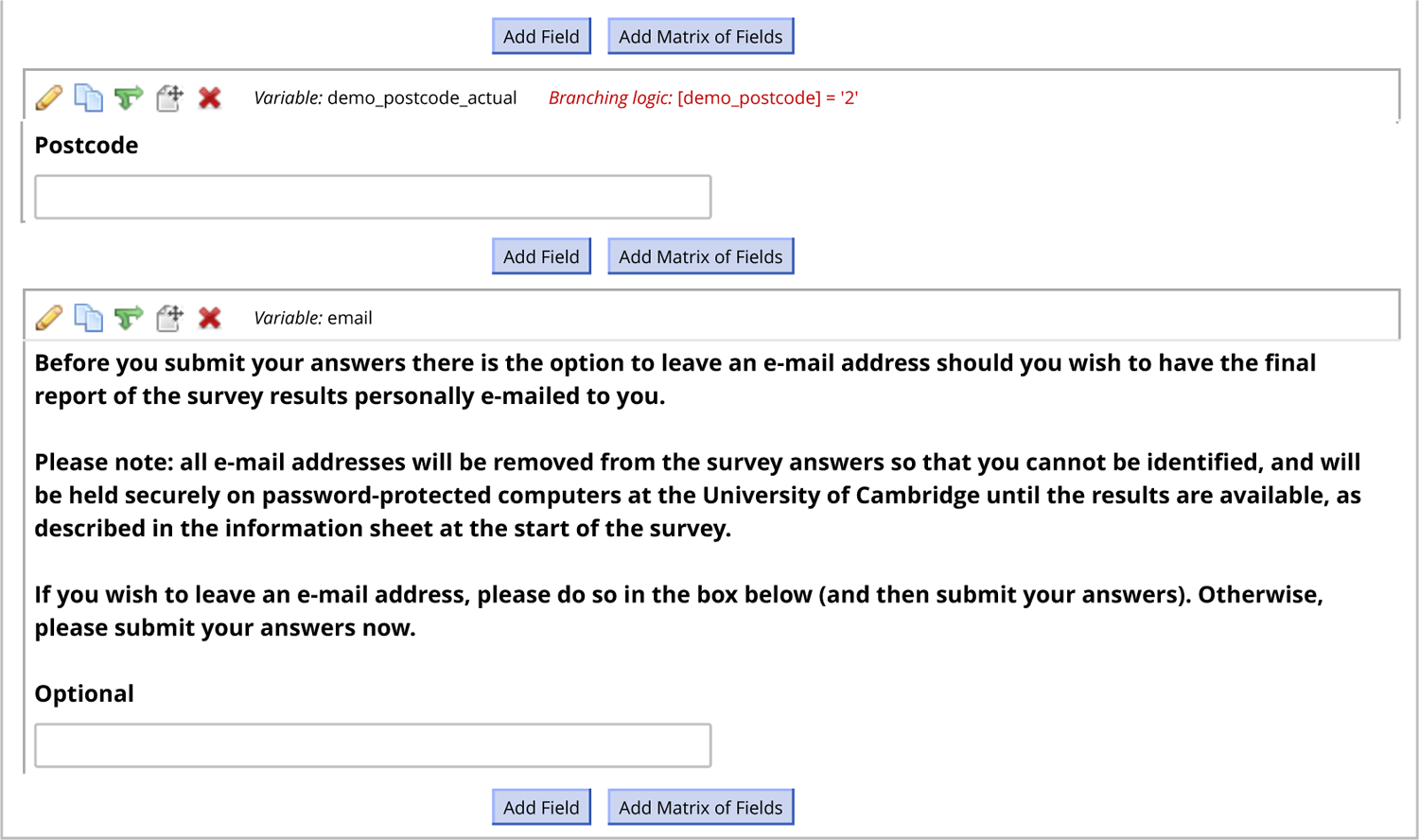
University of Cambridge NHS Health Data Consent Survey

### Appendix B Survey

**Appendix B** shows resulting screenshots of the survey running in a web browser. (Questions labelled 11, 12, and 13 internally, as per **Appendix A**, were the same question, but with differing accompanying framing statements; the “concern” framing is shown here. The last content page is shown twice, once in its starting state and once fully expanded after partial completion with hypothetical data.)

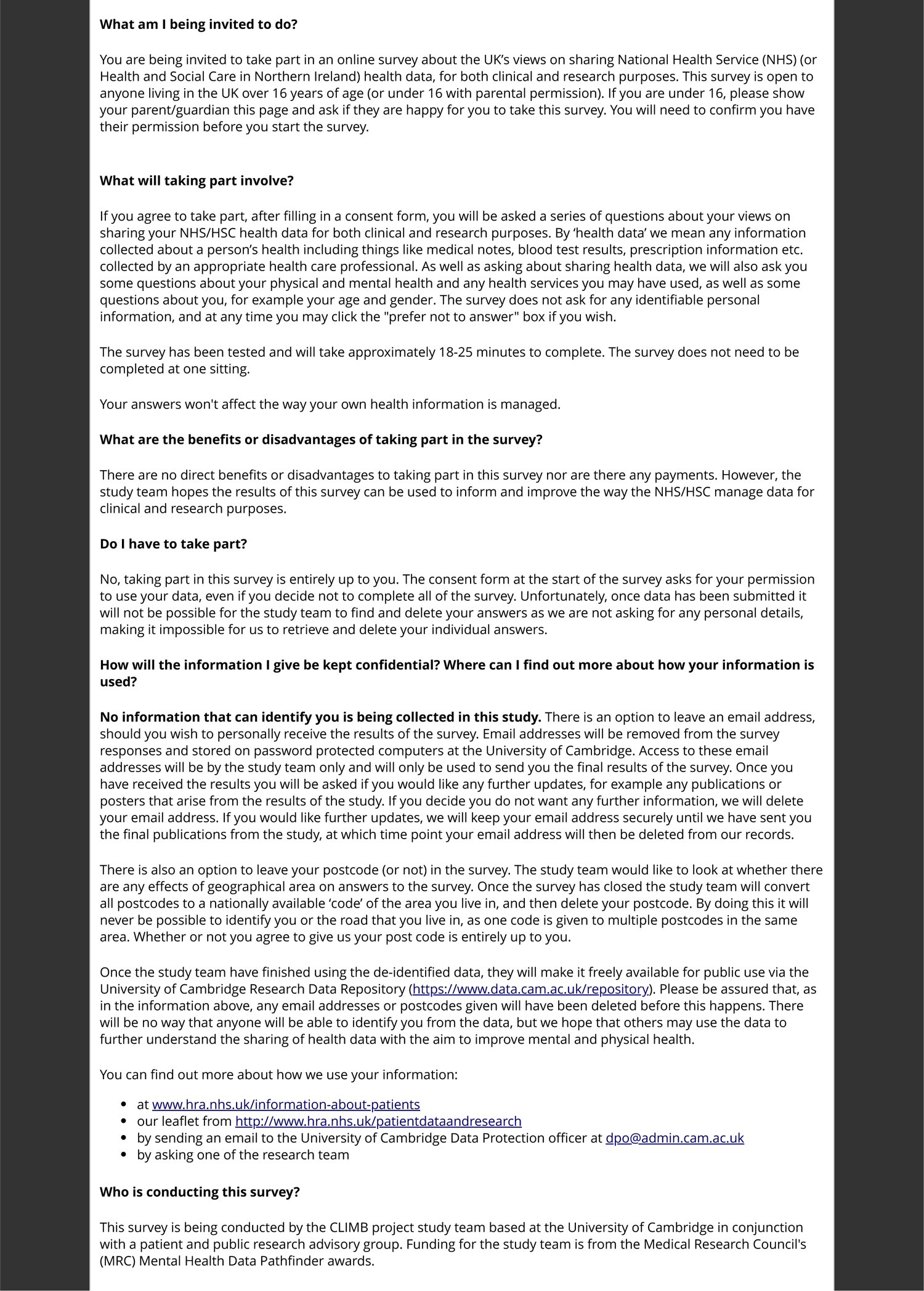
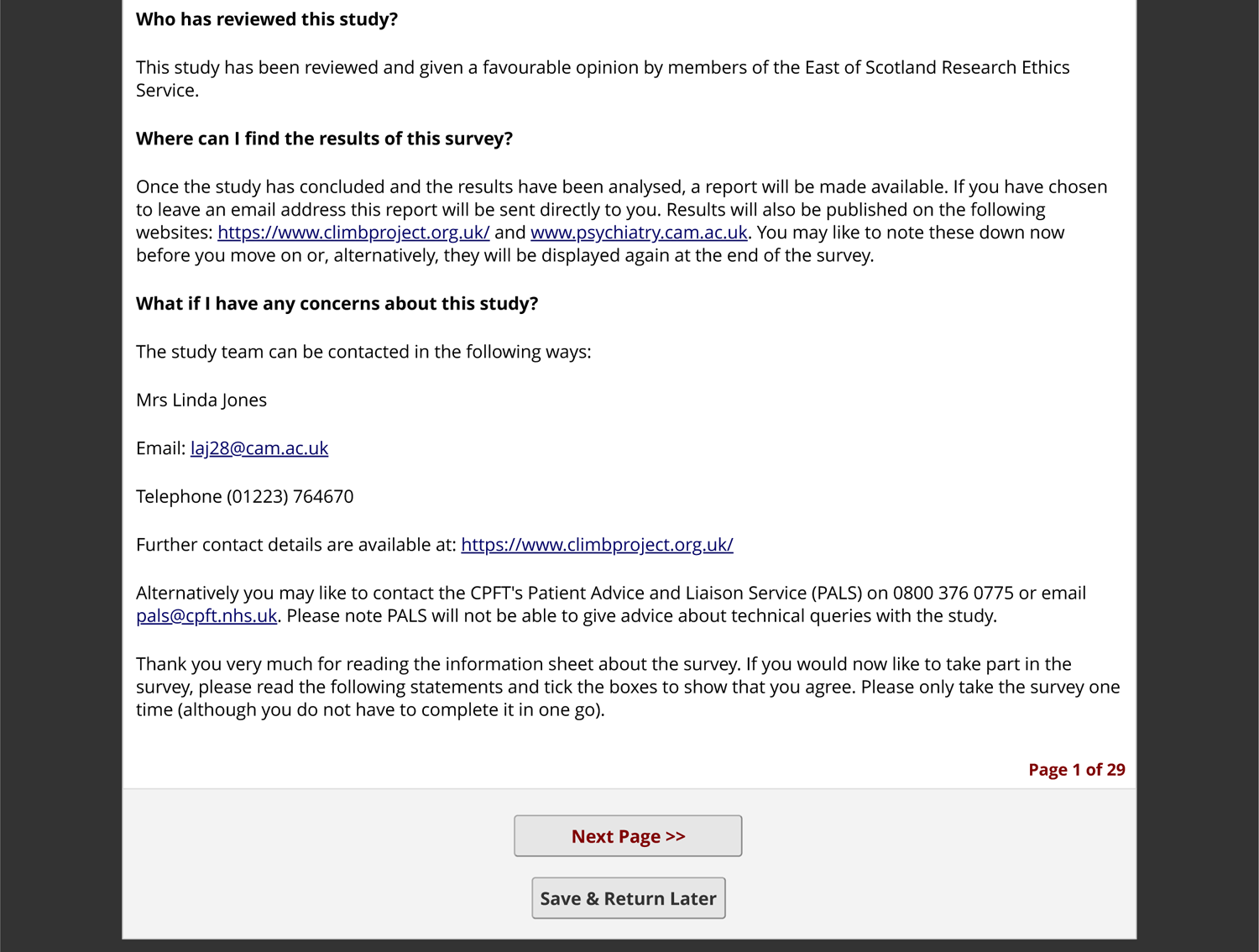
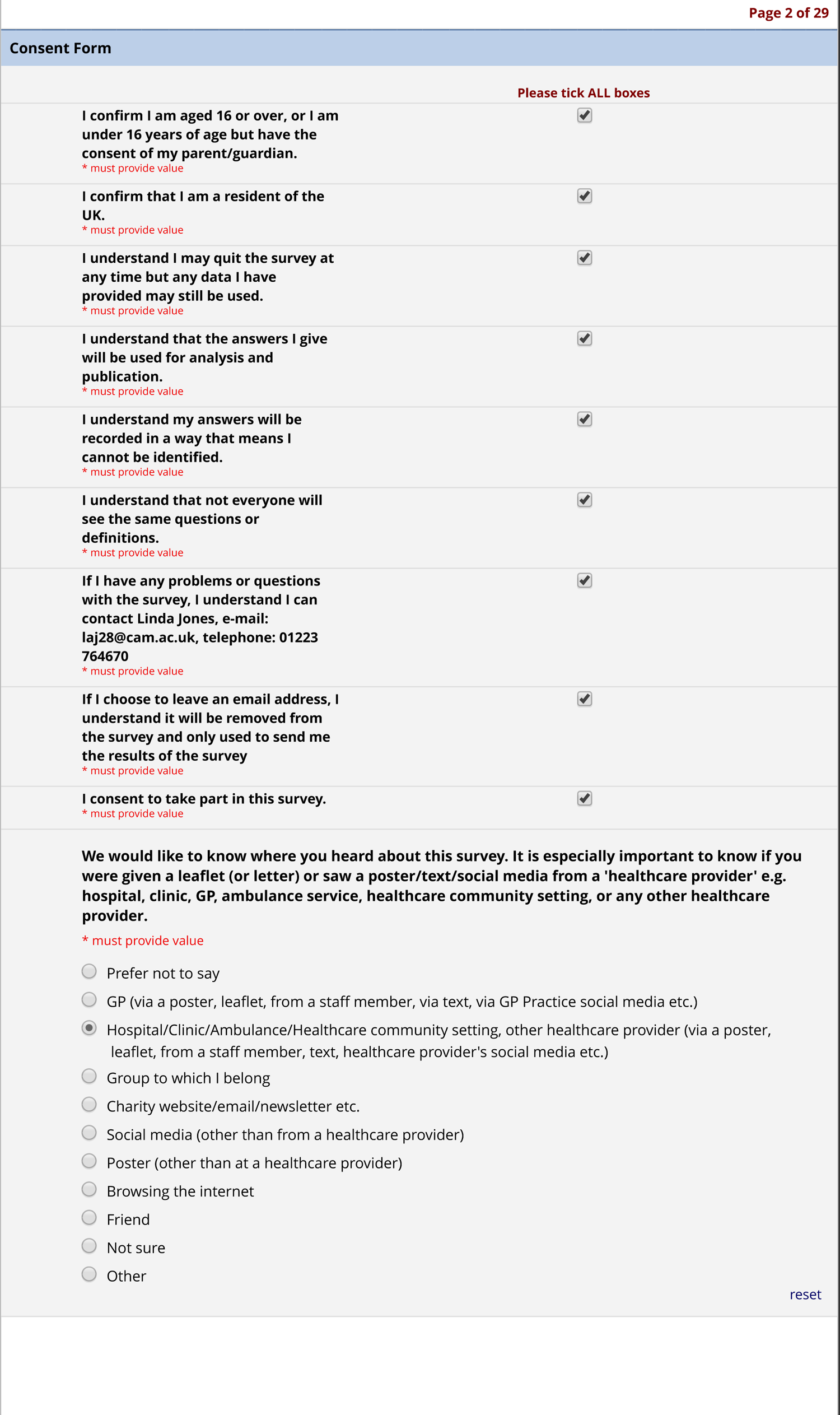
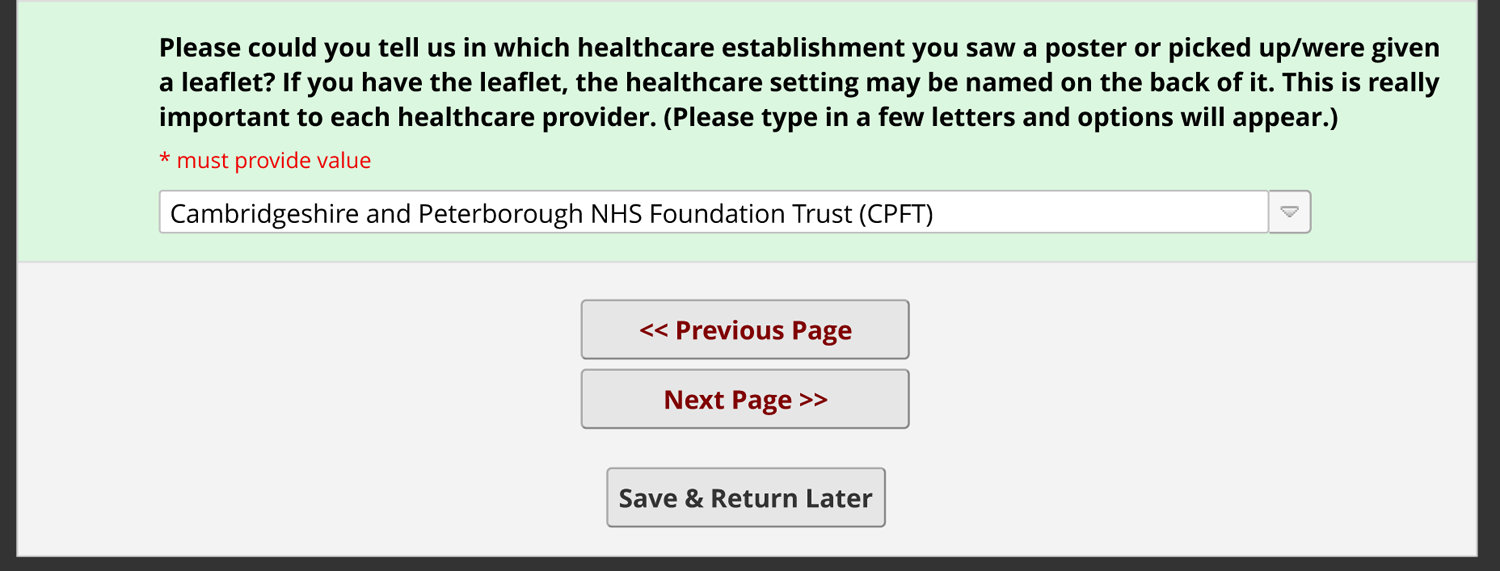
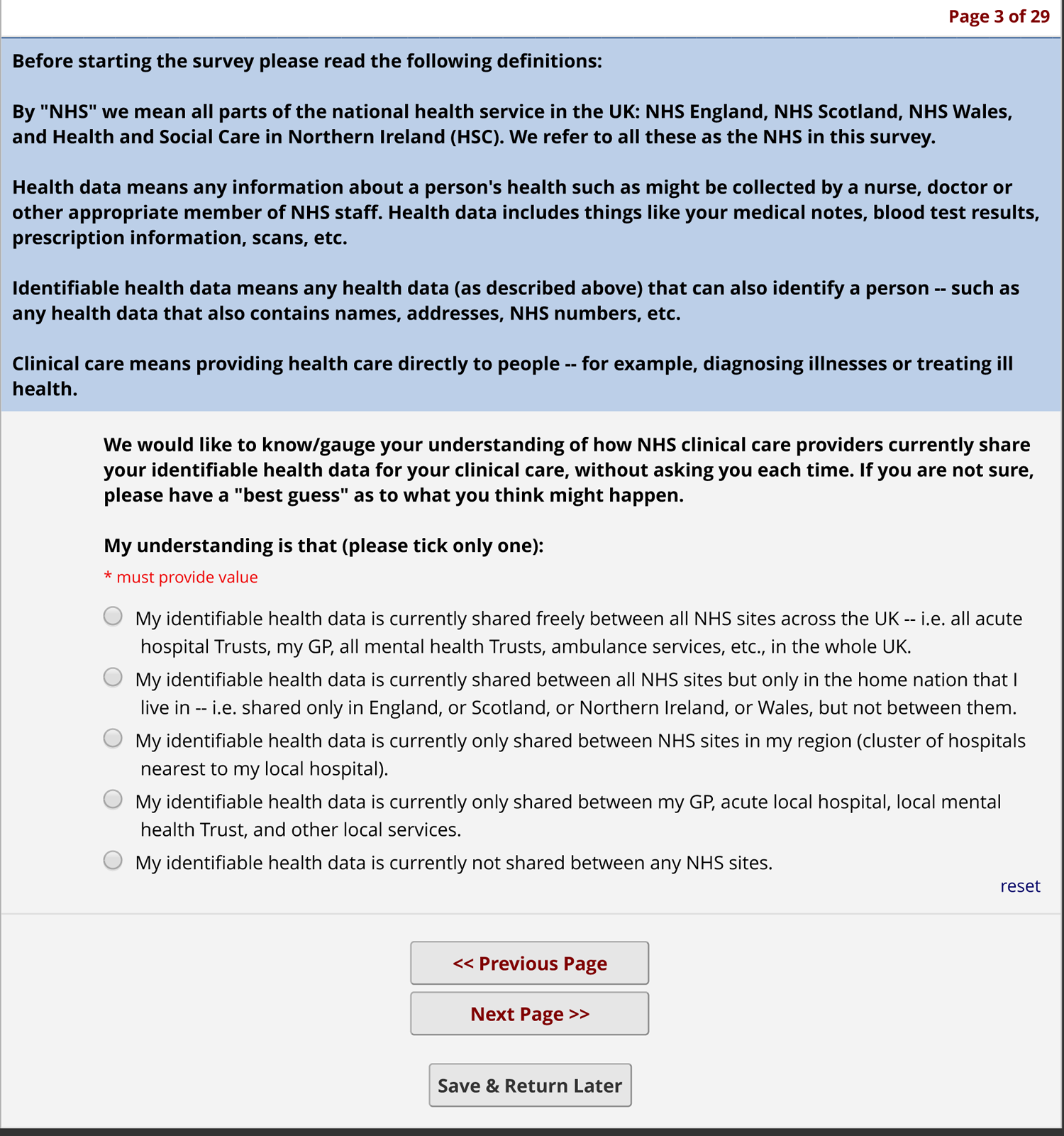
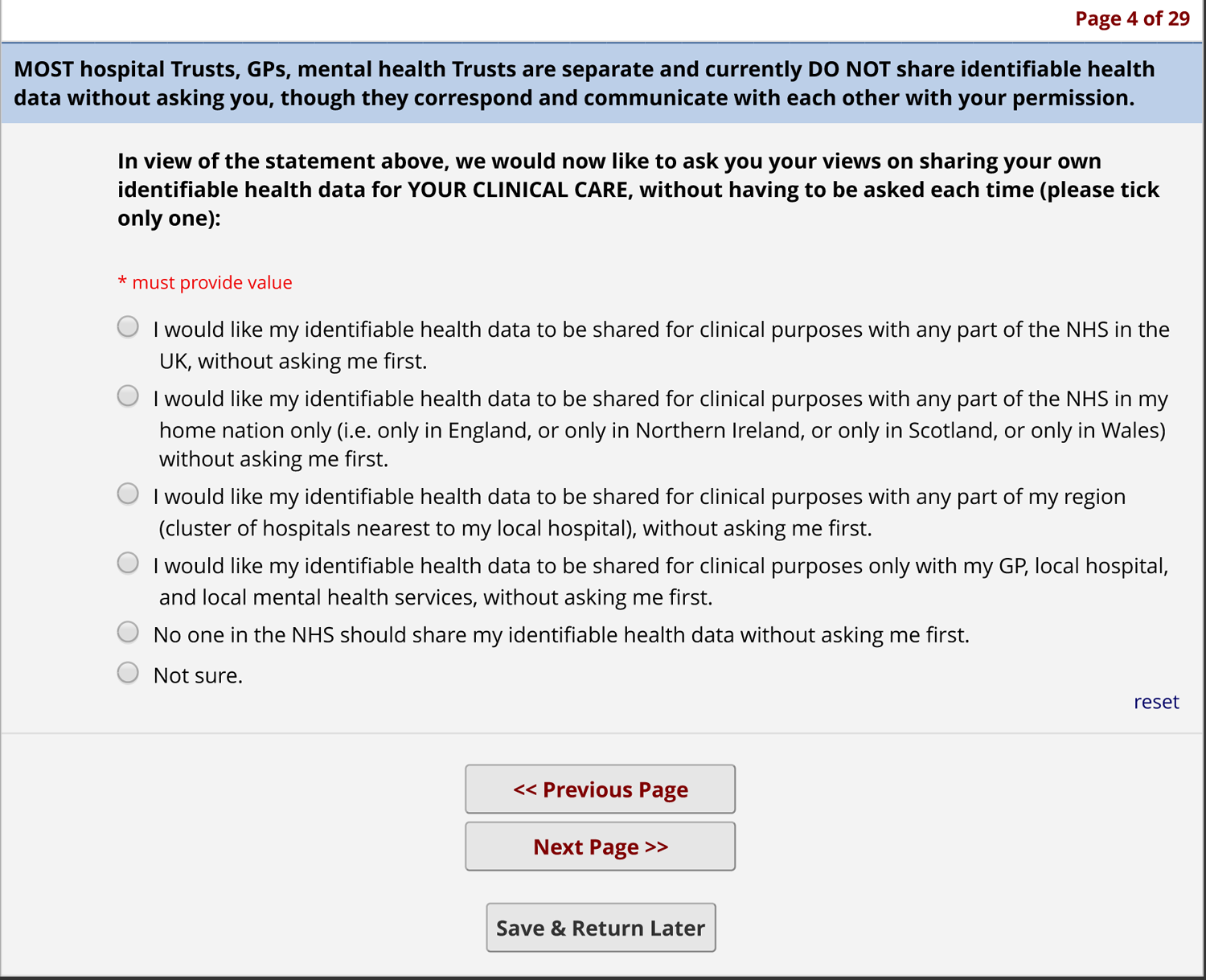
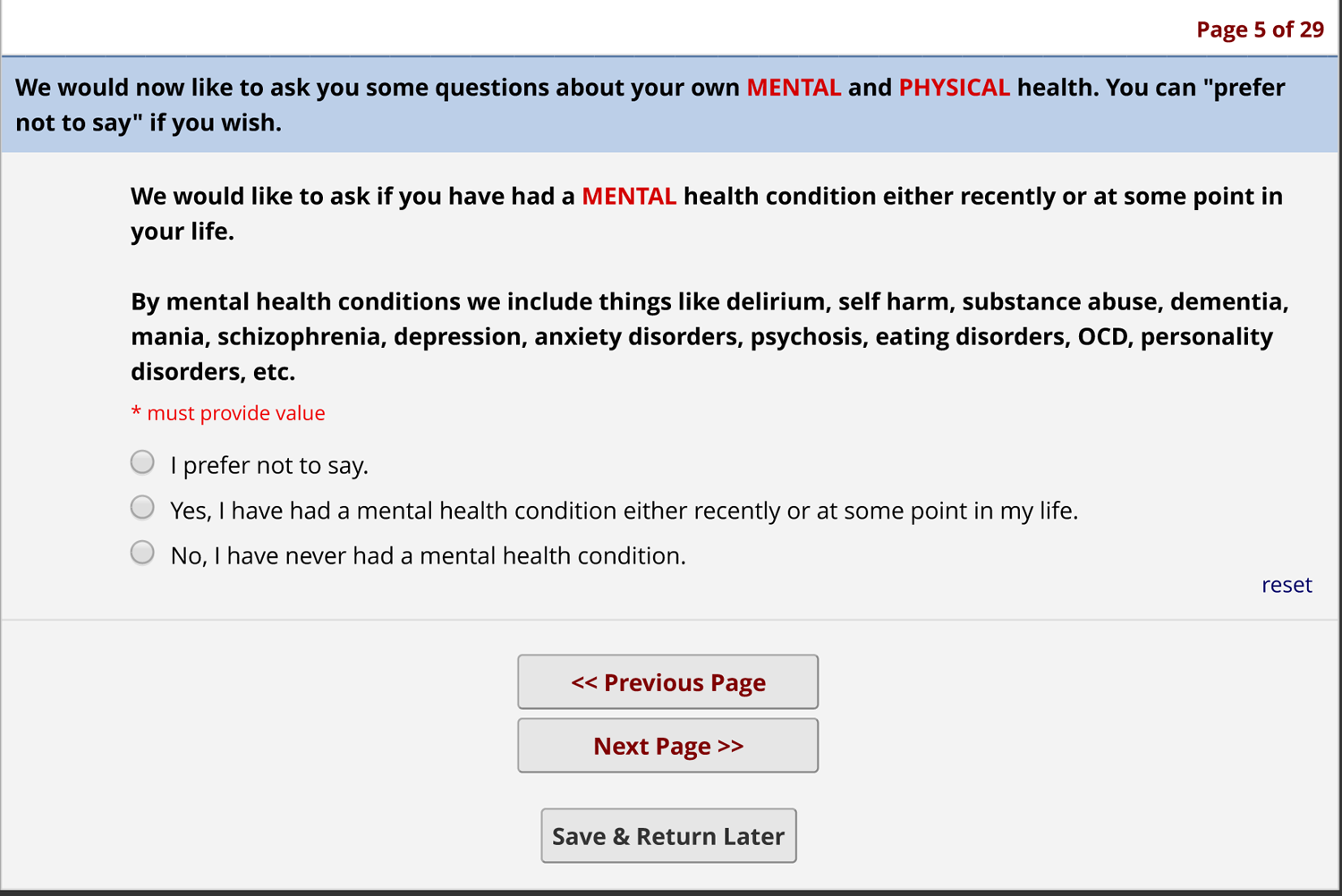
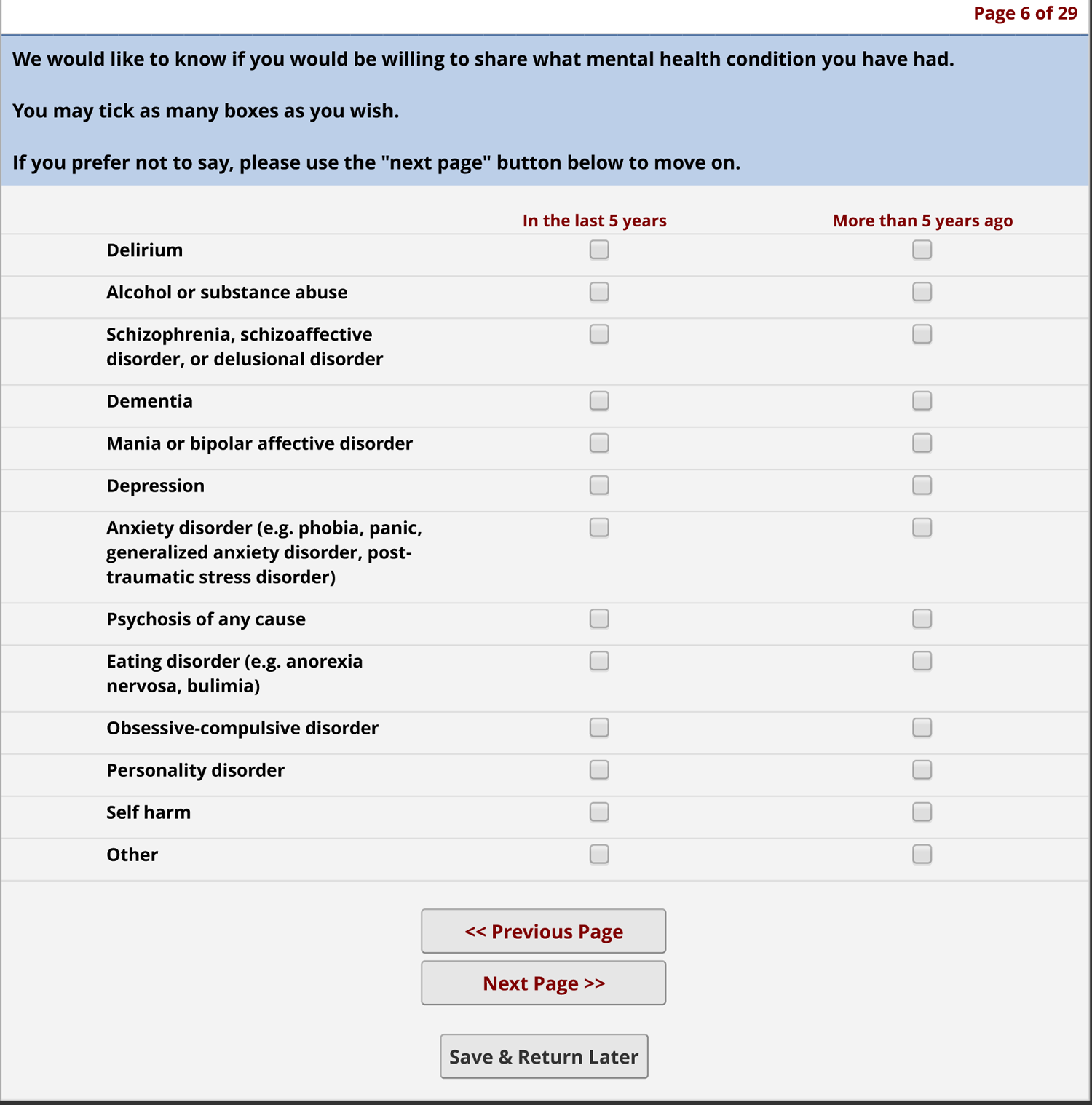
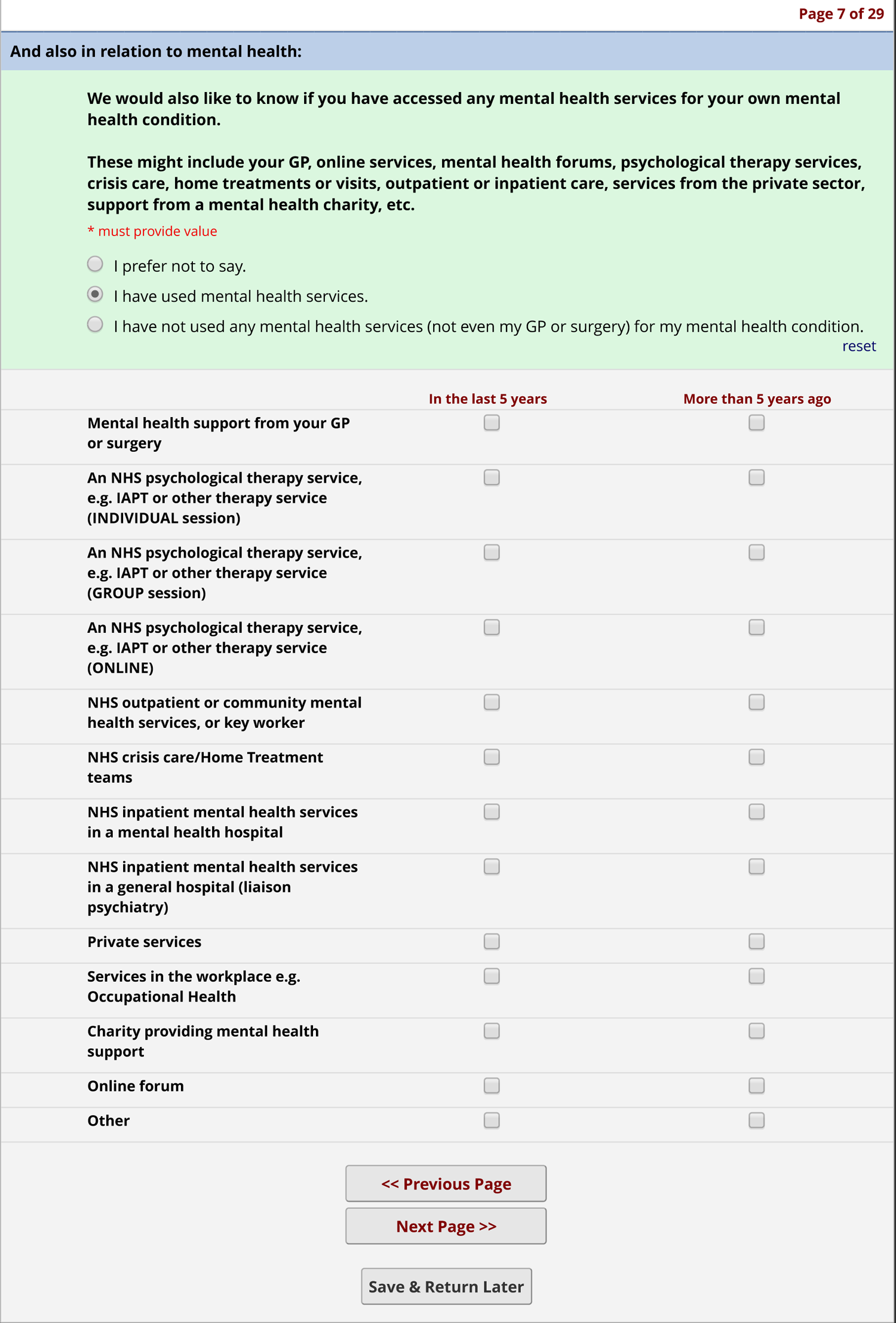
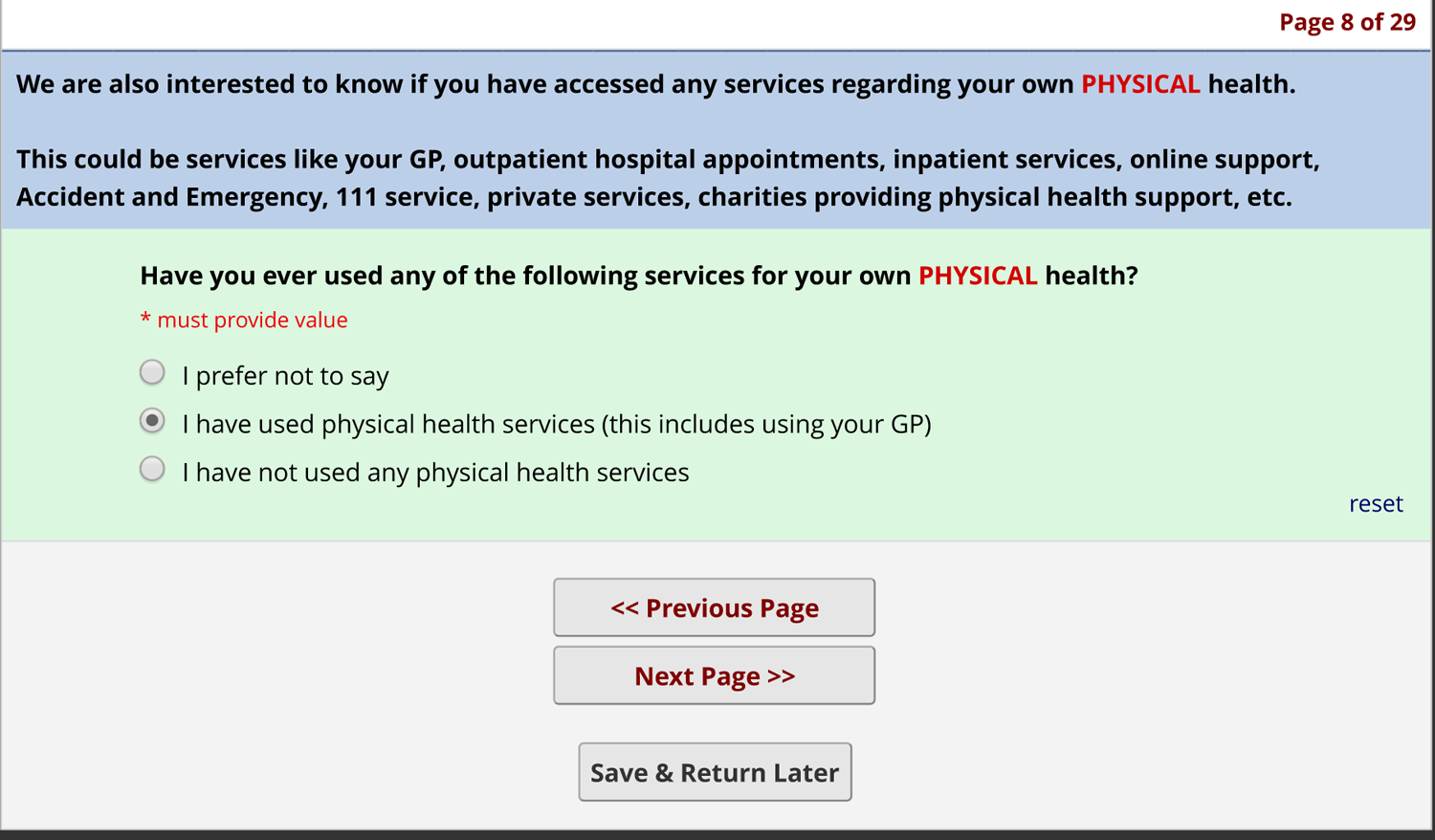
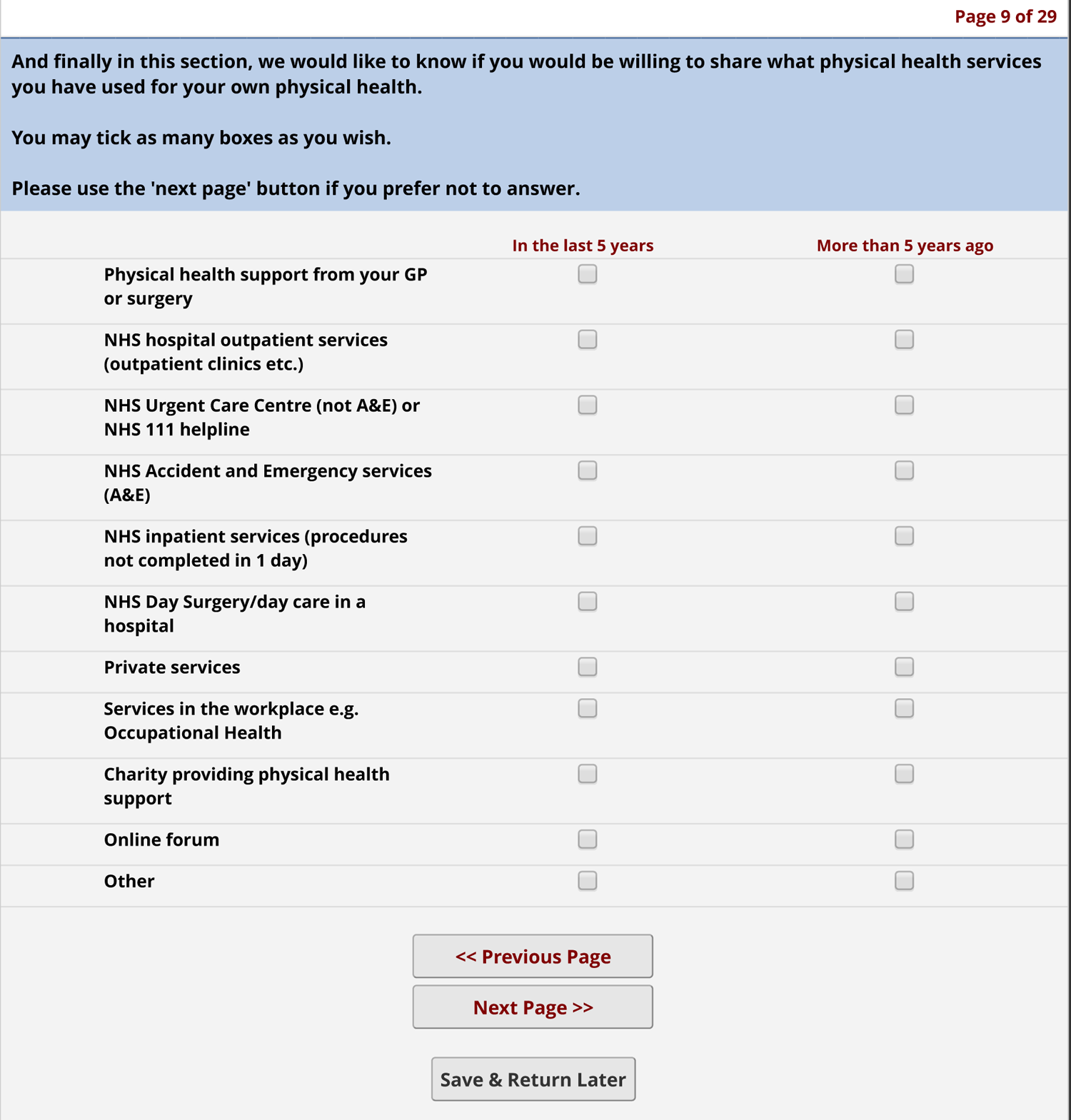
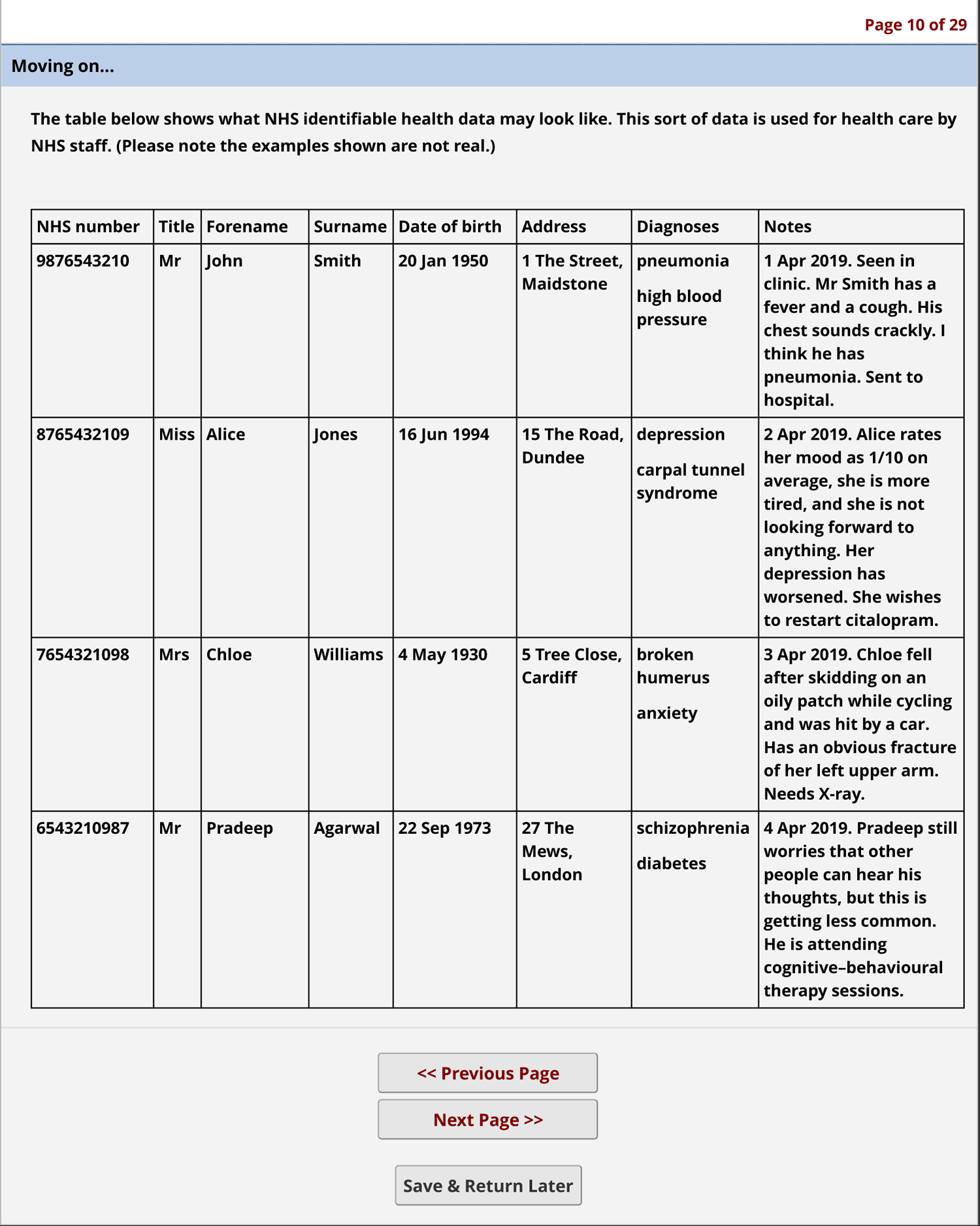
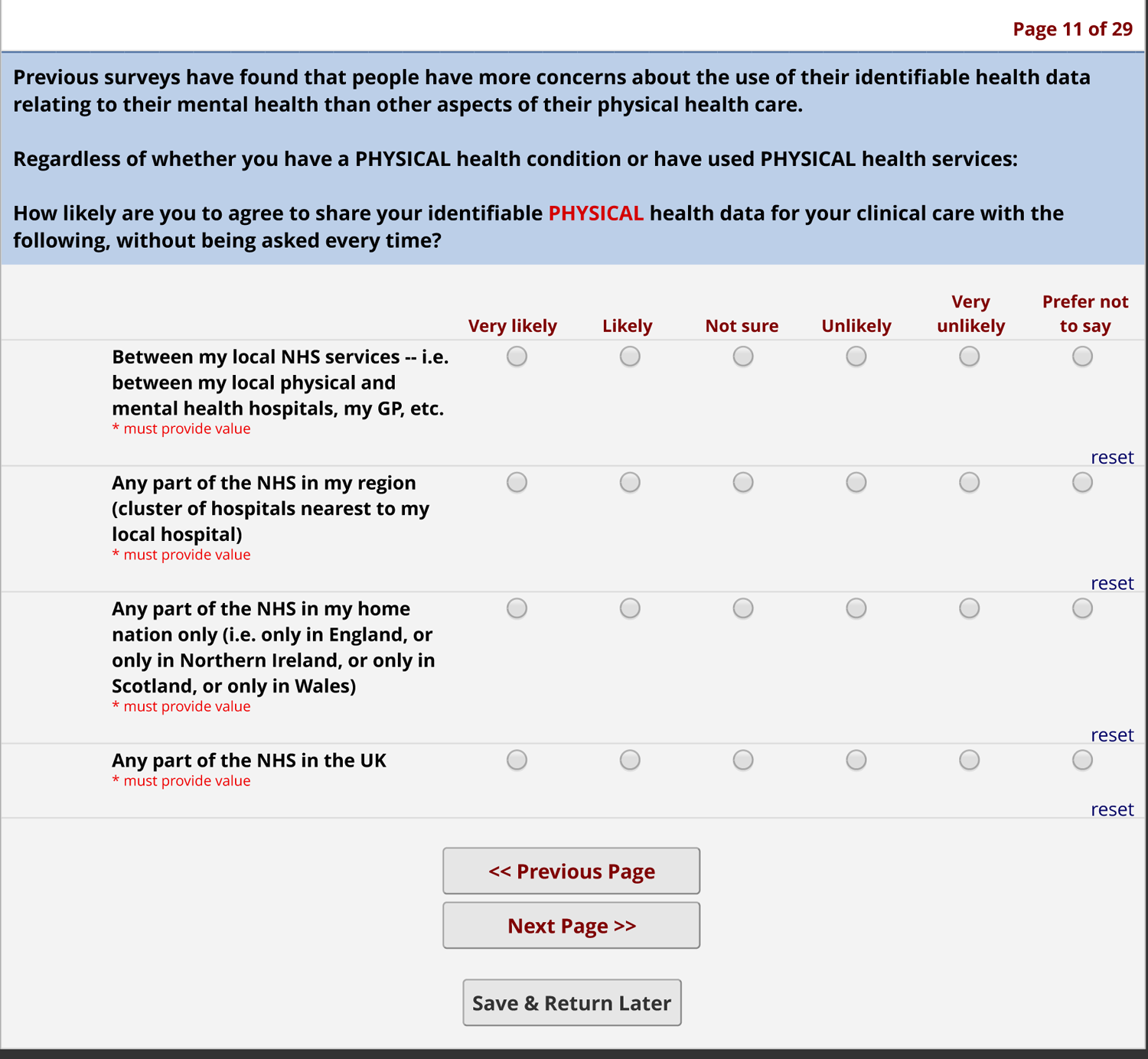
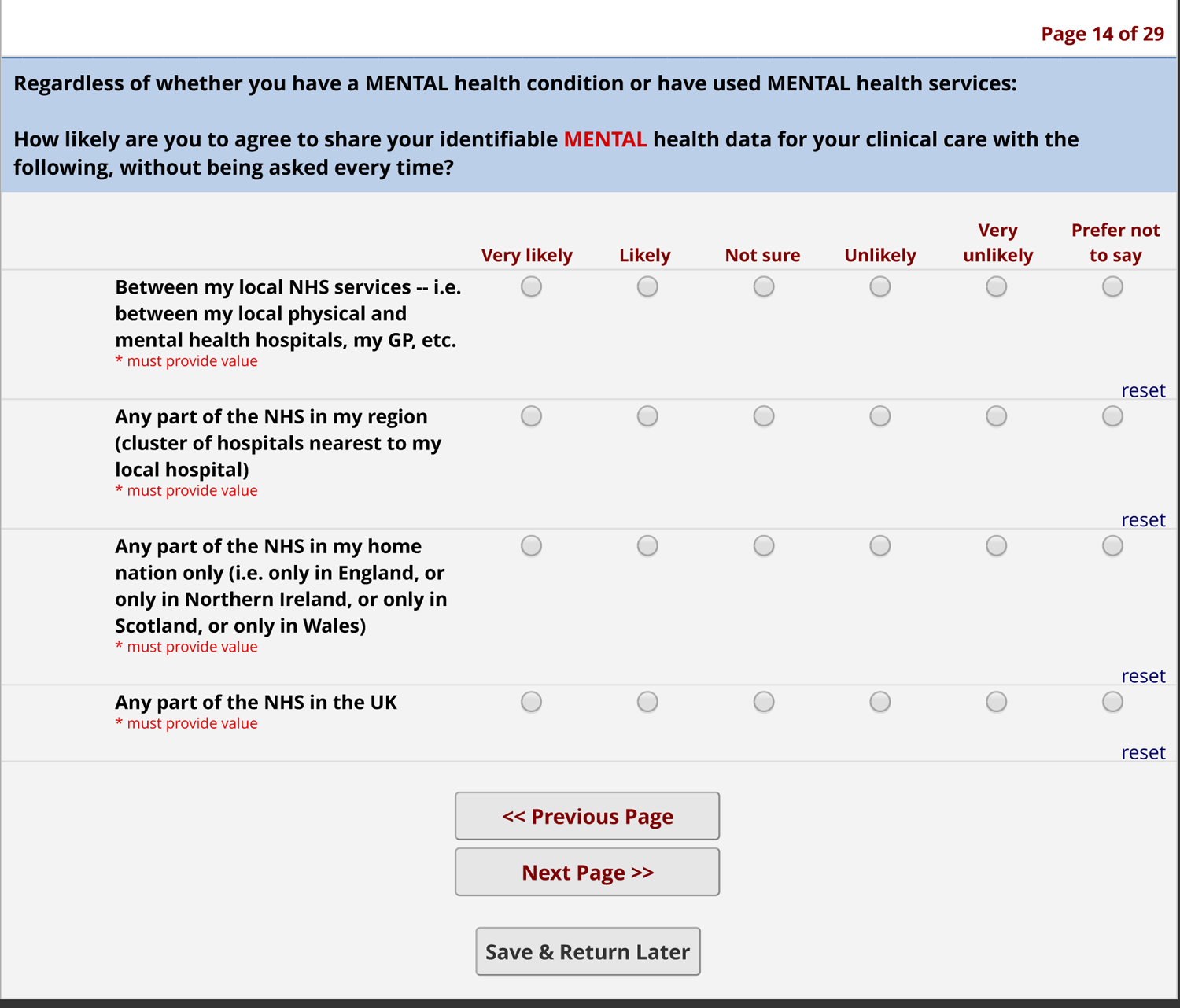
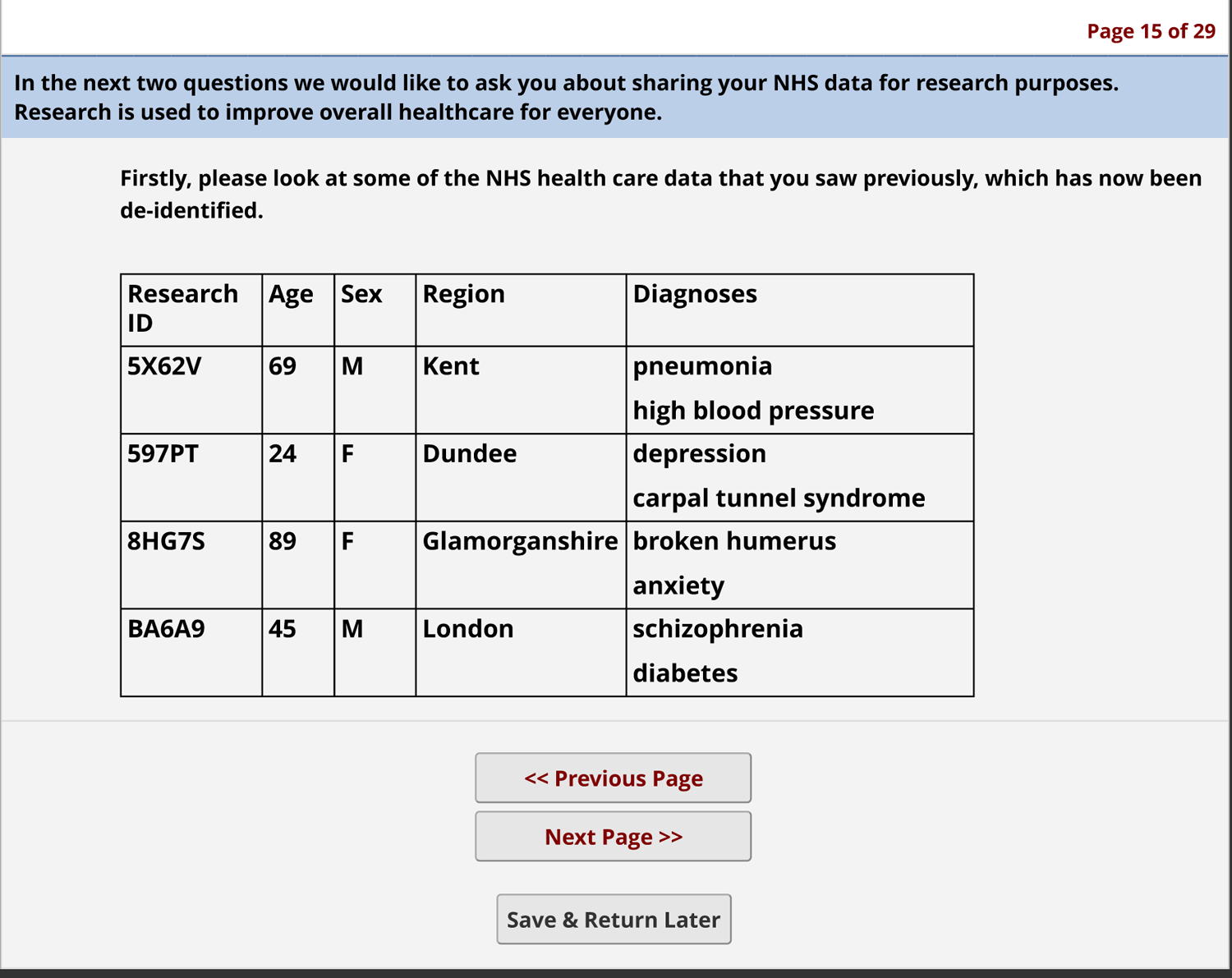
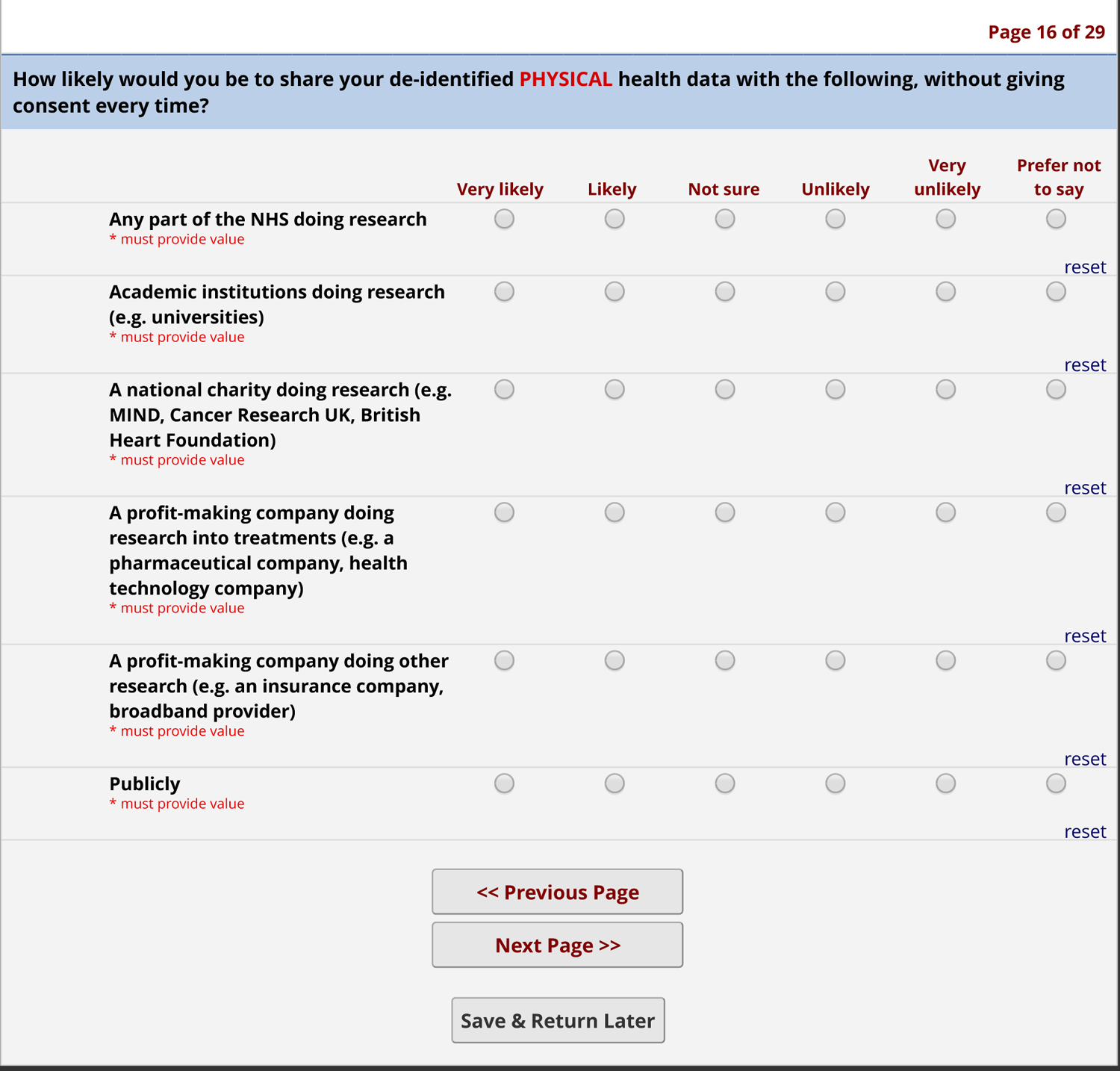
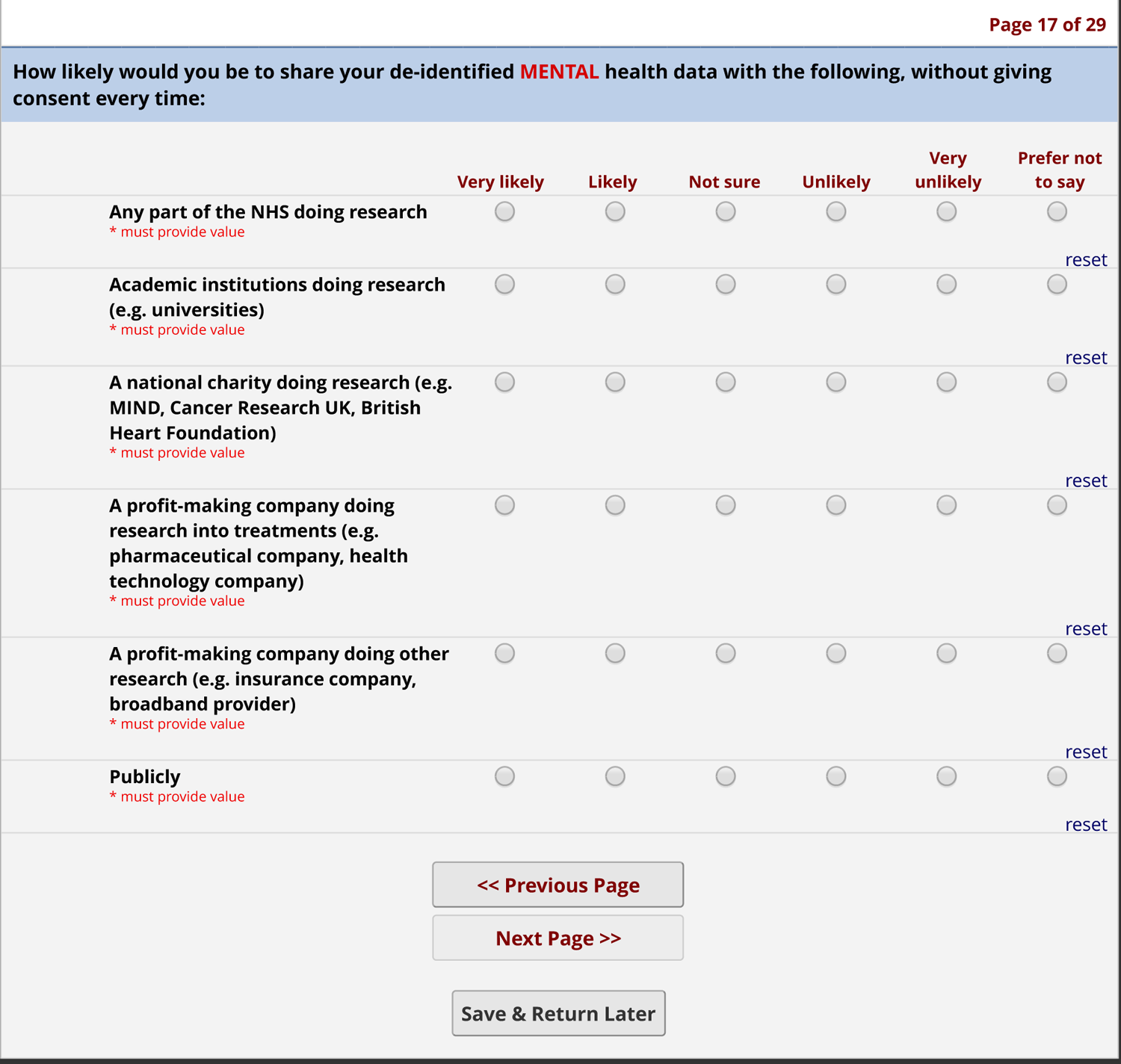
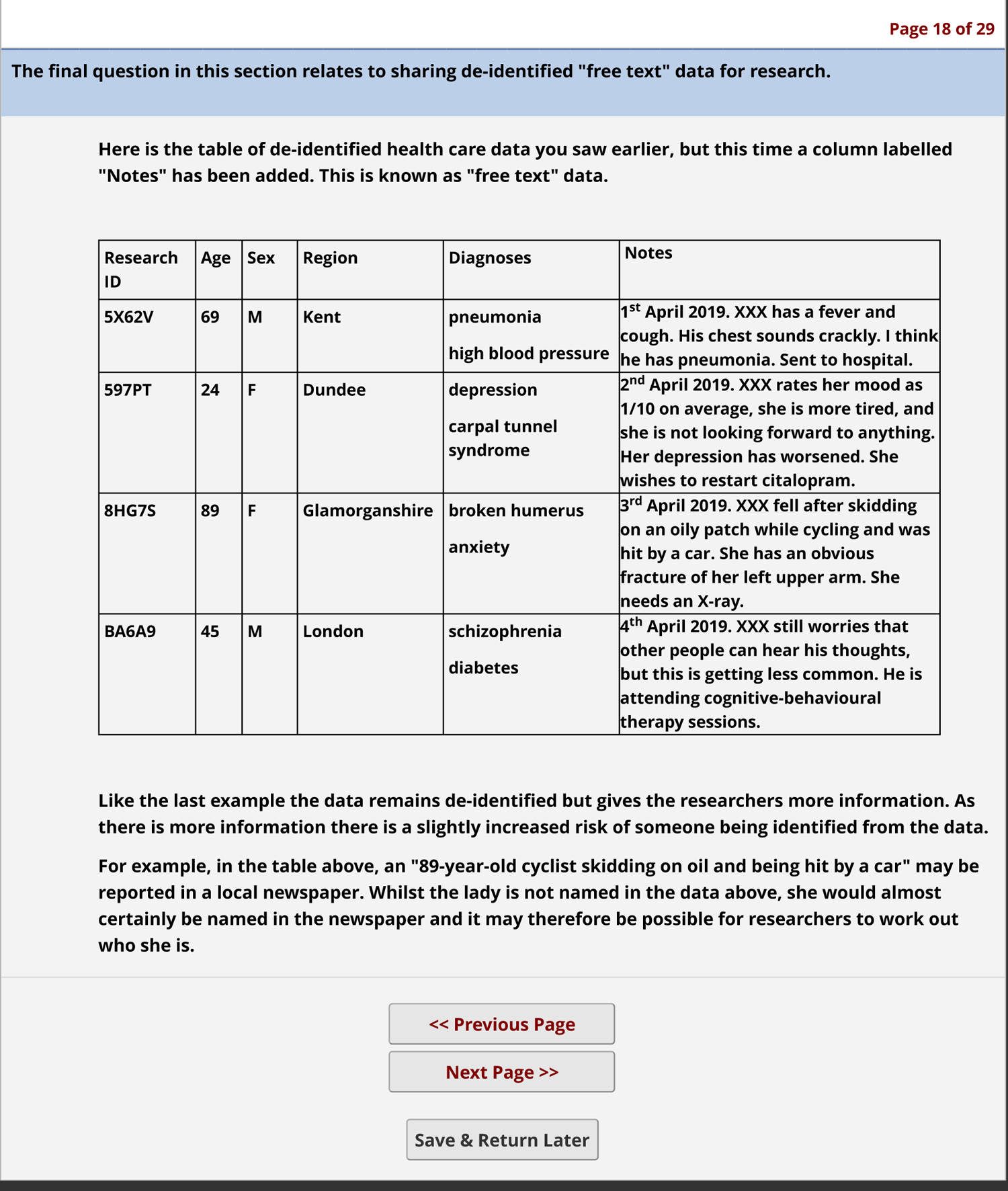
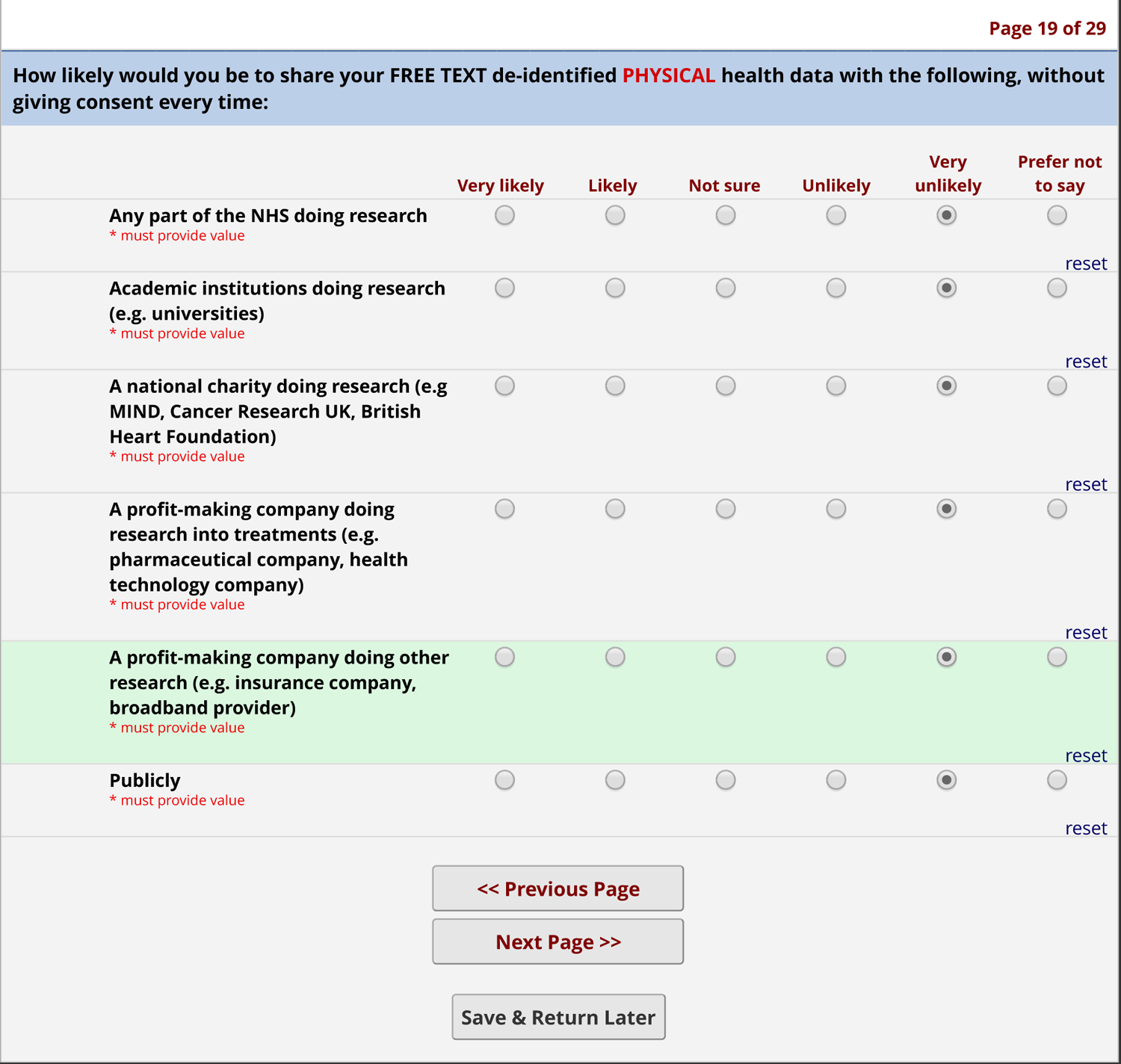
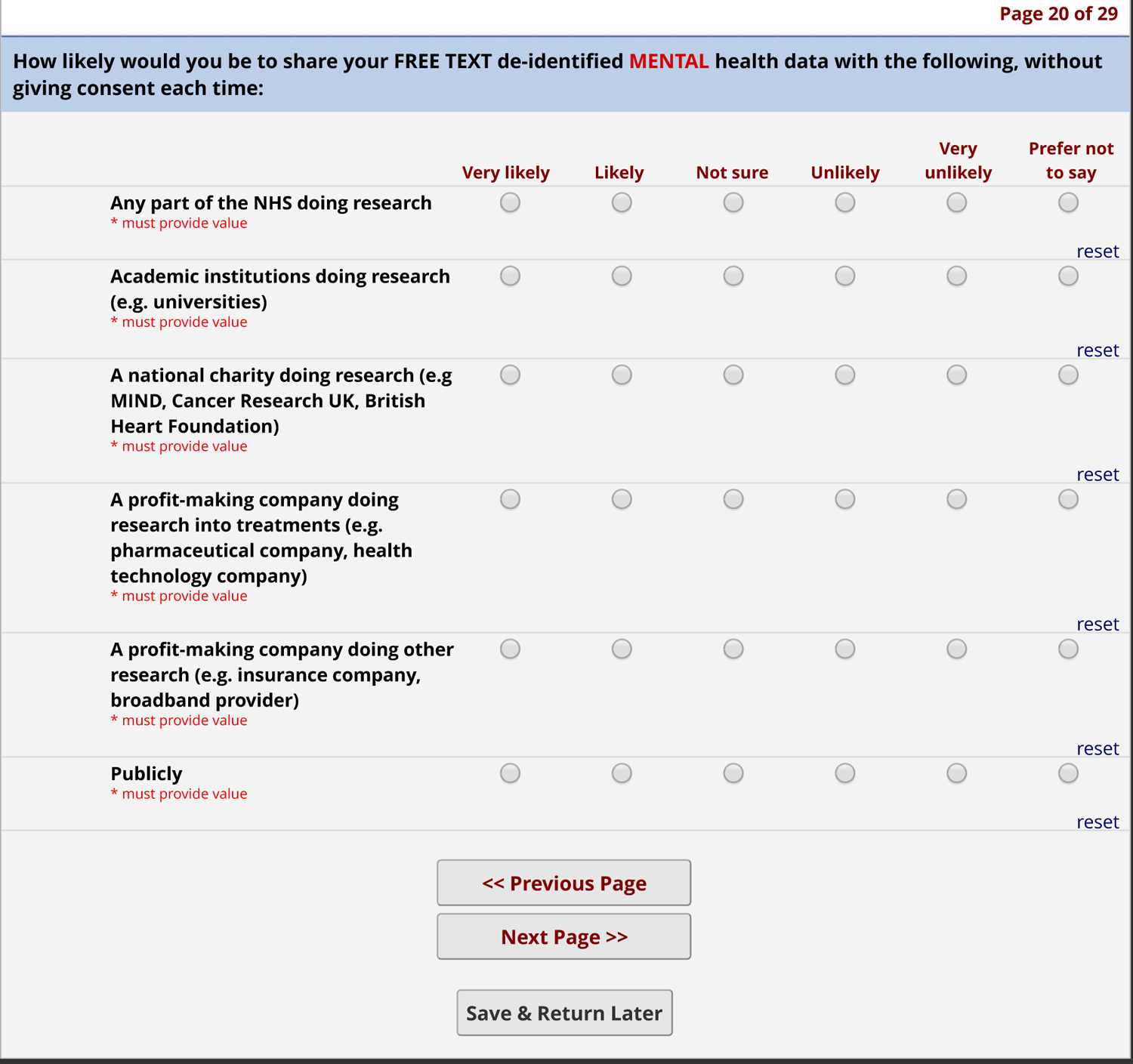
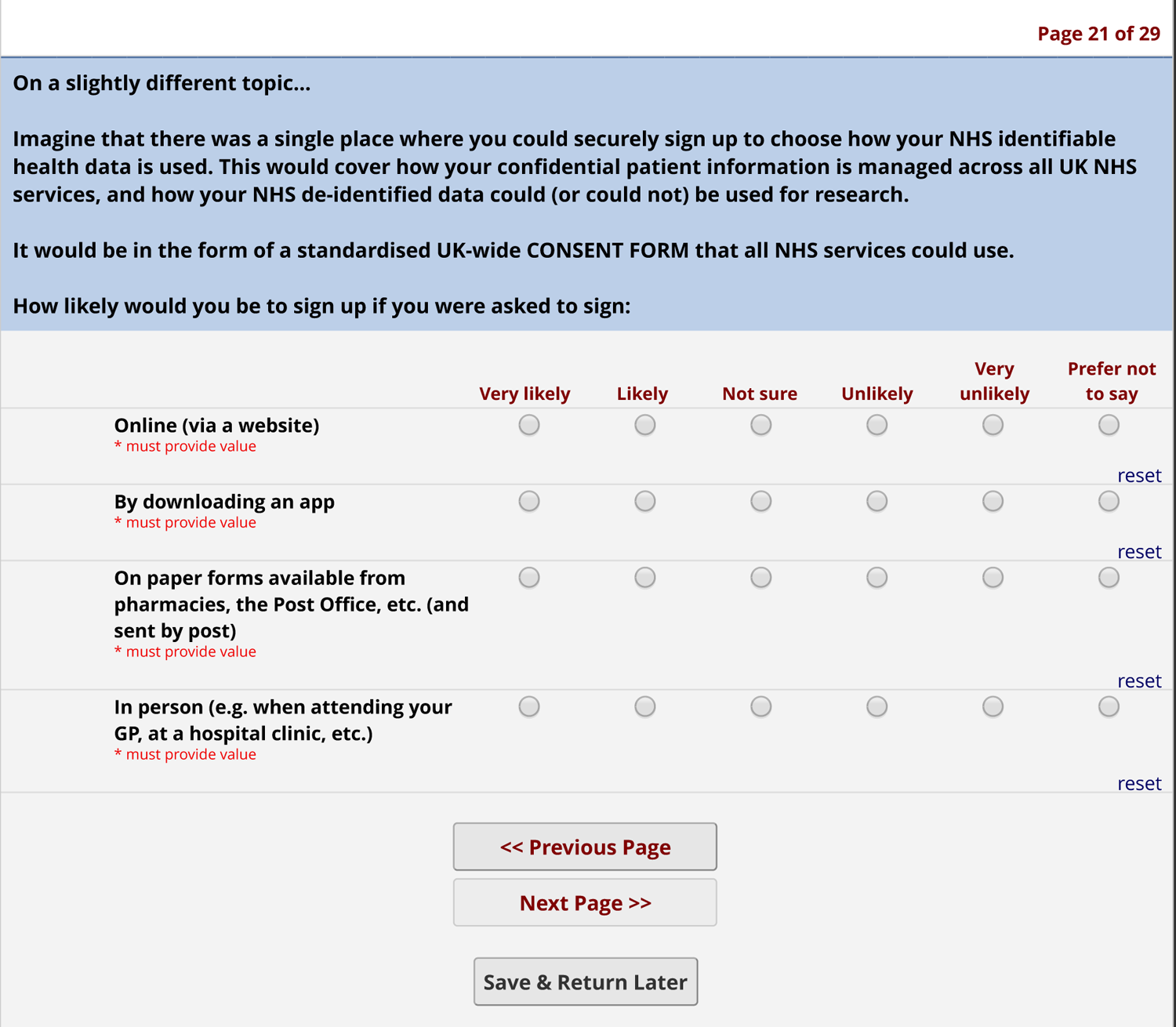
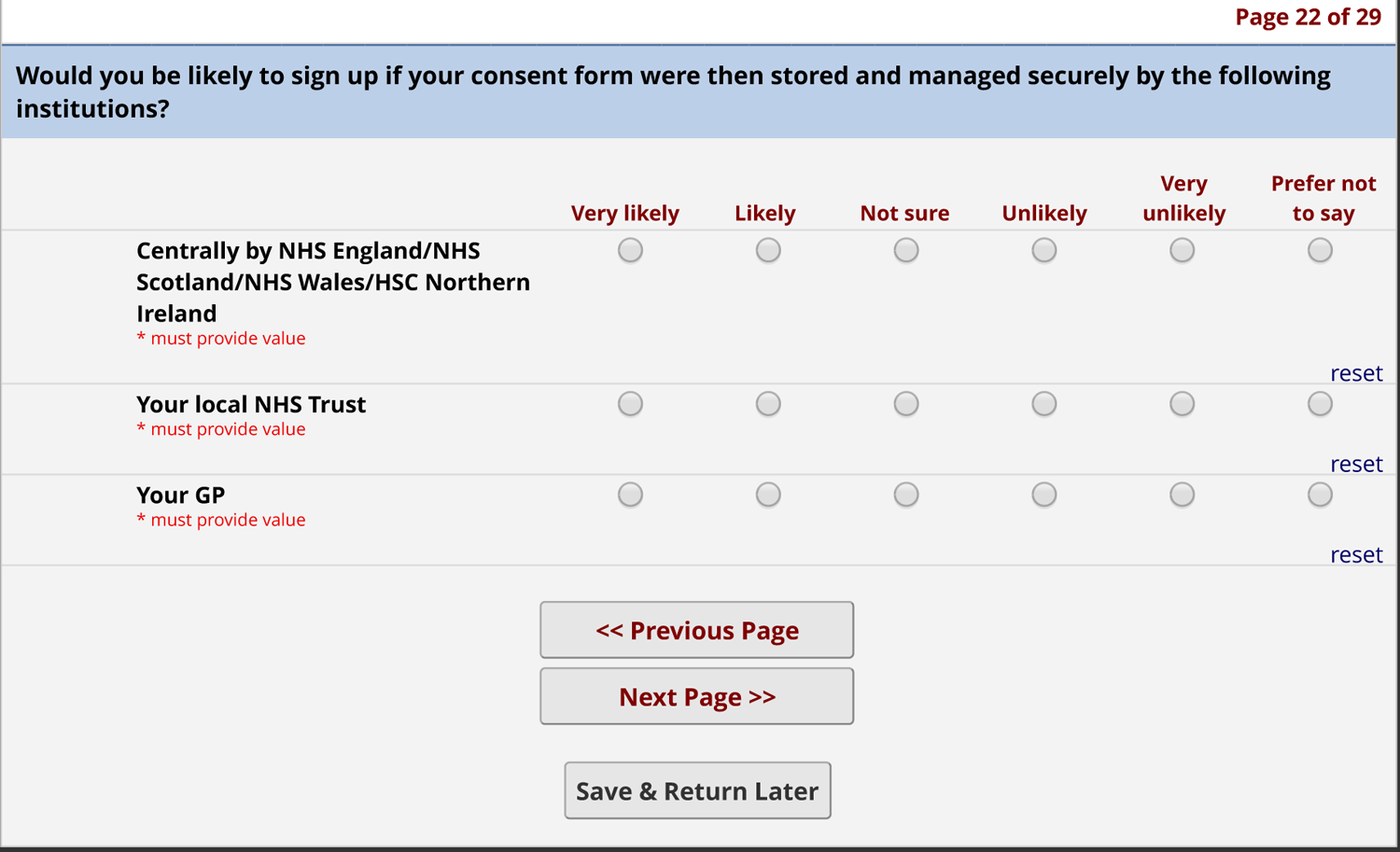
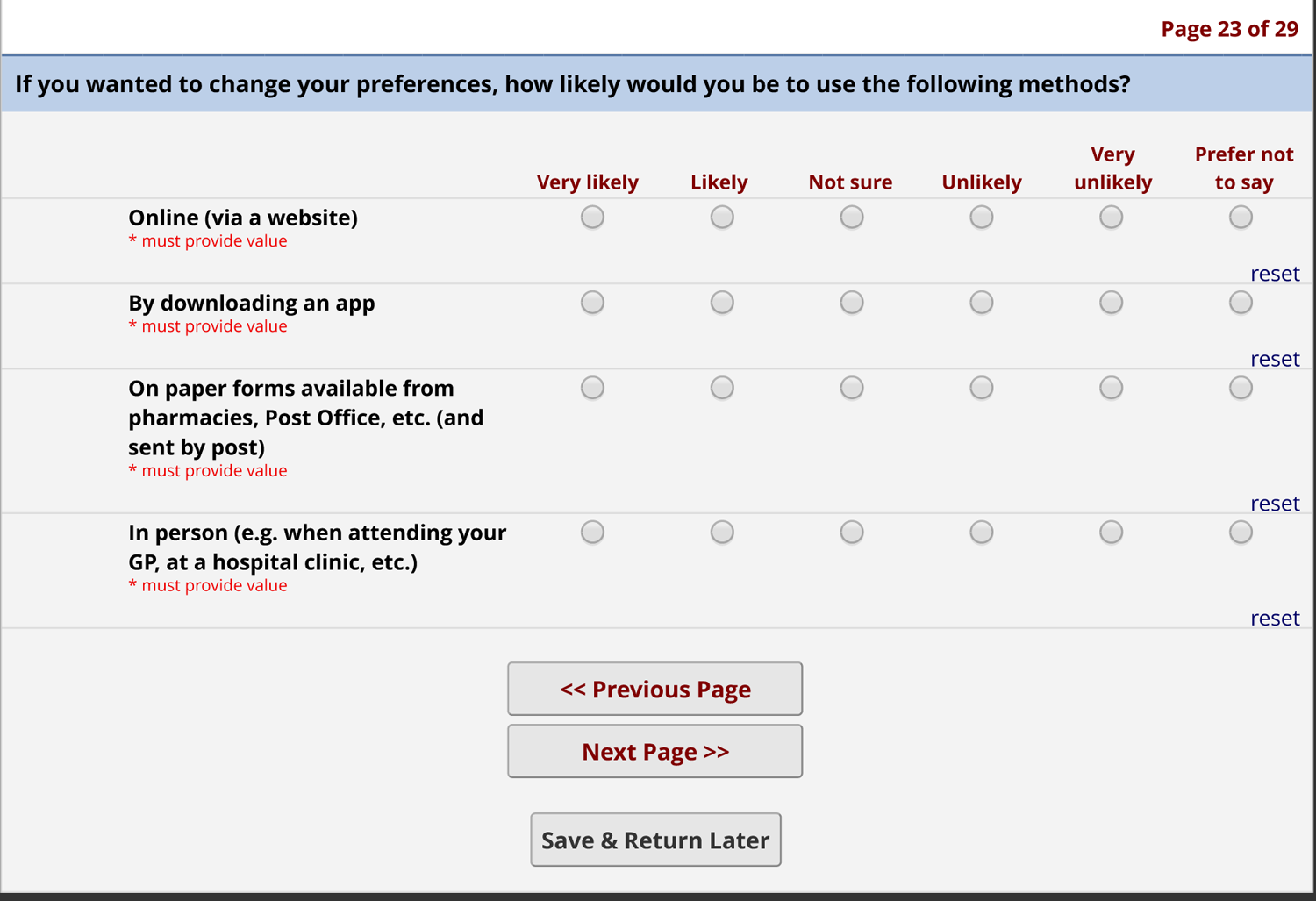
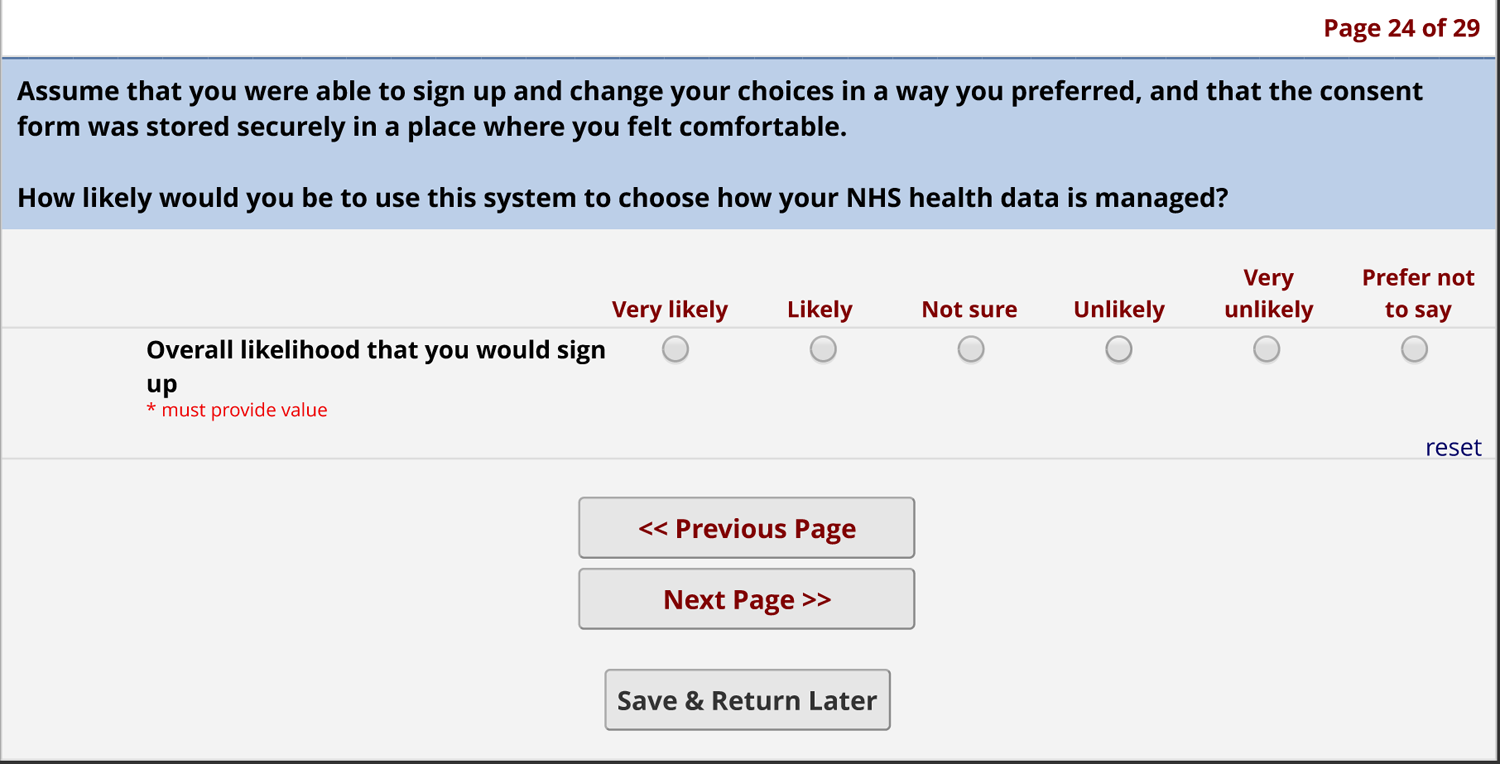
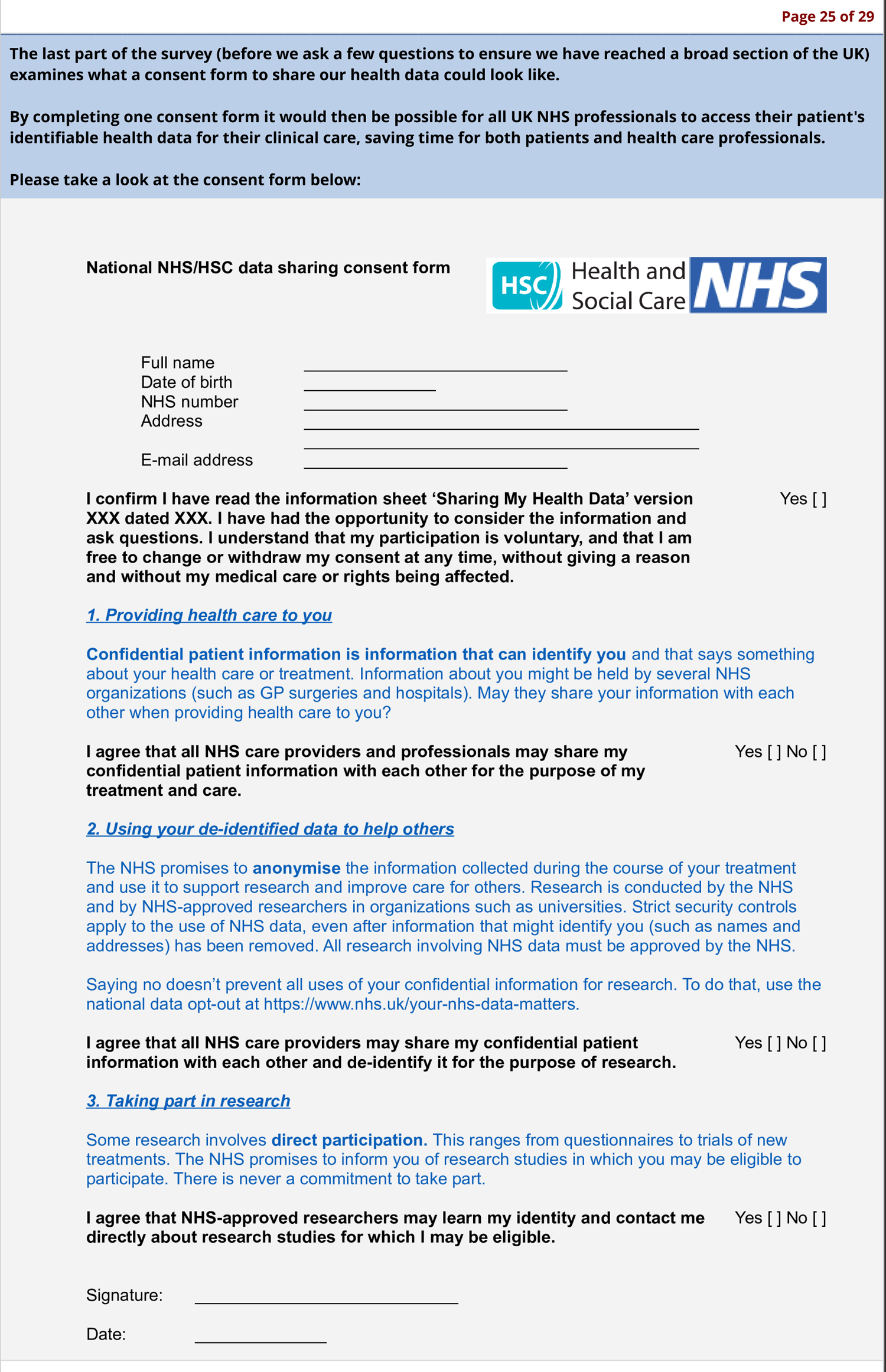
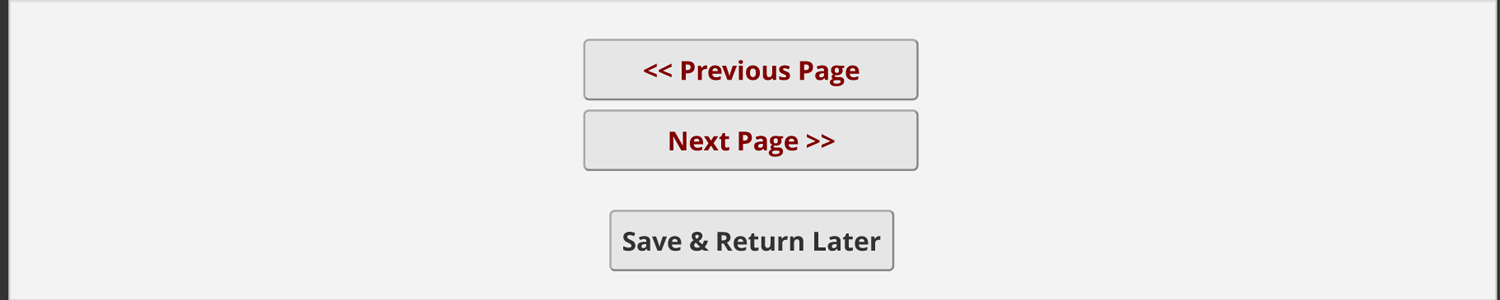
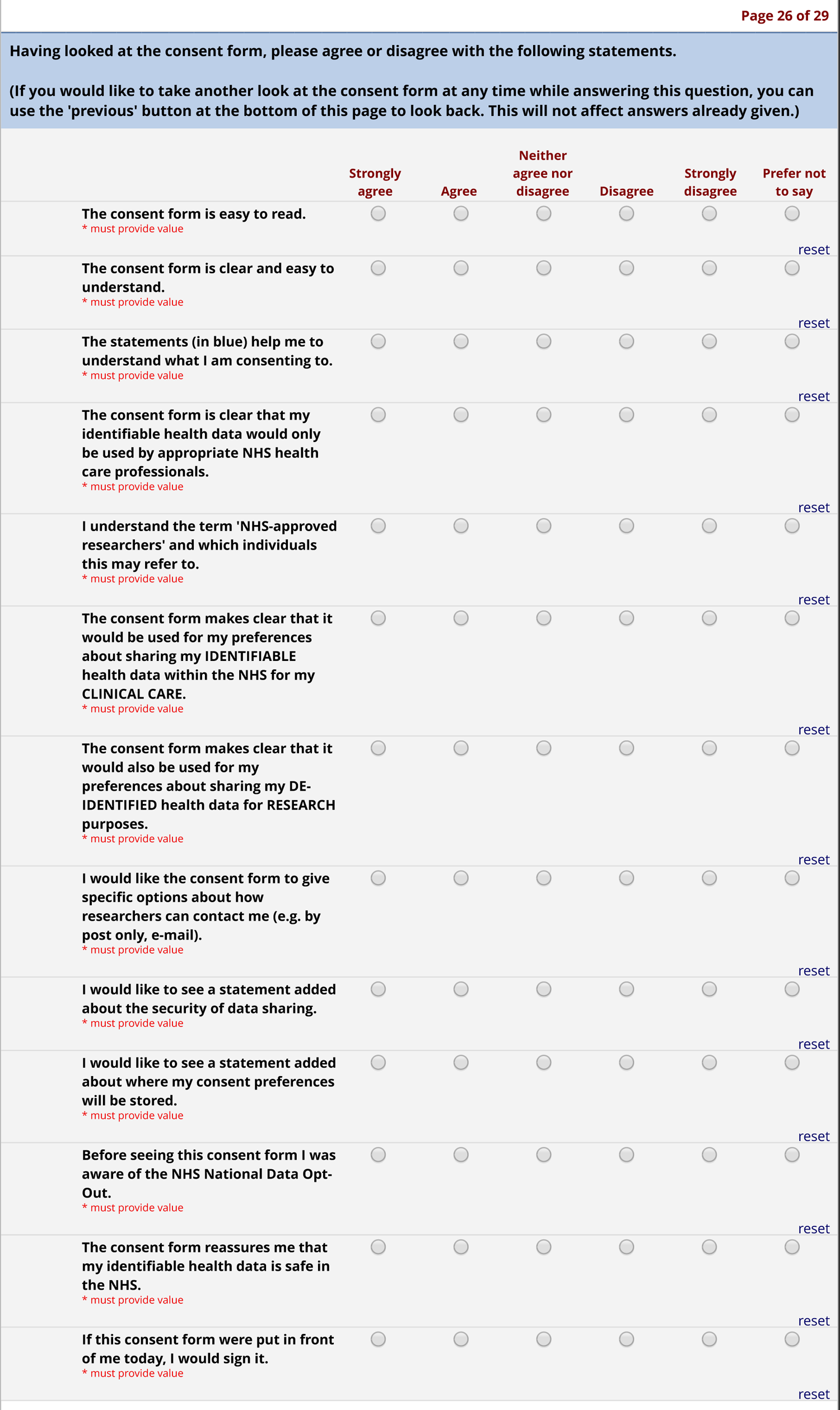
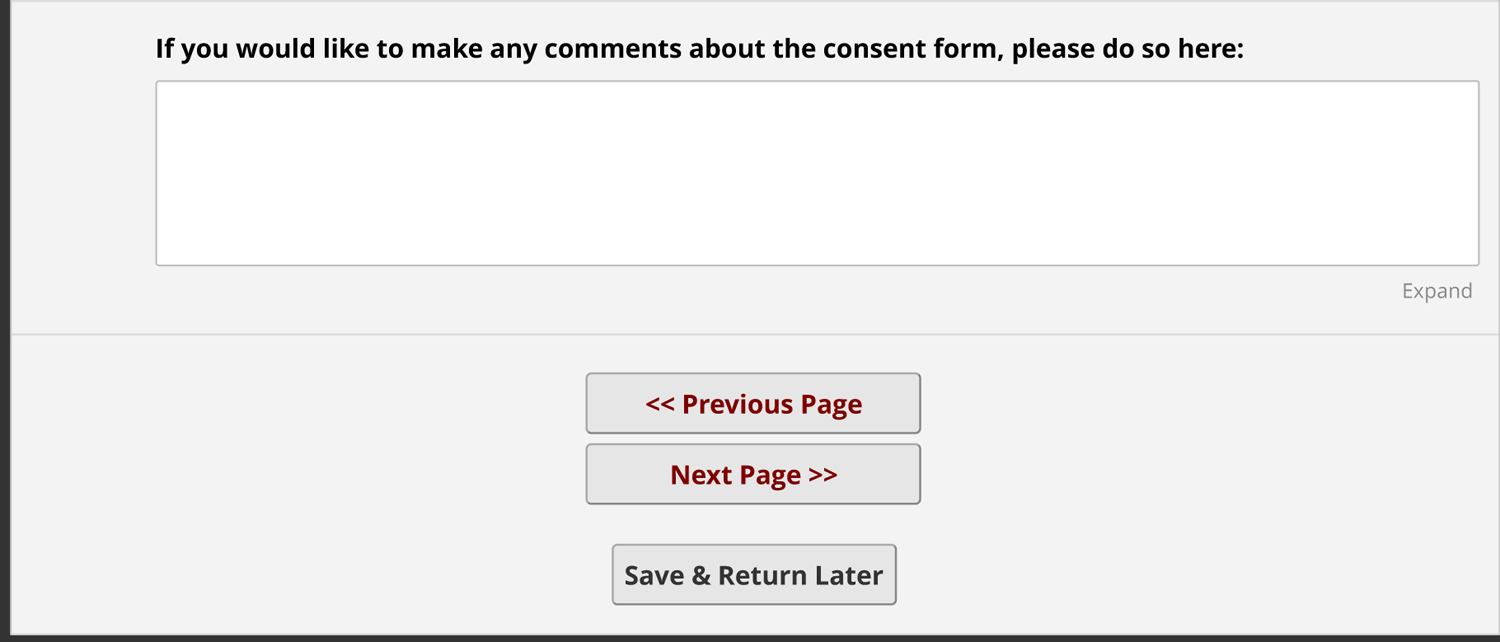
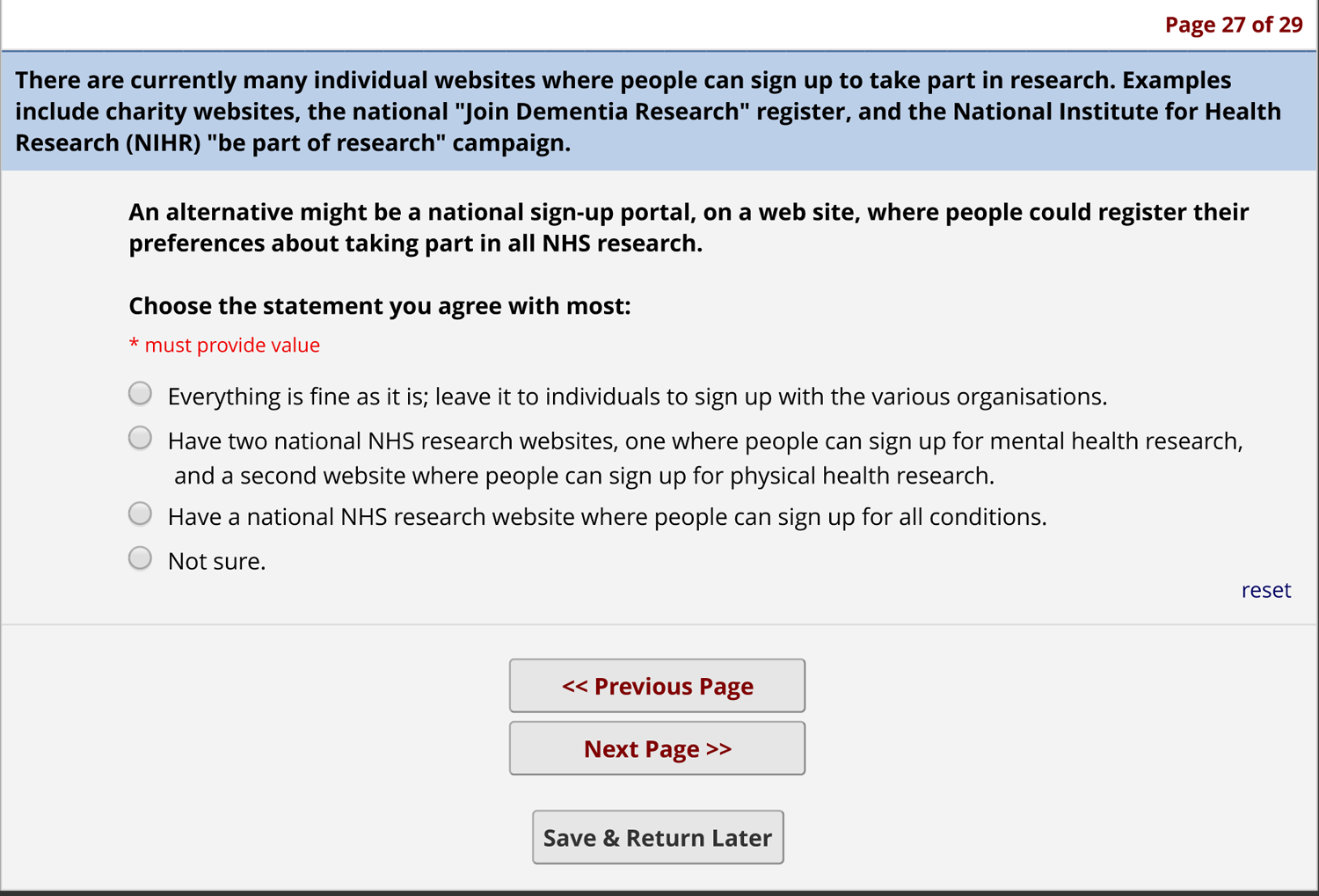
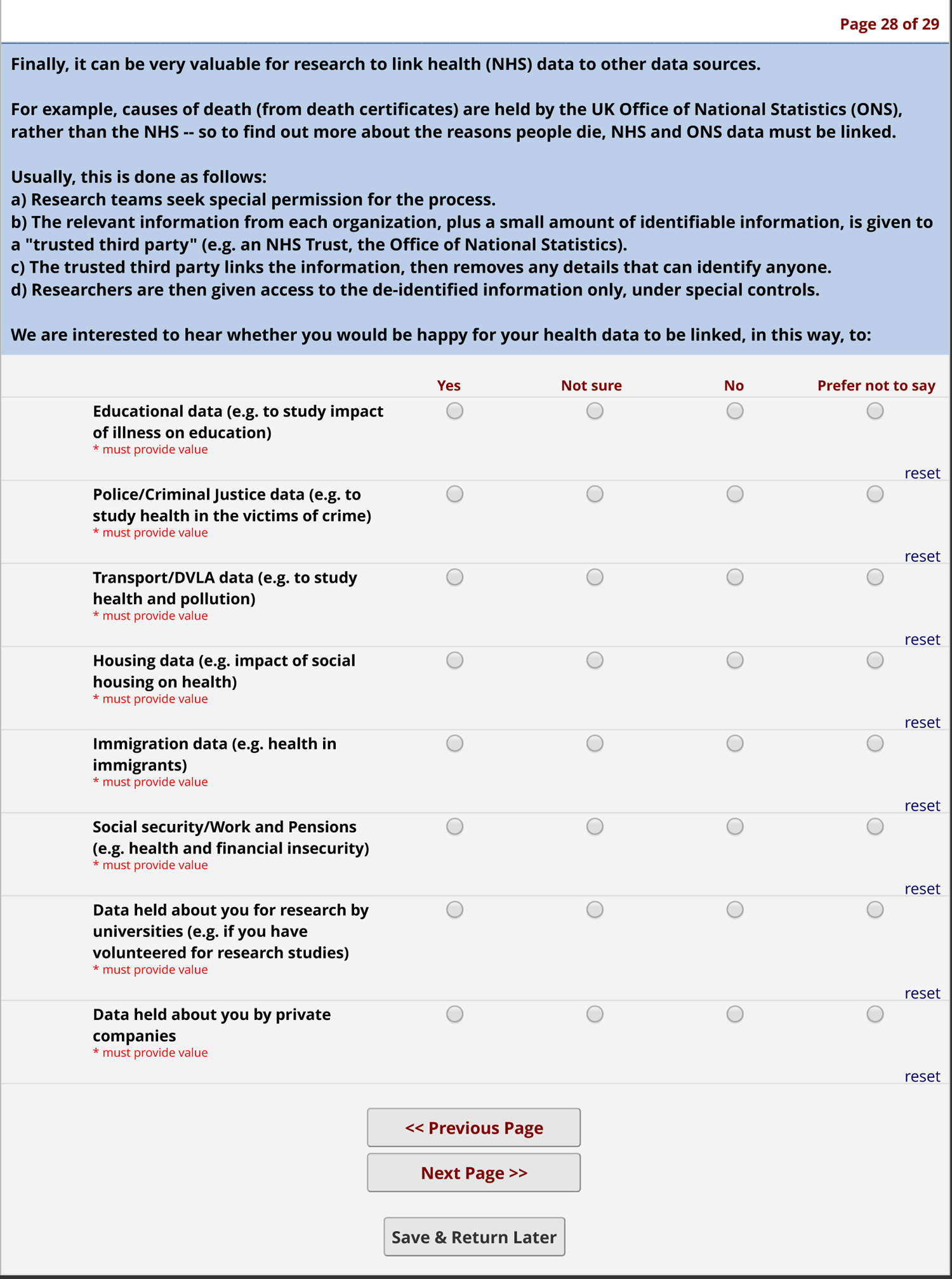
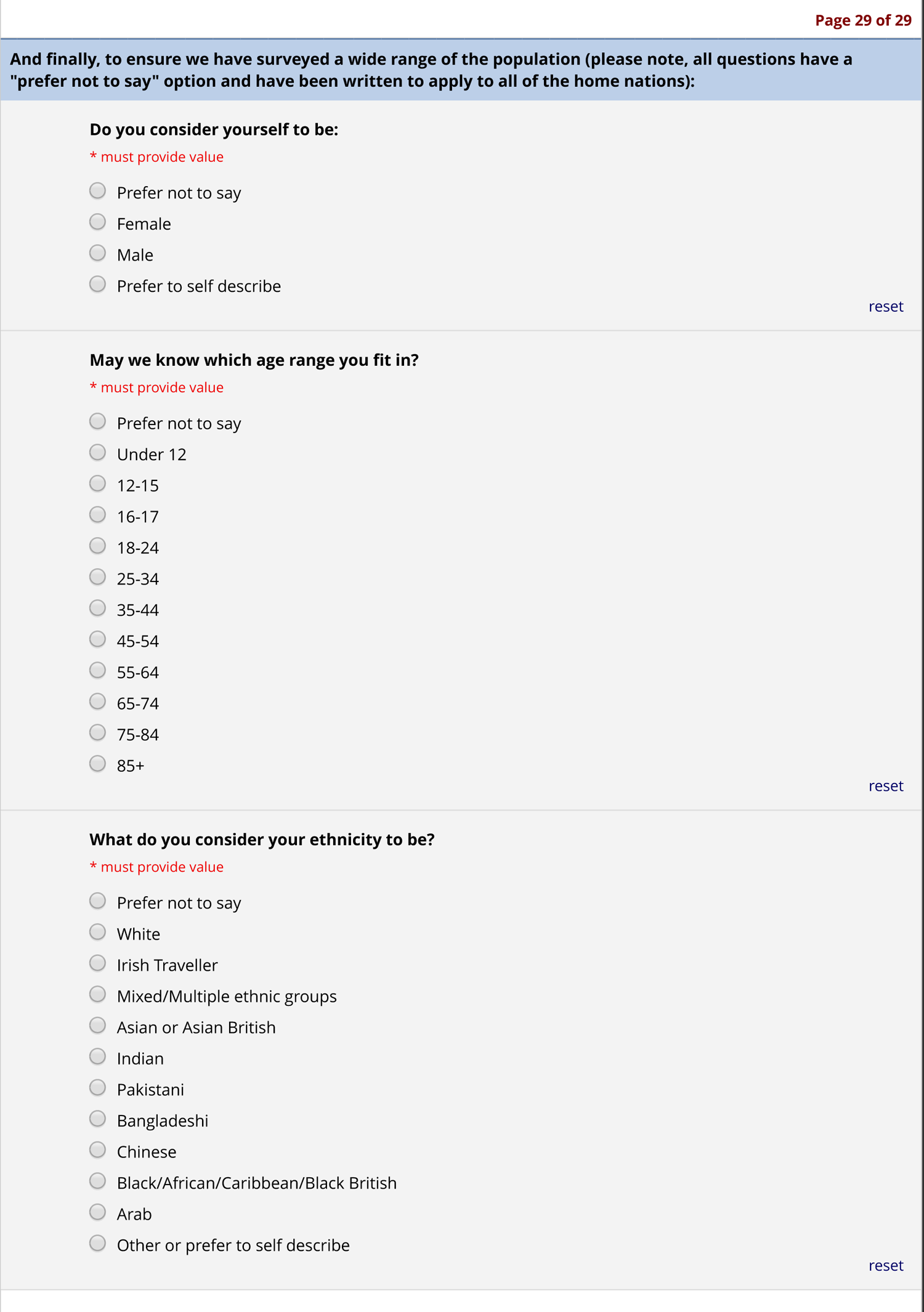
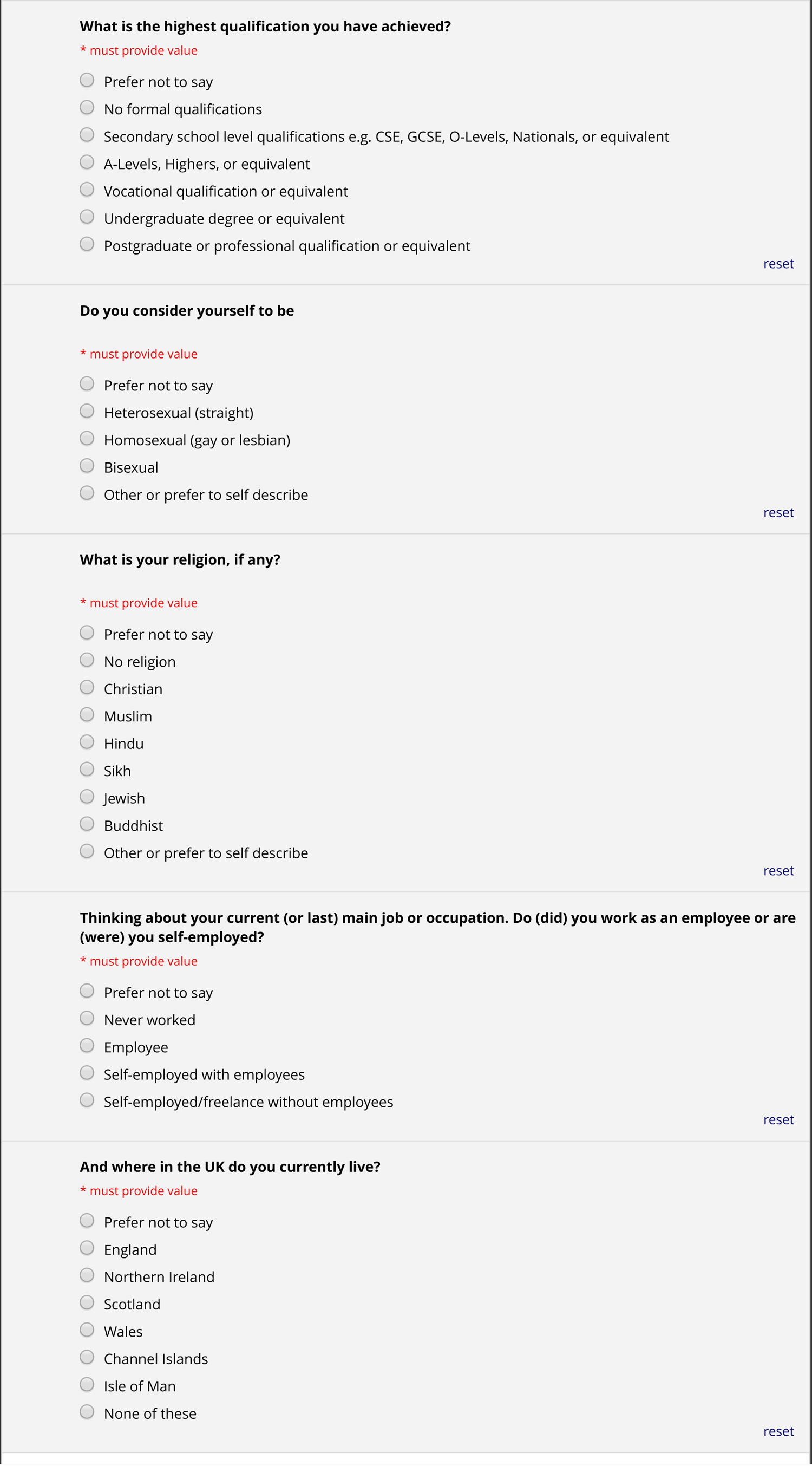
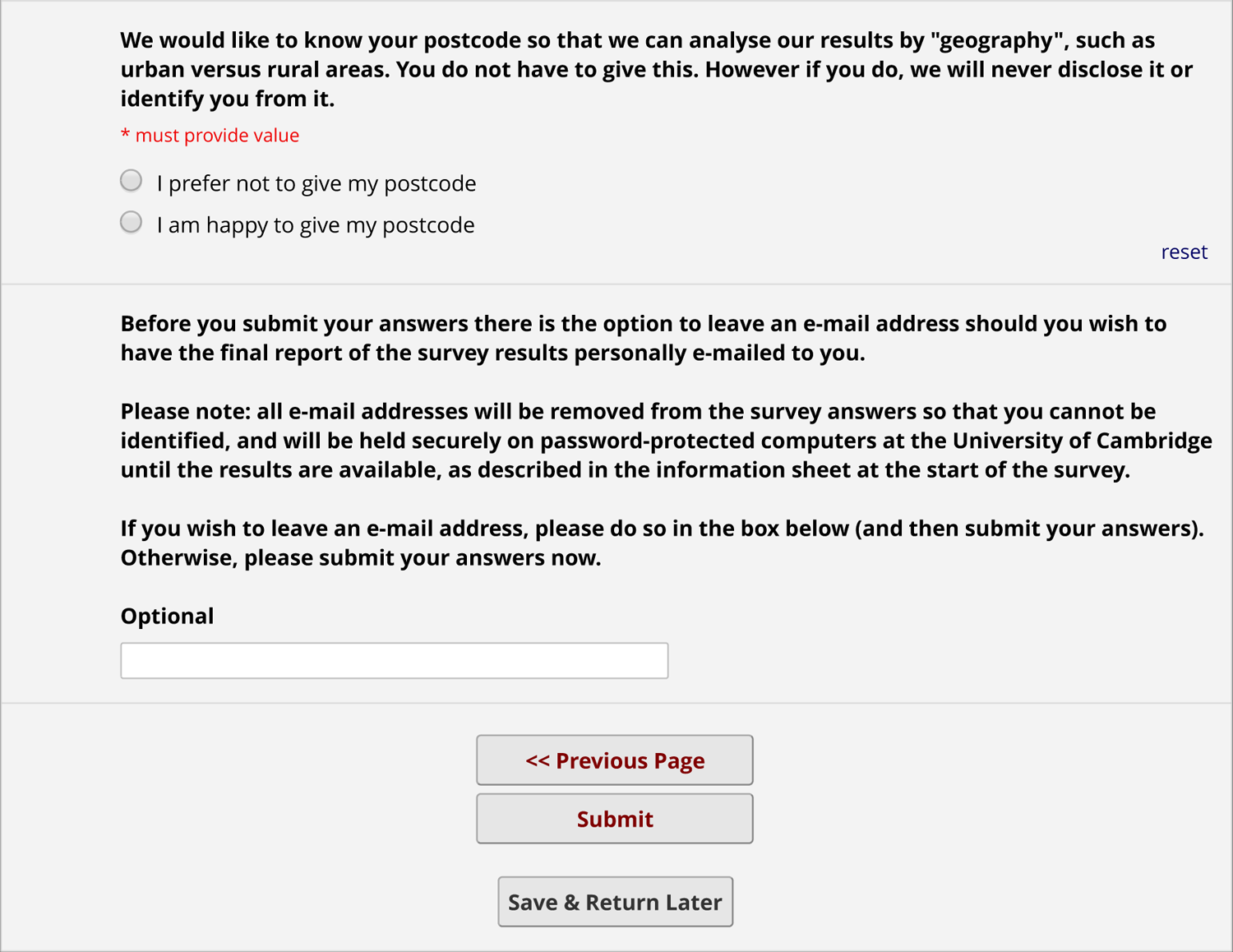
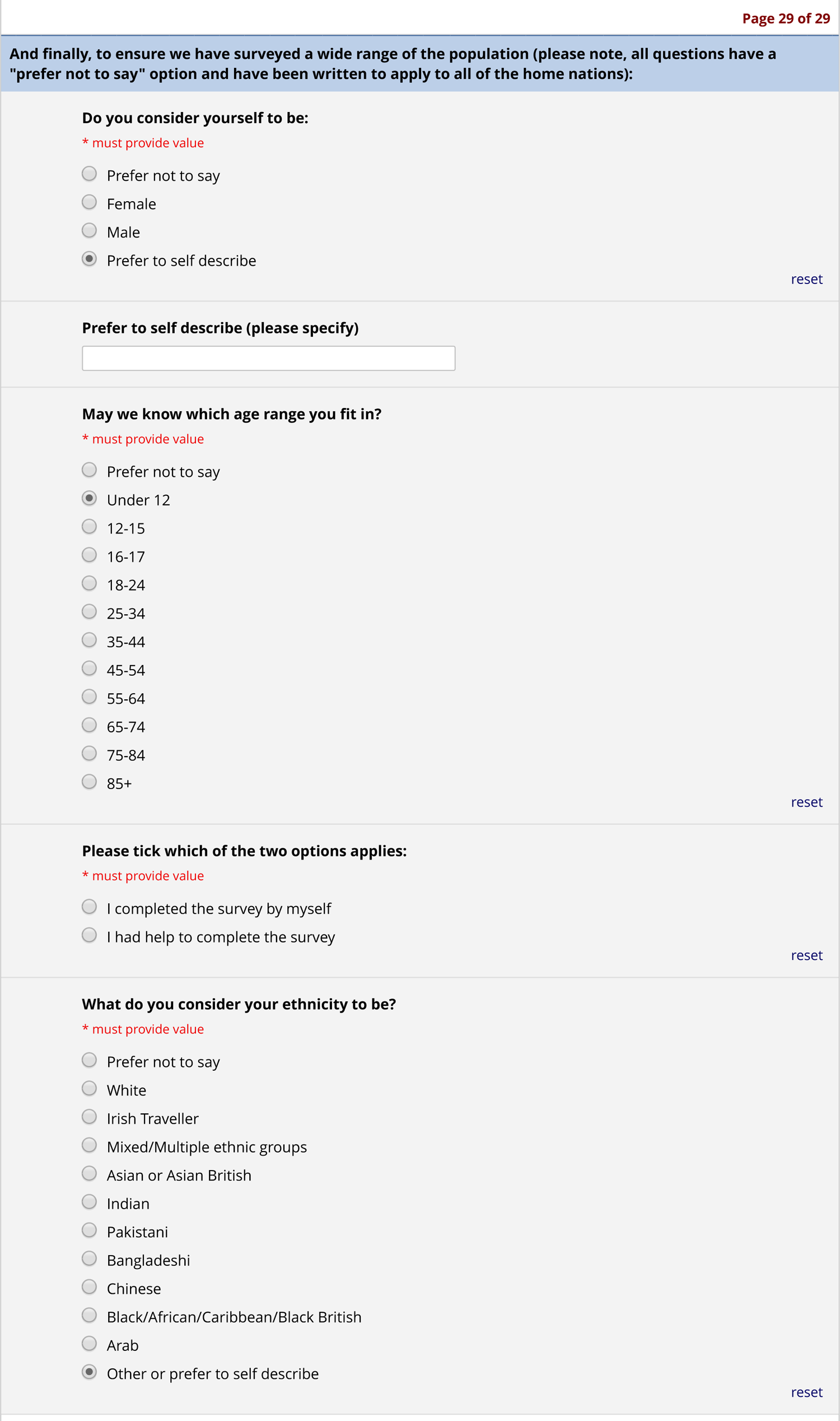
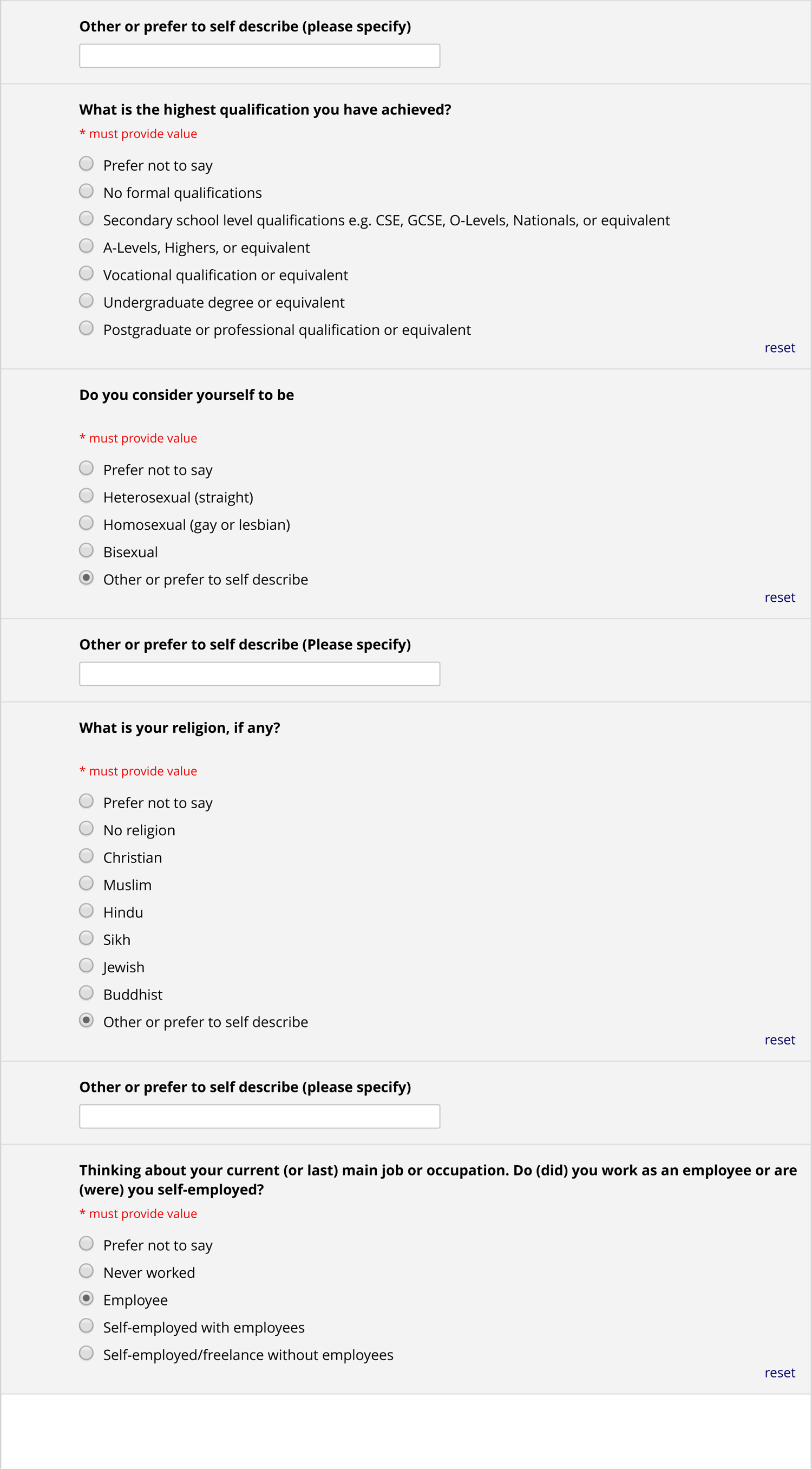
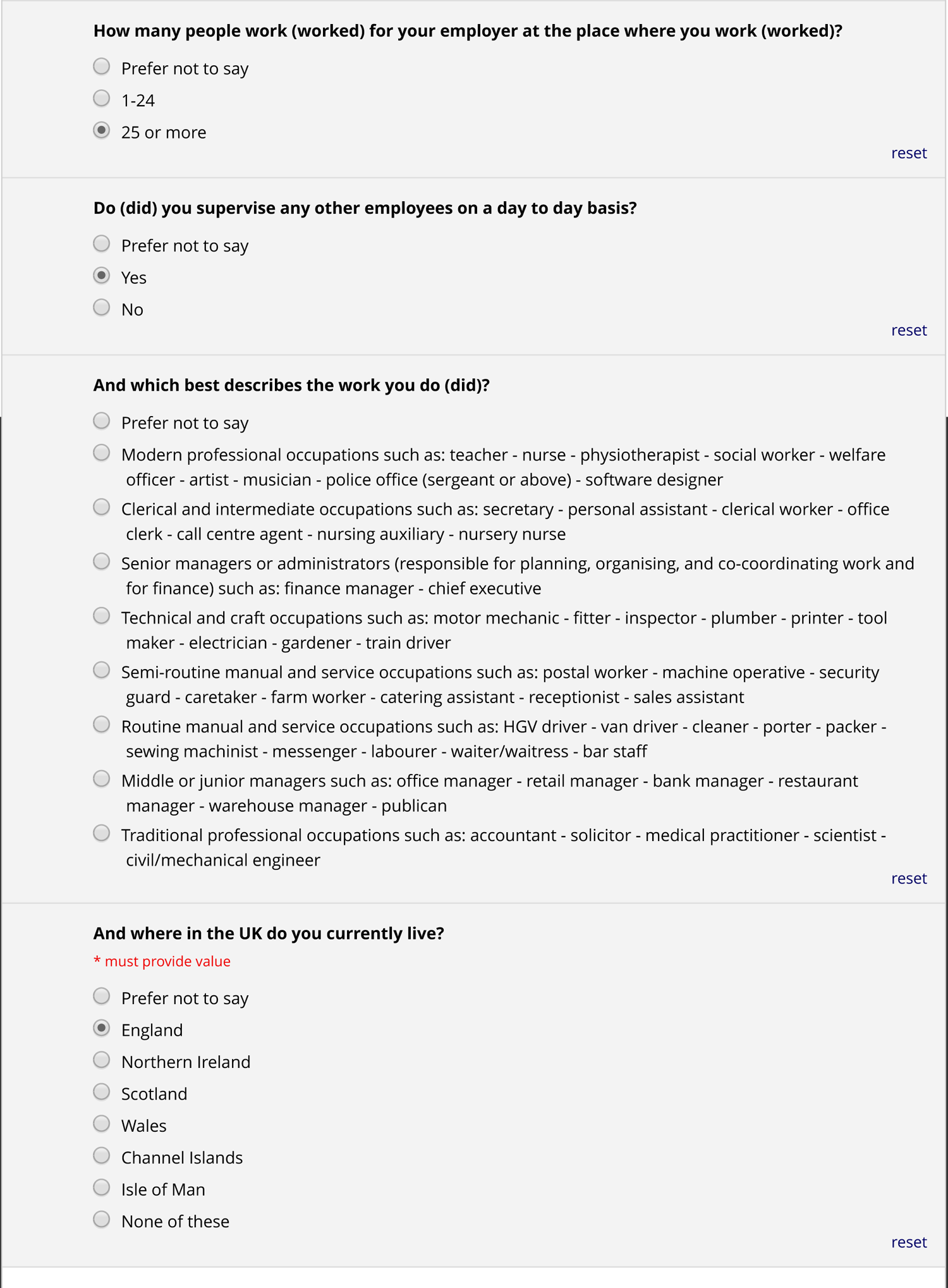
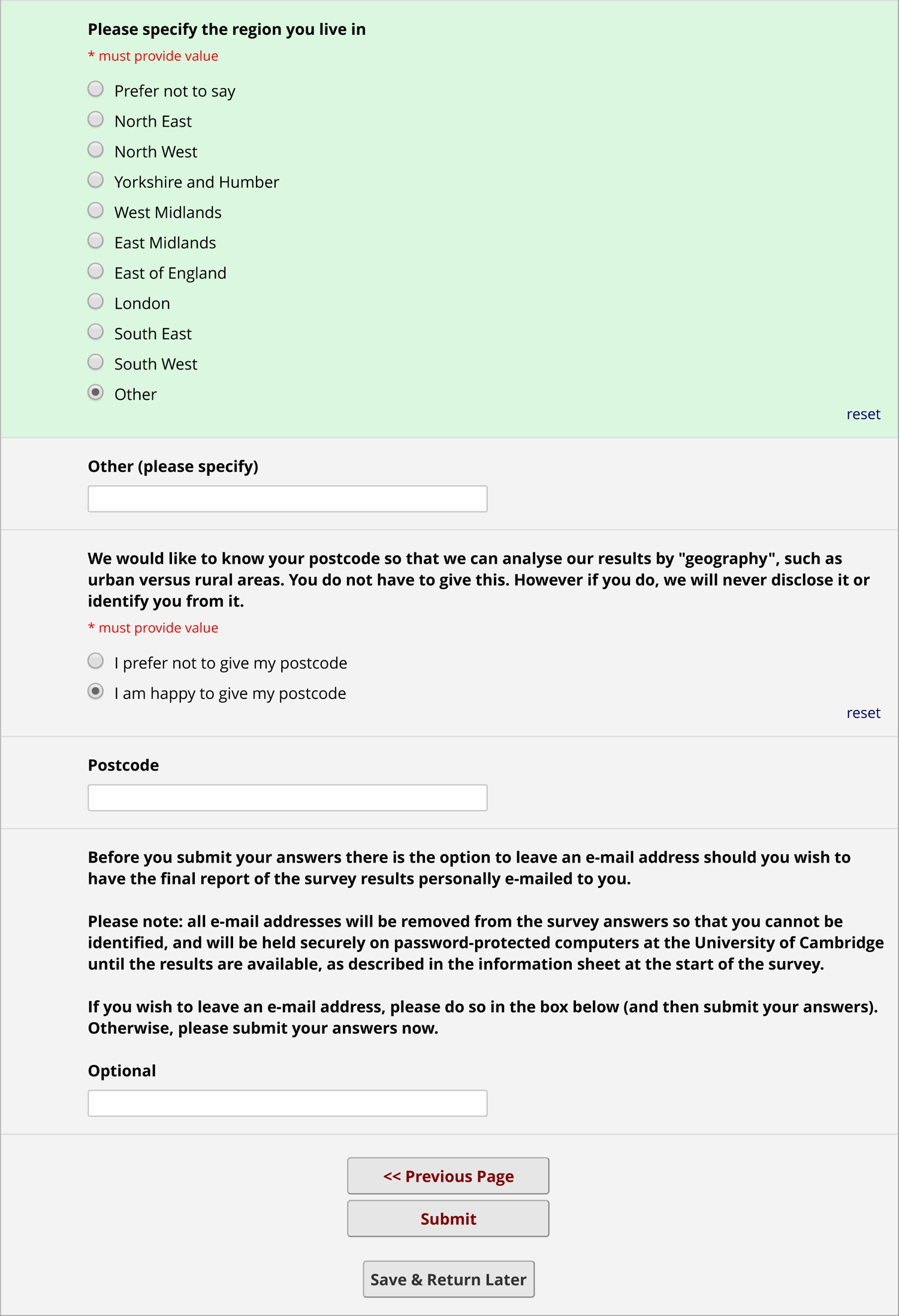
NHS Data Consent Survey

### Appendix C STROBE statement

**Appendix C** is a STROBE statement for cross-sectional studies (page numbers for composite PDF including main manuscript and supplementary materials).

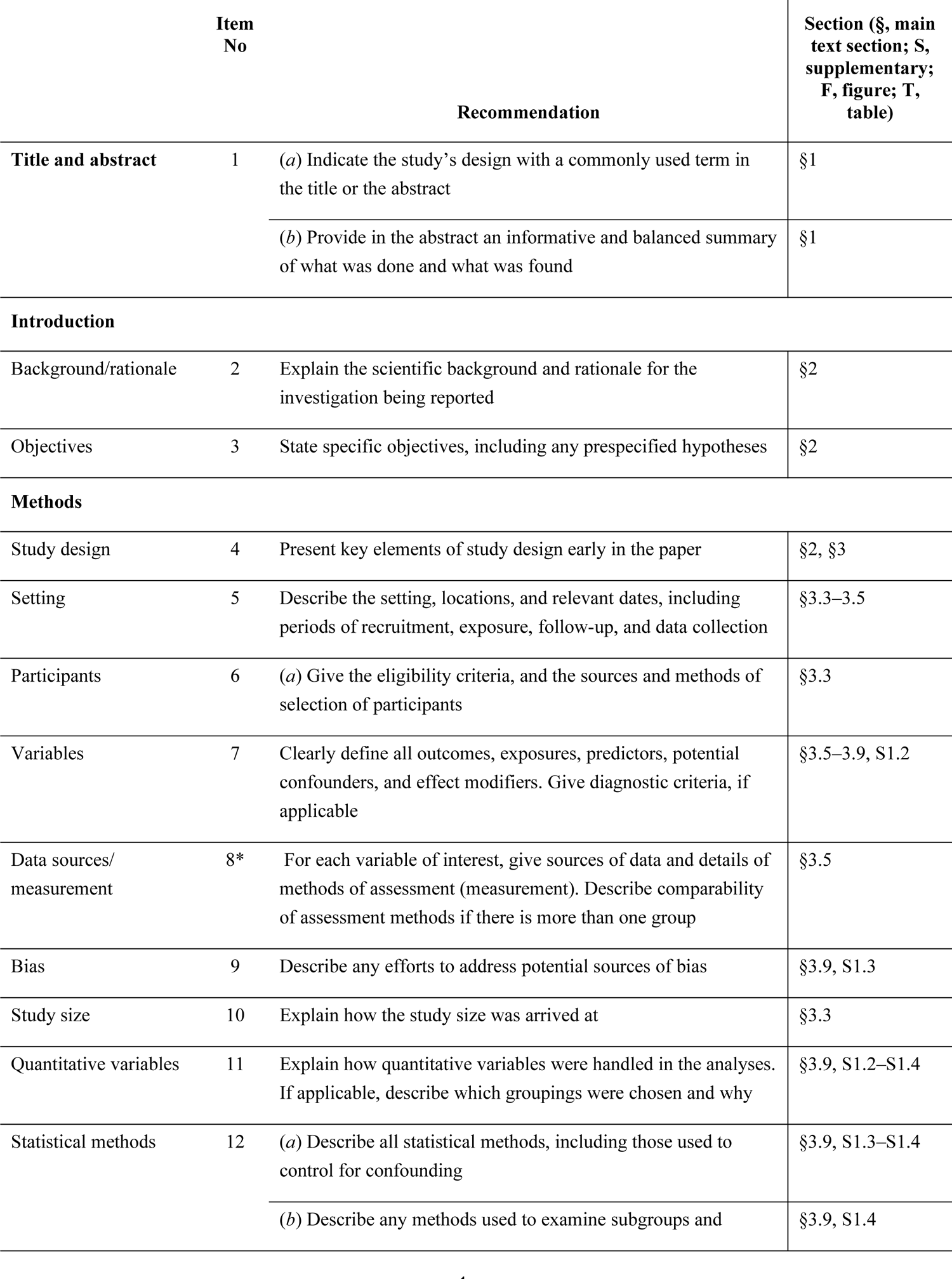
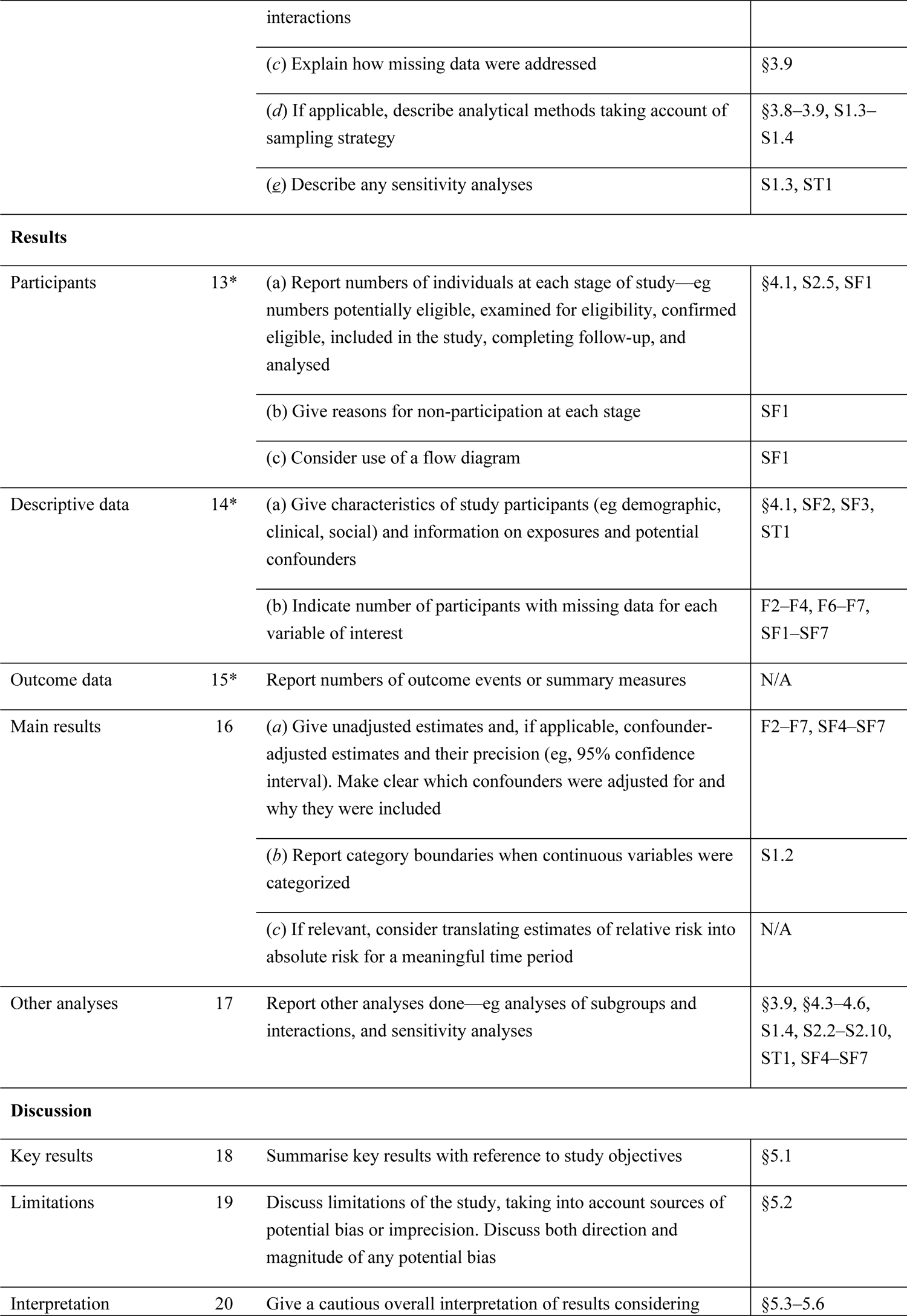
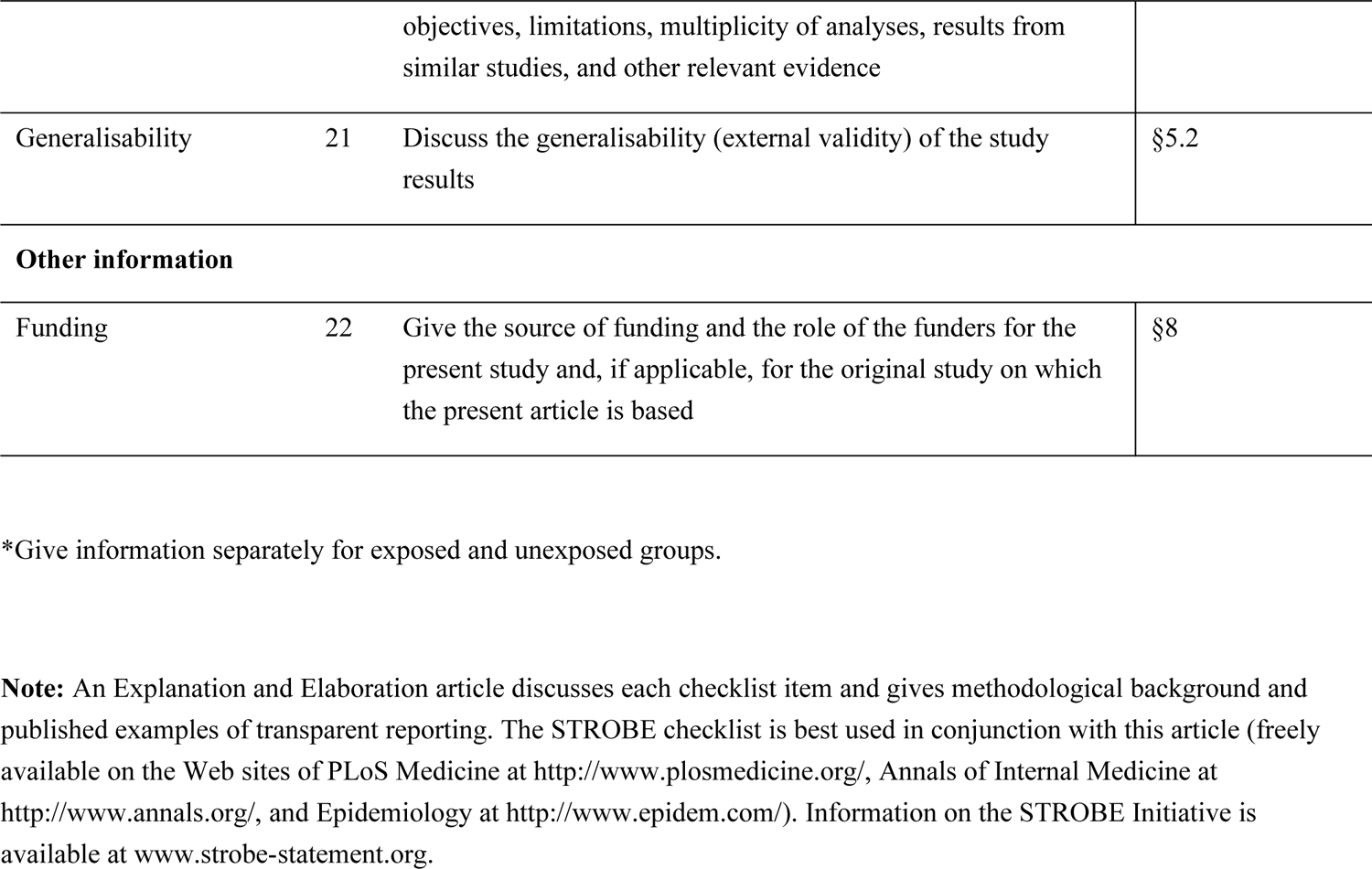

### S6. Supplementary Figure Legends

**Supplementary Figure 1.**
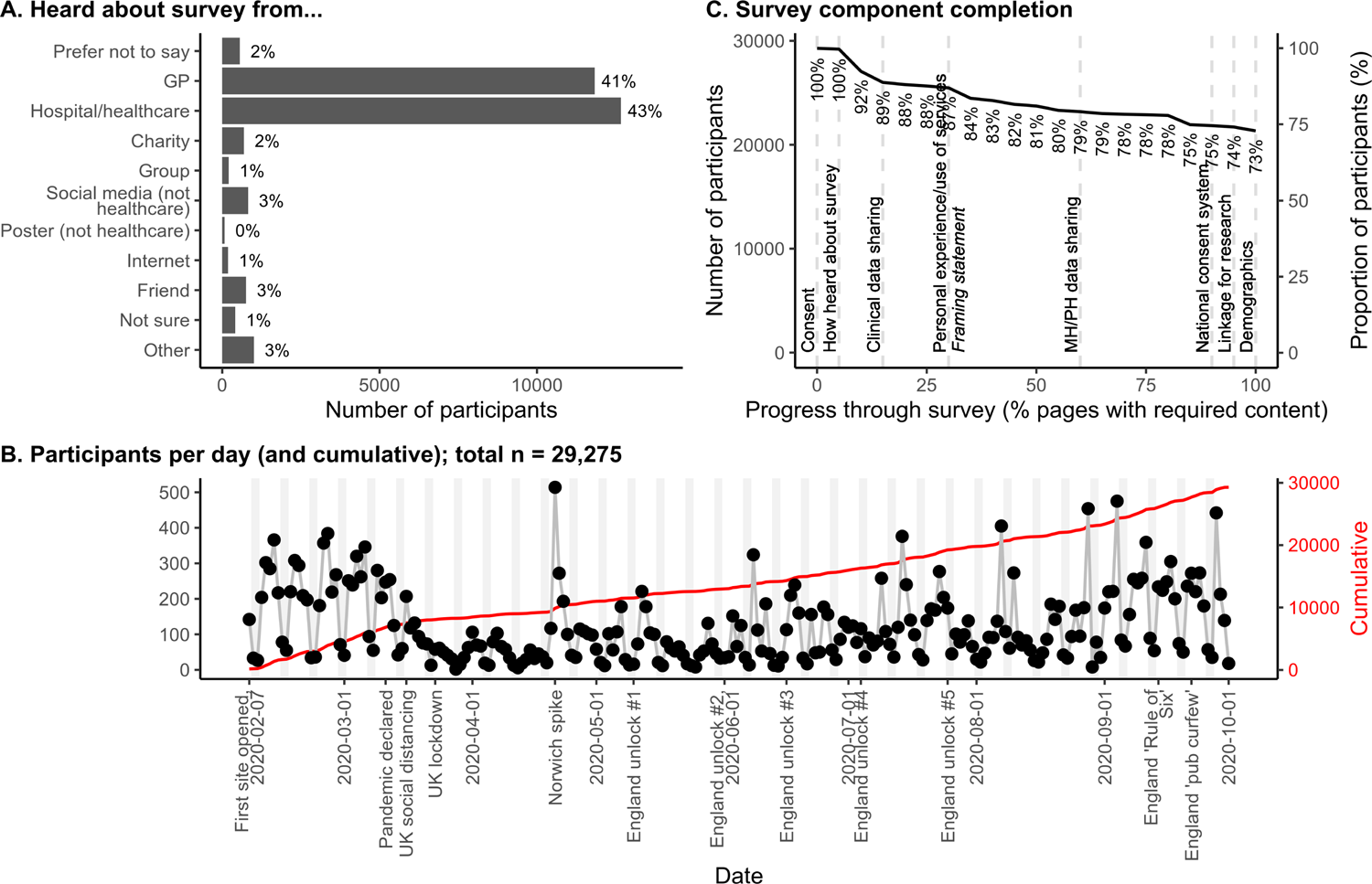
Recruitment sources, participation over time (shading indicates weekends), and survey completion. The denominator for percentages is the total number of consented participants.

**Supplementary Figure 2.**
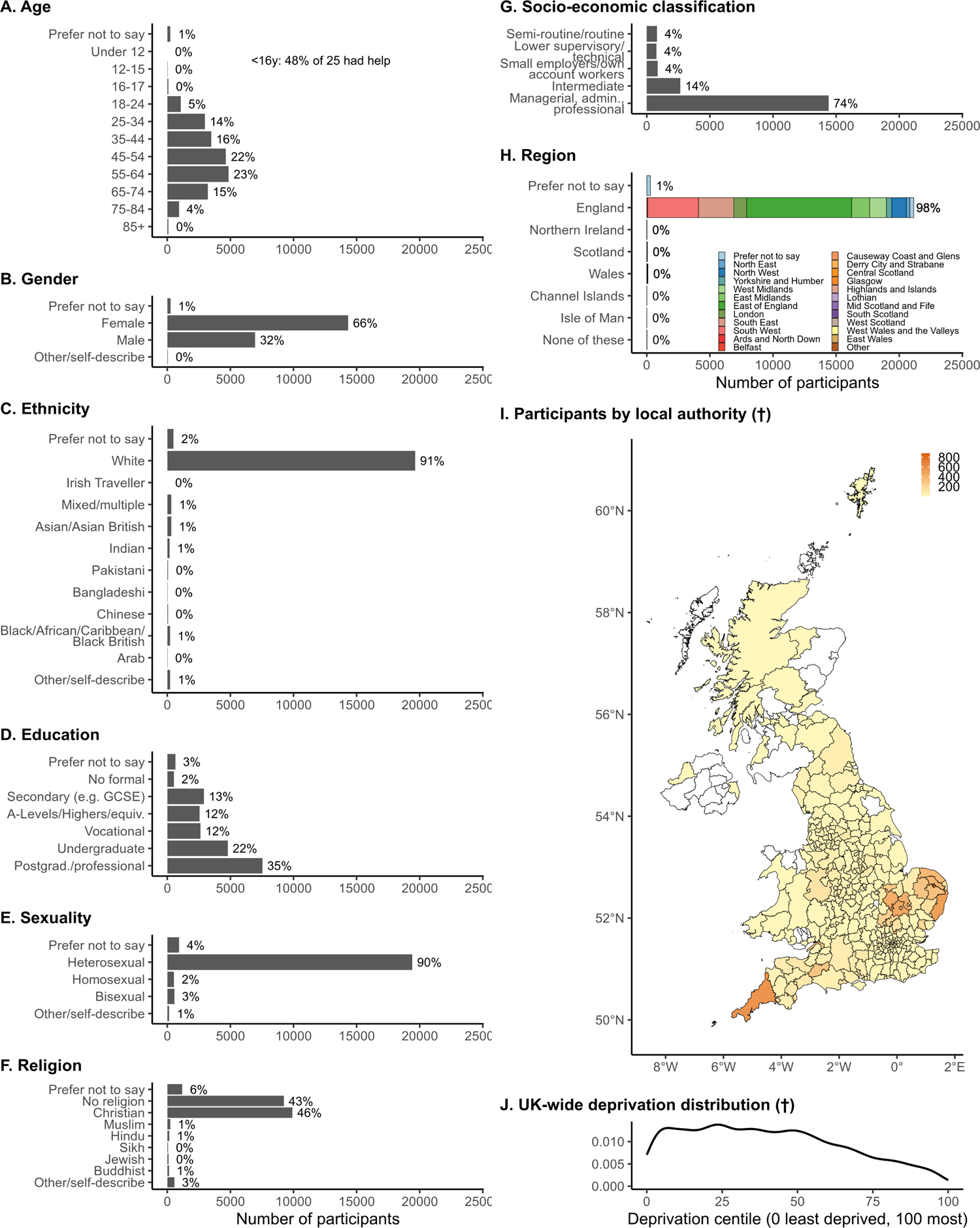
Demographics of respondents. The denominator for percentages is the number of people who answered each question. For panels marked †, only participants who provided a postcode are included. ONS local authority map and deprivation centiles exclude the Channel Islands. See **Supplementary Methods** for details of geography and deprivation calculations.

**Supplementary Figure 3.**
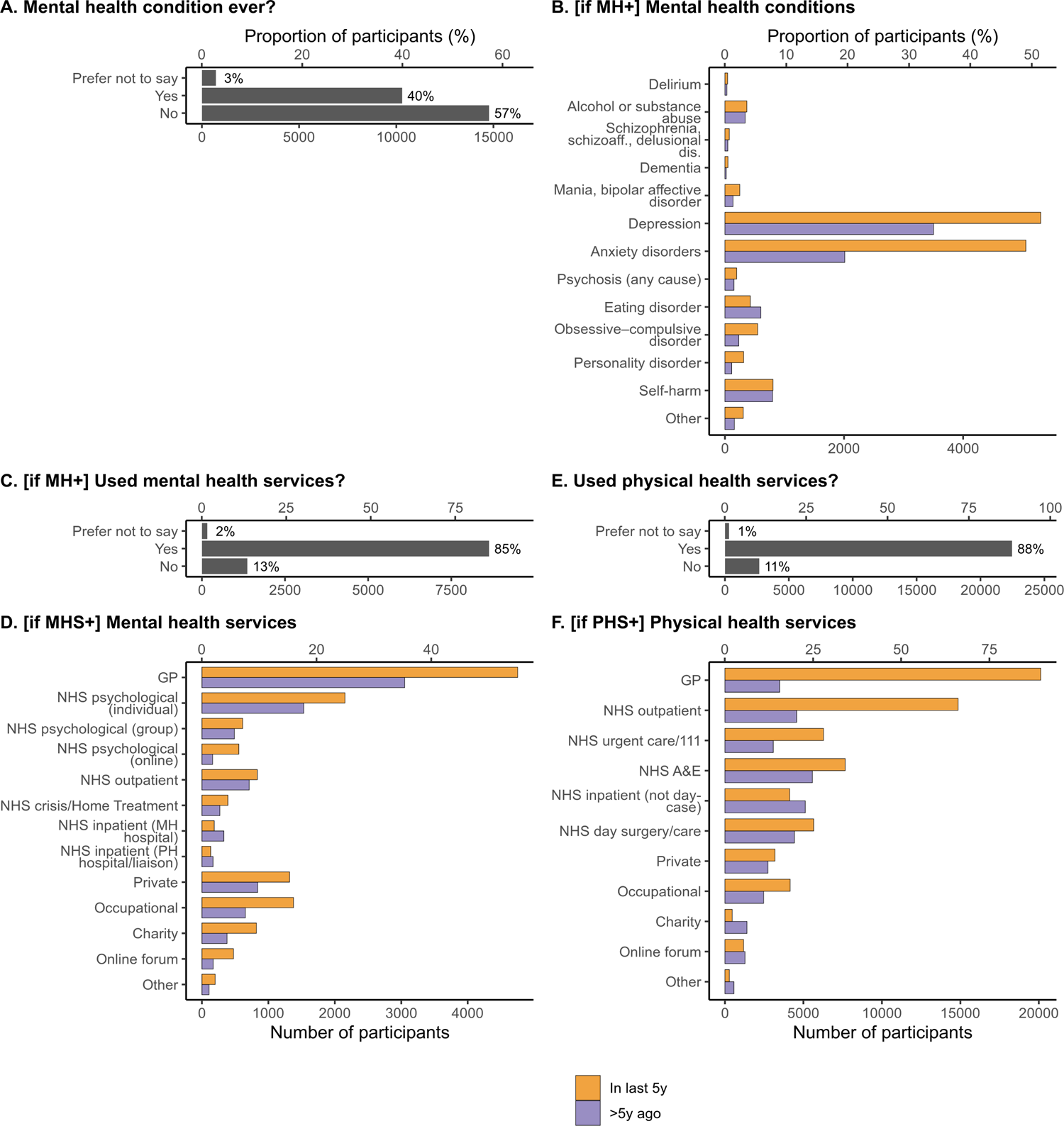
Respondents’ experience of mental health conditions/services and physical health services. The denominator for percentages is the number of people who answered each question.

**Supplementary Figure 4.**
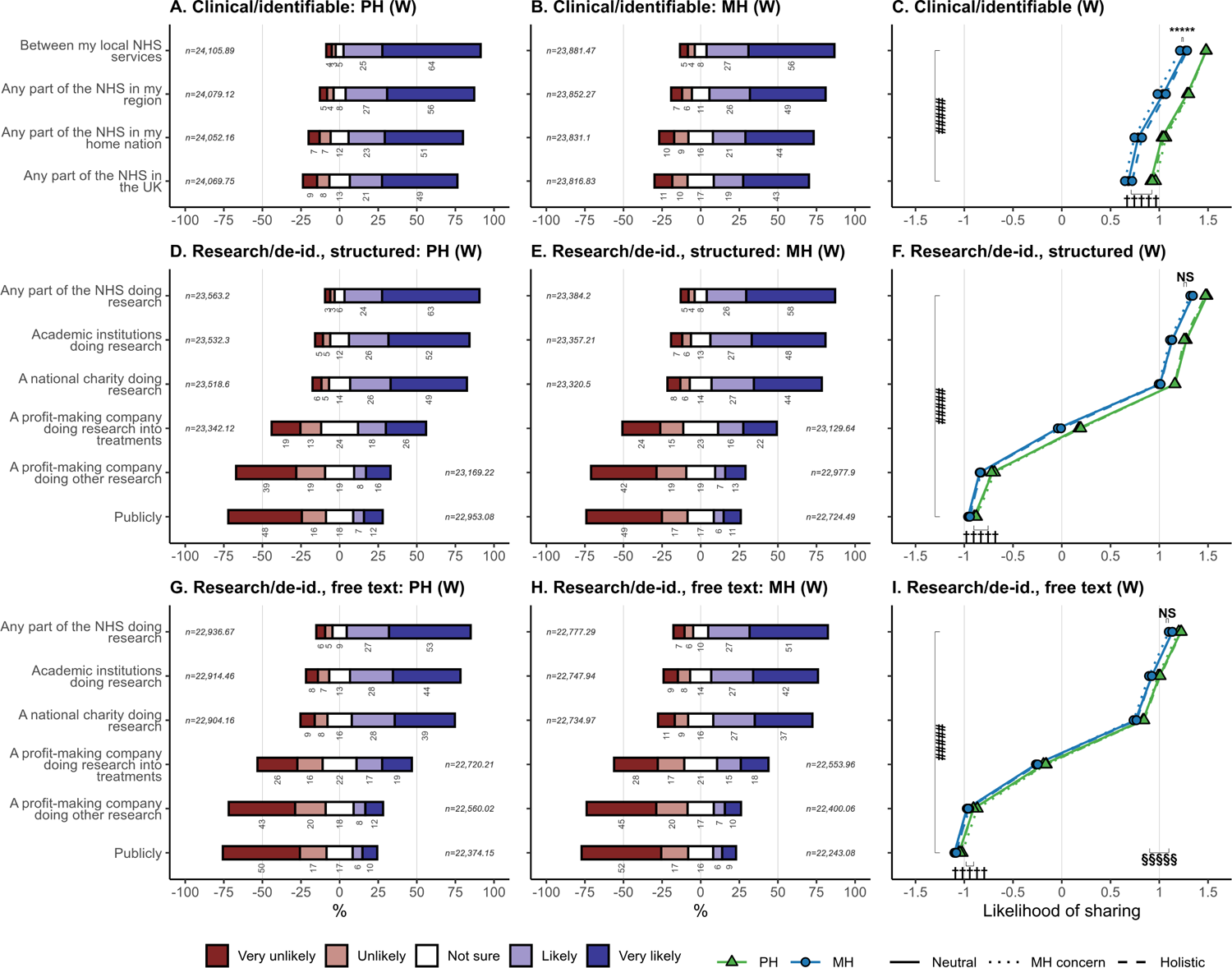
Opinions about sharing mental and physical health data, weighted according to demographic variables. This is equivalent to **Figure 3** apart from the weighting (see **Supplementary Methods**), with identical conventions. “(W)”: weighted.

**Supplementary Figure 5.**
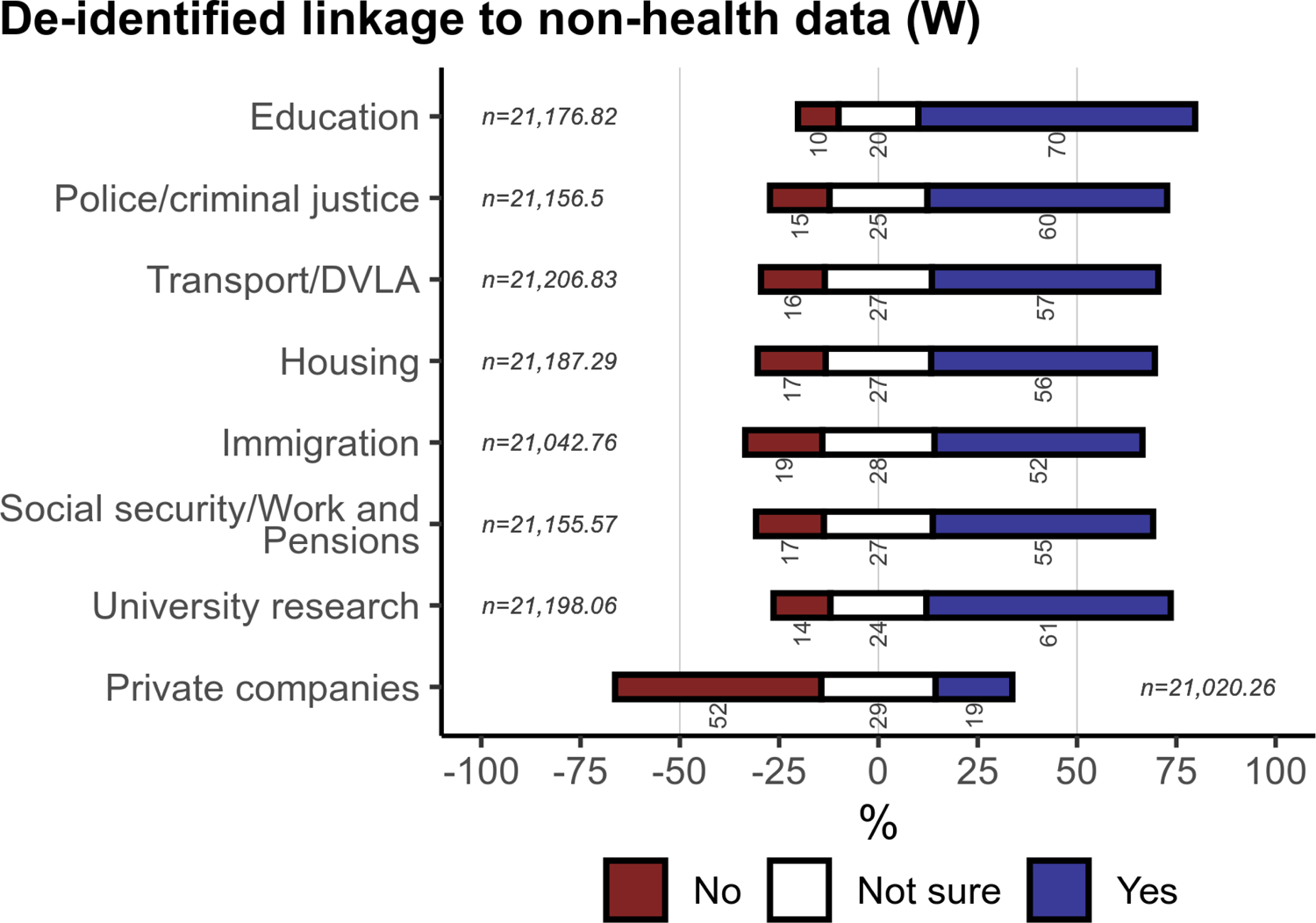
Views on linkage to non-health data for research, weighted according to demographic variables. This is equivalent to **Figure 4** apart from the weighting (see **Supplementary Methods**). “(W)”: weighted.

**Supplementary Figure 6.**
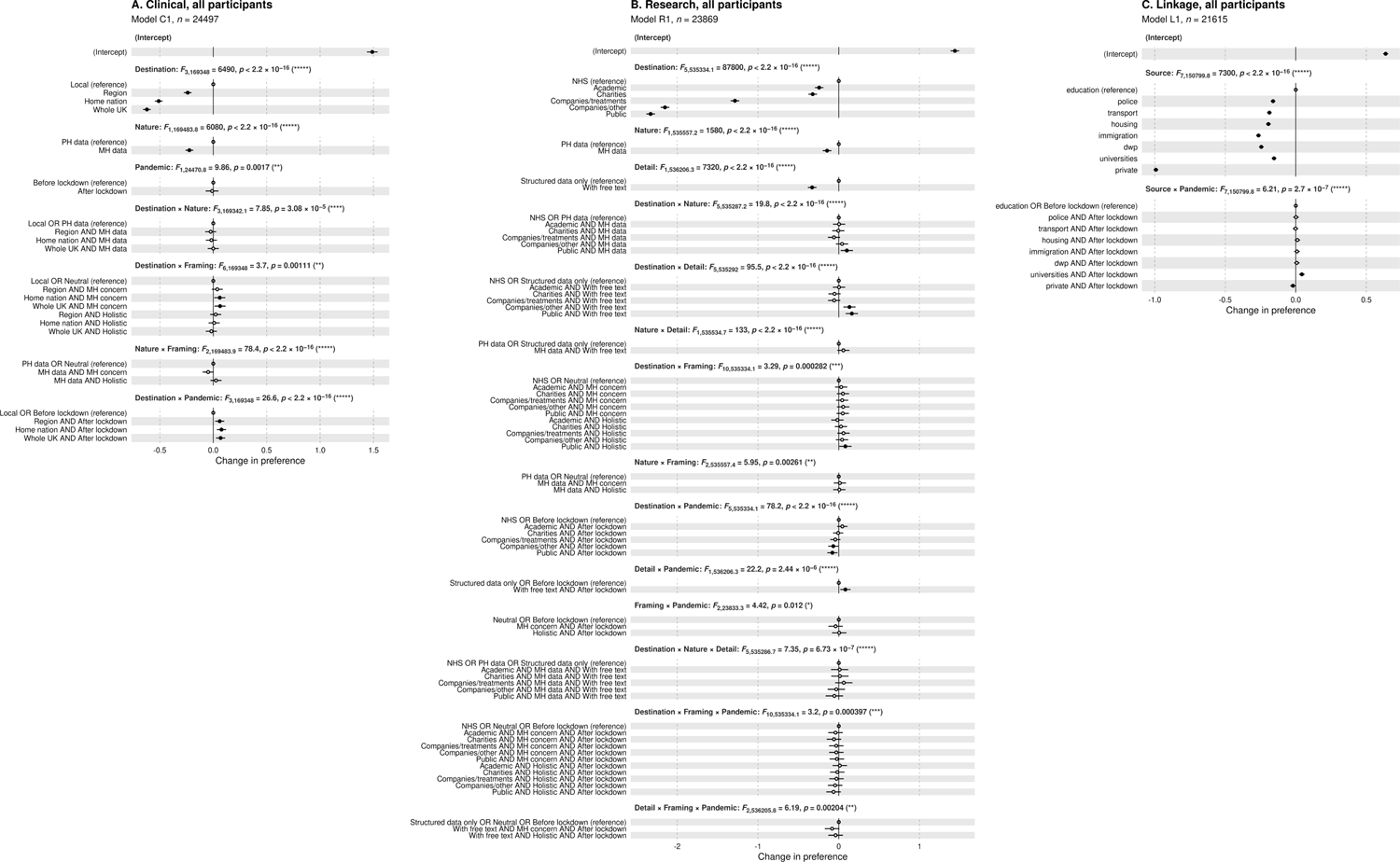
Effect sizes for predictors of willingness to **(A)** share data for clinical purposes (compare **Figure 3C**), **(B)** share data for research purposes (compare **Figure 3F,H**), or **(C)** support linkage to non-health data for research (compare **Figure 4**). Analysis via models C1, R1, and L1 respectively. These models include all participants, including those not supplying full demographic details, and therefore do not include the demographic predictors; compare **Figure 6**. Conventions as for **Figure 6**.

**Supplementary Figure 7.**
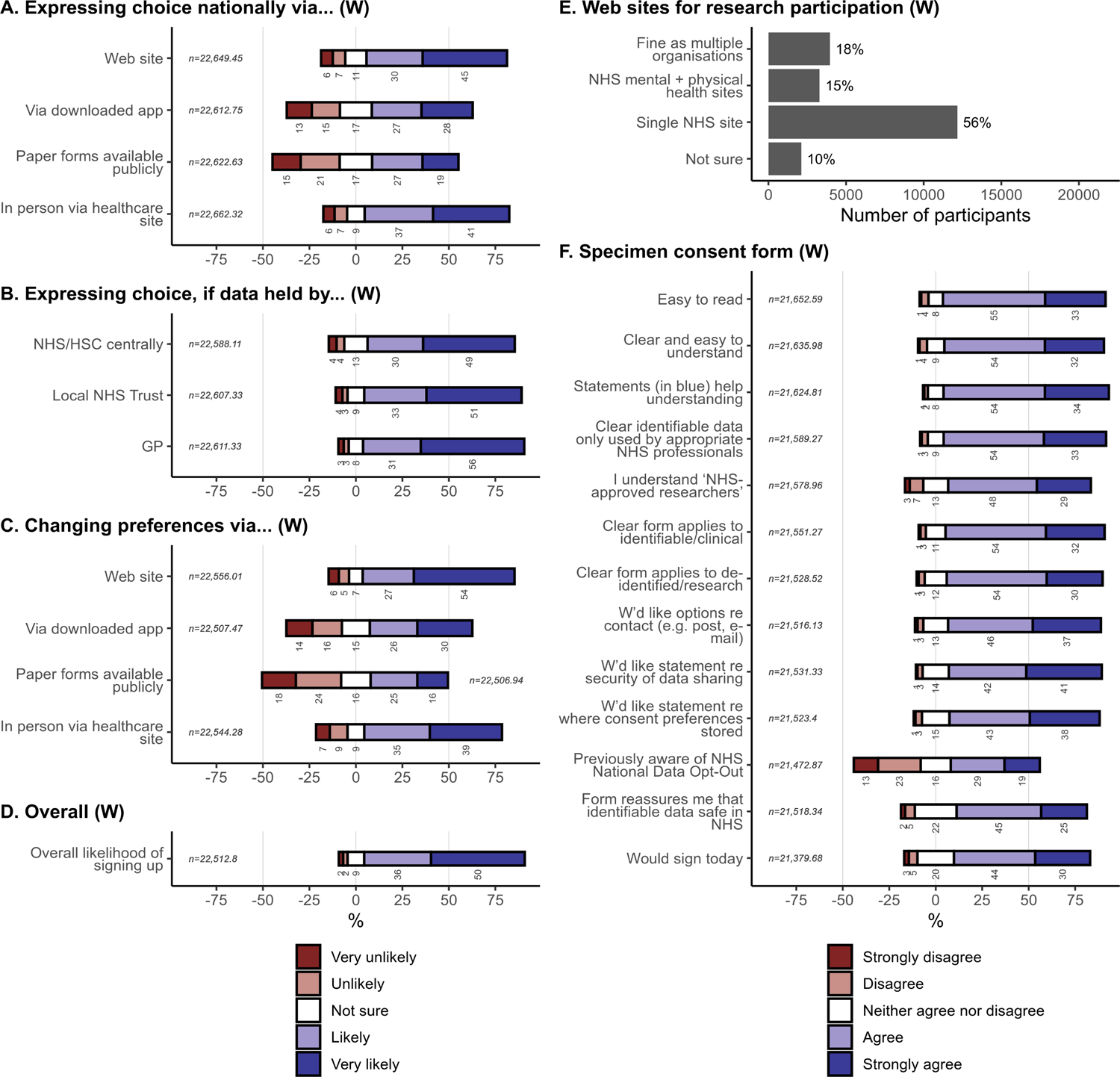
Views on a national data sharing consent system, weighted according to demographic variables. This is equivalent to **Figure 7** apart from the weighting (see **Supplementary Methods**). “(W)”: weighted.

